# Physiological Waveform Features Based Aging Clocks Associate with Cardiovascular Disease

**DOI:** 10.64898/2026.11.03.26367198

**Authors:** Ravi Pal, Anna Barney, Maxime Cannesson

## Abstract

Aging affects the morphology of photoplethysmography (PPG) and arterial blood pressure (ABP) waveforms. Here, we demonstrate that machine learning (ML) models trained on features derived from PPG or ABP waveforms can predict an individual’s age. We further investigated whether the discrepancy between predicted and chronological age (Age-Gap) could serve as a measure of cardiovascular health by testing its association with cardiovascular disease (CVD). From a large perioperative dataset of 17,327 surgical patients, high-fidelity PPG and ABP waveforms were analyzed, and over 840 time-, frequency-, and statistical-domain features per cardiac cycle were extracted using a signal-processing-based tool. These features were then used to develop ML-based aging clocks, PPGAge and ABPAge, trained on PPG and ABP features, respectively. For each aging clock, we evaluated five ML models: Light Gradient Boosting Machine (LightGBM), eXtreme Gradient Boosting (XGBoost), Random Forest, Lasso, and Elastic Net. Individuals were classified as having accelerated, normal, or delayed aging based on their Age-Gap relative to the model-specific mean absolute error (MAE, rounded up to the nearest whole year): accelerated aging was defined as Age-Gap ≥ +MAE, normal aging as −MAE ≤ Age-Gap < +MAE, and delayed aging as Age-Gap < −MAE. LightGBM provided the best age predictions for both aging clocks, with Pearson correlation coefficients (*r*) of 0.92 and 0.92 and *R*^*2*^ values of 0.81 and 0.82 for PPGAge and ABPAge, respectively. Moreover, we found that accelerated aging was significantly associated with higher odds of CVD for both aging clocks, PPGAge (odds ratio, 1.87 [95% CI, 1.59-2.21], *p<0*.*001*) and ABPAge (odds ratio, 2.28 [95% CI, 1.51-3.43], *p<0*.*001*). Additionally, delayed aging was significantly associated with lower odds of CVD for PPGAge (odds ratio, 0.68 [95% CI, 0.58-0.80], *p<0*.*001*). Our findings suggest that aging clocks based on high-fidelity PPG or ABP waveform features may serve as novel biomarkers for identifying individuals with higher odds of CVD.

## Introduction

In this study, we investigated the potential of photoplethysmography (PPG) and separately of arterial blood pressure (ABP) waveform features for predicting an individual’s age using machine learning (ML). The scientific basis of our hypothesis is that aging alters arterial compliance, which in turn changes the shape of these waveforms^1-9^. Previous studies have shown that the difference between predicted and chronological age, referred to as the Age-Gap, reflects biological aging. We therefore investigated whether the Age-Gap is associated with cardiovascular disease (CVD).

Aging is a universal biological process associated with gradual physical decline, which increases the risk of disease and death^10-12^. It also guides a broad spectrum of clinical and policy decisions, from disease screening and management to the determination of insurance and retirement benefits^13^. Although all individuals age chronologically, the rate of biological aging, also referred to as physiological aging, differs widely among individuals^14,15^. While chronological age indicates how long an organism has lived, biological age reflects the gradual decline in functional capacity of a person or organ and captures the cumulative effects of time, genetics, environmental, lifestyle, and other unknow variables that affect aging^16,17^. Although chronological age is a major risk factor for chronic diseases and mortality^18^, it is an imperfect measure of the aging process^19^, as individuals of the same chronological age may experience aging very differently^15,18,20,21^. Some experience physiological decline and require assistance with daily activities relatively young, whereas many remain independent and show little deterioration until a very advanced chronological age^18^, reflecting the heterogeneous nature of aging^21,22^. Biological aging better captures this heterogeneity^23,24^; however, measuring the aging process beyond chronological age remains challenging^13,25^. In recent years, several aging biomarkers have been proposed using machine learning (ML) methods^26-28^, including groundbreaking studies that have shown that DNA methylation patterns serve as powerful epigenetic biomarkers of aging^29,30^. However, one major issue with epigenetic clocks is their generalizability across different tissues, populations, and ethnicities^31^. Moreover, they are limited in interpretability, as they provide averaged estimates that do not indicate which cell types in a tissue drive aging^32^. More recently, aging clocks based on physiological waveforms, including wearable photoplethysmography (PPG)^13^, electrocardiograms (ECGs)^16,17,33-36^, electroencephalograms (EEGs)^37-40^, and polysomnograms (PSGs)^41^, have also been introduced. However, these approaches are often validated on small or less diverse datasets, which may limit their ability to generalize to broader populations^35,36^. In addition, it is often unclear which specific waveform parameters drive the predictions^13,17,34^, which limits interpretability and explainability.

High-fidelity physiological waveforms, such as PPG and ABP, provide valuable insights into cardiovascular health^42,43-46^. ABP is a fundamental hemodynamic parameter, frequently used to guide therapeutic interventions, particularly in critically ill patients^47-49^. Continuous ABP is measured using an invasive arterial line, which involves inserting a small catheter into an artery^48^. It can be monitored at various anatomical sites, including the radial, brachial, or femoral arteries^48^. PPG is a non-invasive optical technique that closely mirrors the ABP waveform morphology^44^. However, it measures blood volume changes within the microvascular tissue bed and is frequently used in anesthesia monitoring to assess blood oxygenation levels (SpO_2_)^50^. It is obtained using a light source and a photo-detector placed on the skin^44,51^, typically at sites such as the fingers, ears, wrists, or toes^50,52^. Studies have shown that aging alters the morphology of both PPG and ABP waveforms^42,53,54-65^. For instance, the dicrotic notch, a key marker of systolic and diastolic events, gradually weakens or disappears with age (see Fig. 1)^5-7,56,57,60-62^. Therefore, features extracted from these waveforms could play a crucial role in estimating an individual’s age.

**Fig. 1.**
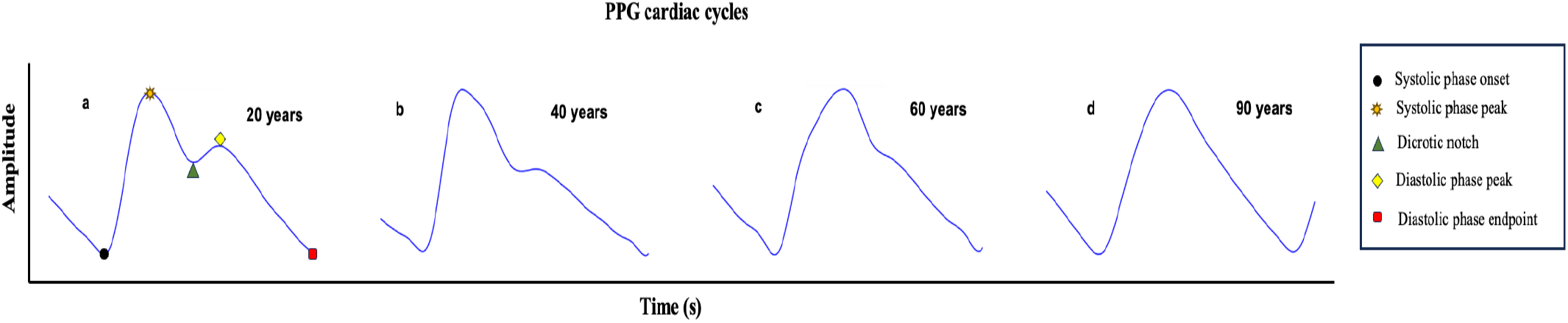
Typical shapes of photoplethysmography (PPG) waveform cardiac cycles in different age groups. **(a–d)**, PPG waveform cardiac cycles from individuals aged 20, 40, 60, and 90 years.

In this study, we developed ML-based aging clocks using features extracted from large real-world high-fidelity clinical physiological waveforms. We developed two aging clocks: PPGAge, based on PPG waveform features, and ABPAge, based on ABP waveform features (see Fig. 2). Using our recently developed feature extraction tool, we derived more than 840 features per cardiac cycle from these waveforms including time-domain, frequency-domain, and statistical features^50^. For each aging clock, we evaluated five ML models: Light Gradient-Boosting Machine (LGBM)^66^, eXtreme Gradient Boosting (XGBoost)^67^, Random Forest^68^, Lasso^69^, and Elastic Net^70^. Additionally, to improve model interpretability, we employed SHapley Additive exPlanations (SHAP) to explain the output of the best performing ML model^71^. SHAP analysis provides information about how much each input feature (in our study, a PPG feature or an ABP feature) contributes to the model output^72^. Furthermore, we assessed the clinical relevance of the aging clocks using the Age-Gap, defined as the difference between predicted and actual chronological age. The Age-Gap provides information on an individual’s physiology and has been shown to be associated with overall health status^13,16,34^. We therefore investigated, for both aging clocks (PPGAge and ABPAge), whether the Age-Gap (PPGAge Gap or ABPAge Gap) was associated with CVD.

**Fig. 2.**
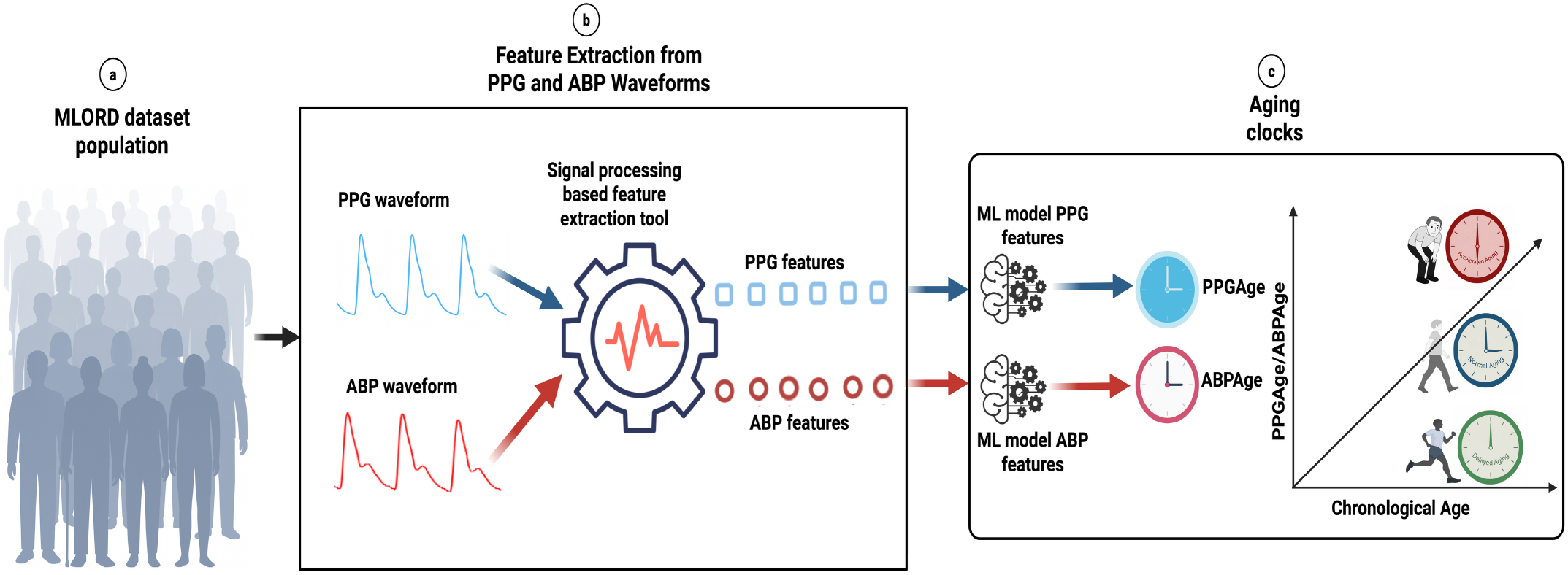
Study overview. **a-c**, Development of machine learning (ML)–based PPGAge and ABPAge aging clocks using photoplethysmography (PPG) waveform features and arterial blood pressure (ABP) waveform features, respectively. **a**, MLORD (machine learning of physiological waveforms and electronic health record data) dataset used in this study. **b**, Feature extraction from PPG and ABP waveforms using a signal processing–based tool that computes >840 features per cardiac cycle, including time-domain, frequency-domain, and statistical characteristics. **c**, Development of aging clocks.

The contributions of this study are as follows:

- Development of PPGAge and ABPAge aging clocks, derived from PPG features and ABP features, respectively, to predict individual’s age.
- The aging clocks were evaluated using a large clinical dataset, the perioperative Machine Learning of Physiological Waveforms and Electronic Health Record Data (MLORD) dataset, which comprises data from 17,327 patients^73^.
- Application of an explainable ML approach (SHAP) to evaluate the contribution of PPG features to the PPGAge aging clock and ABP features to the ABPAge aging clock.
- We demonstrate that accelerated aging, defined as an Age-Gap ≥ the model-specific mean absolute error (MAE; LightGBM), is significantly associated with higher odds of CVD for both the PPGAge and ABPAge aging clocks. Moreover, delayed aging, defined as an Age-Gap < −MAE, is significantly associated with lower odds of CVD, for PPGAge aging clock.

## Results

### PPGAge and ABPAge aging clocks to predict person’s age

The performance of the aging clocks in predicting person’s age is summarized in Table 1. The table reports the Pearson correlation coefficient (*r*), coefficient of determination (*R*^*2*^), mean absolute error (MAE), and root mean square error (RMSE)^74-76^. Across both aging clocks, LightGBM demonstrated slightly better predictive performance than the other four evaluated ML models (see Table 1). The LightGBM-based PPGAge aging clock achieved an *r* of 0.92, *R*^*2*^ of 0.81, MAE of 5.90, and RMSE of 7.53, and the LightGBM-based ABPAge aging clock achieved an *r* of 0.92, *R*^*2*^ of 0.82, MAE of 5.83, and RMSE of 7.31. Figure 3a and Figure 3b show the actual chronological age-versus-predicted age scatter plots for the PPGAge and ABPAge aging clocks, respectively. Moreover, Bland-Altman plots comparing chronological and predicted age for the PPGAge and ABPAge aging clocks are shown in Fig. 4(a) and (b), respectively. Both aging clocks demonstrate a mean bias close to zero, with limits of agreement of ± 14.77 years for PPGAge and ± 14.33 years for ABPAge. Data points in the scatter plots (Fig. 3a, b) and Bland–Altman plots (Fig. 4a, b) are color-coded according to Age-Gap (predicted aging-clock age − chronological age), classifying individuals as having accelerated (red, Age-Gap ≥ +6 years), normal (blue, −6 ≤ Age-Gap < +6 years), or delayed (green, Age-Gap < −6 years) aging. The ±6-year threshold was derived from the respective LightGBM model’s MAE (PPGAge MAE = 5.90 years; ABPAge MAE = 5.83 years), rounded up to the nearest year. For the PPGAge and ABPAge clocks, Age-Gap is referred to as the PPGAge Gap and ABPAge Gap, respectively.

**Table 1.** Performance of the PPGAge and ABPAge aging clocks on held out test sets.

| Aging clock | Number of patients | ML model | Performance evaluators |  |  |  |
|---|---|---|---|---|---|---|
|  |  |  | *r* | *R*<sup>2</sup> | *MAE* | *RMSE* |
| PPGAge | *n*=6,699 | **LightGBM** | **0.92** | **0.81** | **5.90** | **7.53** |
|  |  | XGBoost | 0.91 | 0.79 | 6.30 | 8.03 |
|  |  | Random Forest | 0.91 | 0.79 | 6.30 | 8.03 |
|  |  | Lasso | 0.90 | 0.76 | 6.73 | 8.53 |
|  |  | Elastic Net | 0.90 | 0.76 | 6.71 | 8.51 |
| ABPAge | *n*=884 | **LightGBM** | **0.92** | **0.82** | **5.83** | **7.31** |
|  |  | XGBoost | 0.91 | 0.80 | 6.05 | 7.58 |
|  |  | Random Forest | 0.92 | 0.81 | 5.91 | 7.38 |
|  |  | Lasso | 0.91 | 0.79 | 5.88 | 7.72 |
|  |  | Elastic Net | 0.91 | 0.79 | 5.92 | 7.77 |

PPG photoplethysmography, ABP arterial blood pressure, ML machine learning, LightGBM Light Gradient-Boosting Machine, XGBoost eXtreme Gradient Boosting, Pearson correlation coefficient (*r*), coefficient of determination (*R*<sup>2</sup>), mean absolute error (MAE), and root mean square error (RMSE).

**Fig. 3.**
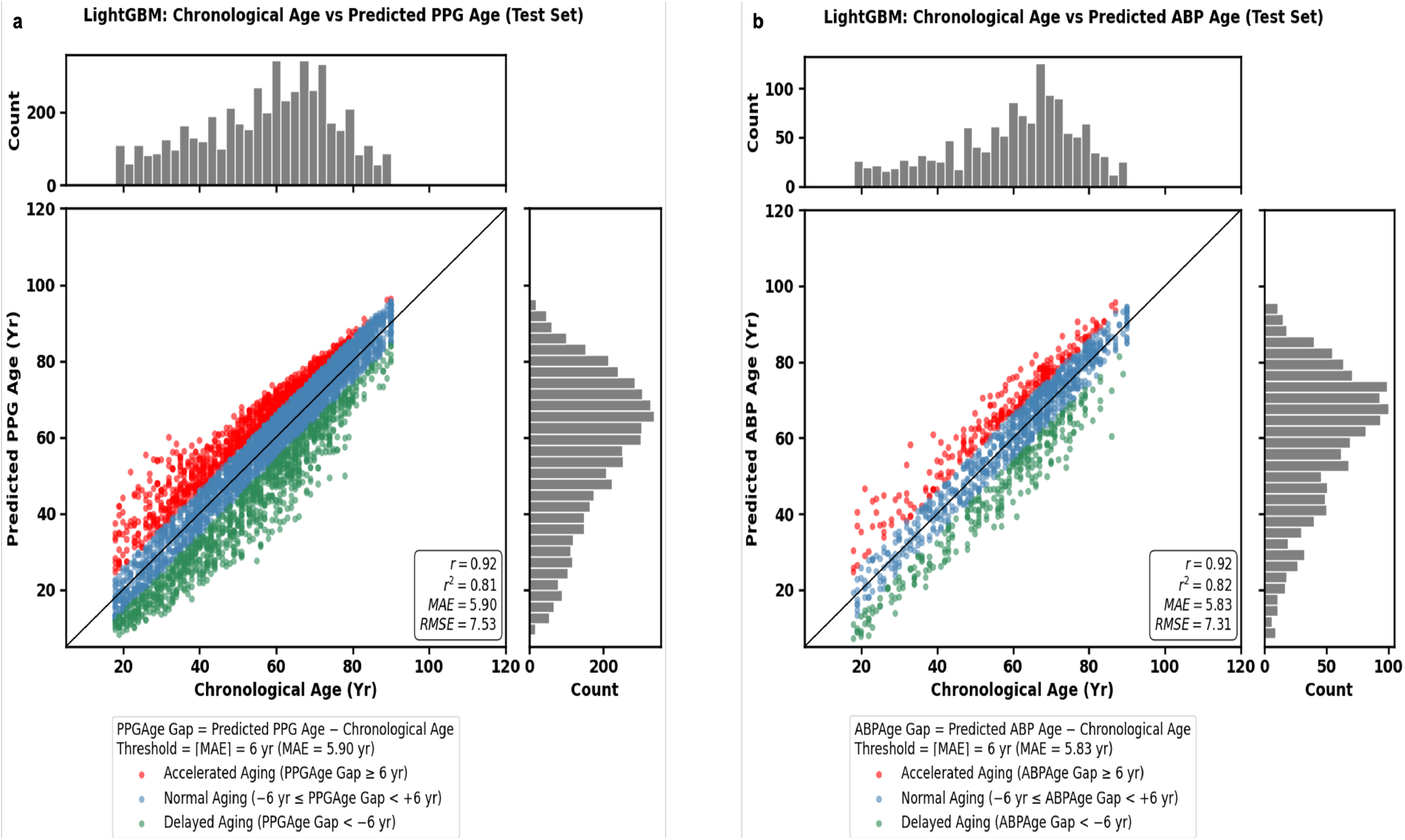
Scatter plots showing the age predictions of the PPGAge and ABPAge aging clocks. **a** Scatter plot of predicted PPG age (PPGAge) against chronological age for the LightGBM model in the held-out test set (n = 4,999). **b** Scatter plot of predicted ABP age (ABPAge) against chronological age for the LightGBM model in the held-out test set (n = 1,357). For both aging clocks, points are colored by Age-Gap (predicted minus chronological age) relative to the model’s mean absolute error (MAE): red, accelerated aging (Age-Gap ≥ MAE); blue, normal aging (−MAE ≤ Age-Gap < +MAE); green, delayed aging (Age-Gap< -MAE). The Age-Gap is referred to as the PPGAge Gap for the PPGAge aging clock and as the ABPAge Gap for the ABPAge aging clock. In our analysis, the model’s MAE was rounded up to the next whole number. Black line represents the identity line (predicted age = chronological age). Top and right marginal histograms show the distributions of chronological age and predicted age, respectively.

**Fig. 4.**
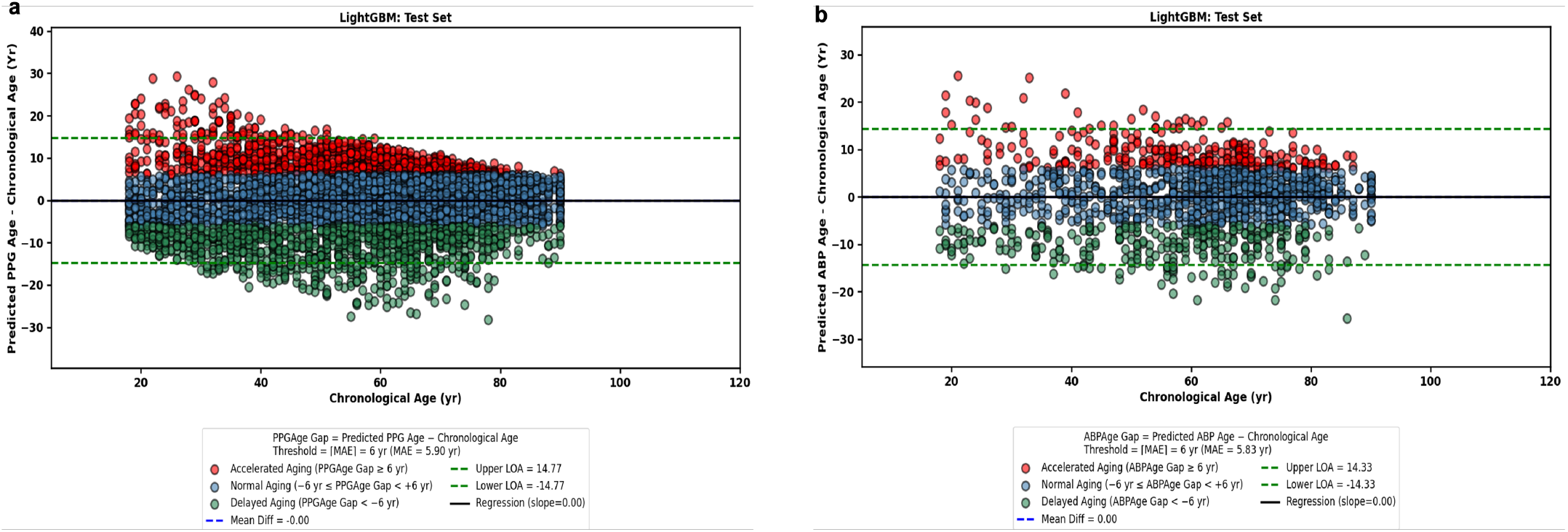
Bland Altman plots. Agreement between chronological and predicted age for (a) PPGAge (n = 4,999) and (b) ABPAge (n = 1,357) aging clocks. Points are coloured by Age-Gap group (red = accelerated aging, blue = normal aging, green = delayed aging).

### SHAP method to explain the prediction of PPGAge and ABPAge aging clocks

SHAP was used to evaluate the contributions of individual PPG and ABP features to age prediction using the PPGAge and ABPAge aging clocks, respectively. Figure 5 (a) and (b) presents the SHAP summary plots for PPGAge and ABPAge aging clocks, respectively. Each plot displays the top 10 most important features in descending order of importance, where features positioned higher on the y-axis have a greater influence on the model’s predictions of individual’s age. The complete description of these features is provided in Table 2. In addition to ranking feature importance, the plots also show the distribution of SHAP values for each feature across all samples, thereby highlighting both the magnitude and direction of their effects. A positive SHAP value indicates that the feature increases the predicted value, whereas a negative SHAP value indicates that the feature decreases it. The distance from zero reflects the magnitude of this contribution, with points farther from zero indicating a stronger influence on the prediction. Each point represents an individual sample, and its colour reflects the corresponding feature value, with high values shown in red and low values in blue. This visualization provides insight into how variations in feature values influence the model’s predictions.

**Table 2.** Descriptions of the features shown in Fig. 5 (a-b) that contributed most to the PPGAge and ABPAge aging clocks.

| **Feature** | **Description** |
|---|---|
| PPG_AM_SD_Bpeak_wrtDpeak_mean | PPG cardiac cycle second derivative Bpeak amplitude with respect to Dpeak, averaged across cardiac cycles |
| PPG_D_RT_NAre_wsDIAPE_SYSPO_wrtDIAN_PPG_D_NAre_wsDIAPE_SYSPO_wrtSYSPP_mean | The ratio of the area under the normalized PPG cardiac cycle systolic phase onset with respect to dicrotic notch, with subtracted diastolic phase end point to the area under the normalized PPG cardiac cycle systolic phase onset with respect to systolic phase peak, with subtracted diastolic phase end point, averaged across cardiac cycles |
| PPG_D_RT_NAre_SYSPP_wrtDIAN_PPG_D_NAre_SYSPO_wrtDIAPP_mean | The ratio of the area under the normalized PPG cardiac cycle systolic phase peak with respect to dicrotic notch to the area under the normalized PPG cardiac cycle systolic phase onset with respect to diastolic phase peak, averaged across cardiac cycles |
| PPG_D_NMed_SYSPO_wrtDIAPE_std | Standard deviation across cardiac cycles of the median duration of the normalized PPG cardiac cycle systolic phase onset with respect to diastolic phase end point |
| PPG_D_Are_SYSPO_wrtSYSPP_mean | Area under the PPG cardiac cycle systolic phase onset with respect to systolic phase peak, averaged across cardiac cycles |
| PPG_AM_SD_Bpeak_wrtzero_mean | PPG cardiac cycle second derivative Bpeak amplitude with respect to zero, averaged across cardiac cycles |
| PPG_D_RT_NAre_wsDIAPE_SYSPP_wrtDIAN_PPG_D_NAre_wsDIAPE_SYSPO_wrtDIAPP_mean | The ratio of the area under the normalized PPG cardiac cycle systolic phase peak with respect to dicrotic notch, with subtracted diastolic phase end point to the area under the normalized PPG cardiac cycle systolic phase onset with respect to diastolic phase peak, with subtracted diastolic phase end point, averaged across cardiac cycles |
| PPG_AM_RT_DIAPE_wrtSYSPO_PPG_DIAPE_wrtZero_std | Standard deviation across cardiac cycles of the ratio of the PPG cardiac cycle diastolic phase end point amplitude with respect to systolic phase onset to the PPG cardiac cycle diastolic phase endpoint amplitude with respect to zero |
| PPG_D_NRMS_SYSPO_wrtDIAPE_mean | Root mean square duration of the normalized PPG cardiac cycle systolic phase onset with respect to diastolic phase end point, averaged across cardiac cycles |
| PPG_D_RT_Are_wsDIAPE_SYSPP_wrtDIAN_PPG_D_Are_wsDIAPE_SYSPO_wrtDIAPP_mean | The ratio of the area under the PPG cardiac cycle systolic phase peak with respect to dicrotic notch, with subtracted diastolic phase end point to the area under the PPG cardiac cycle systolic phase onset with respect to diastolic phase peak, with subtracted diastolic phase end point, averaged across cardiac cycles |
| ABP_pkamp_std | Standard deviation across cardiac cycles of the amplitude of the first peak observed in the FFT of the ABP cardiac cycle |
| ABP_pkamp_mean | Mean across cardiac cycles of the amplitude of the first peak observed in the FFT of the ABP cardiac cycle |
| ABP_AM_RT_SD_Dpeak_wrtEpeak_ABP_AM_SD_Cpeak_wrtEpeak_std | Standard deviation across cardiac cycles of the ratio of the ABP cardiac cycle second derivative Dpeak amplitude with respect to Epeak to the ABP cardiac cycle second derivative Cpeak amplitude with respect to Epeak |
| ABP_AM_RT_DIAPE_wrtZero_ABP_SYSPO_wrtZero_mean | The ratio of the ABP cardiac cycle diastolic phase end point amplitude with respect to zero to the ABP cardiac cycle systolic phase onset amplitude with respect to zero, averaged across cardiac cycles |
| ABP_D_FD_Upeak_wrtVpeak_mean | Duration of the ABP cardiac cycle first derivative Upeak with respect to Vpeak, averaged across cardiac cycles |
| ABP_AM_RT_DIAPE_wrtSYSPO_ABP_DIAPE_wrtZero_std | Standard deviation across cardiac cycles of the ratio of the ABP cardiac cycle diastolic phase end point amplitude with respect to systolic phase onset to the ABP cardiac cycle diastolic phase endpoint amplitude with respect to zero |
| ABP_D_Avg_wsDIAPE_DIAPP_wrtDIAPE_std | Standard deviation across cardiac cycles of the average duration of the ABP cardiac cycle diastolic phase peak with respect to diastolic phase end point, with subtracted diastolic phase end point |
| ABP_AM_RT_SD_Apeak_wrtCpeak_ABP_AM_SD_Cpeak_wrtzero_mean | The ratio of the ABP cardiac cycle second derivative Apeak amplitude with respect to Cpeak to the ABP cardiac cycle second derivative Cpeak amplitude with respect to zero, averaged across cardiac cycles |
| ABP_AM_RT_DIAPE_wrtZero_ABP_SYSPO_wrtZero_std | Standard deviation across cardiac cycles of the ratio of the ABP cardiac cycle diastolic phase end point amplitude with respect to zero to the ABP cardiac cycle systolic phase onset amplitude with respect to zero |
| ABP_D_Avg_wsDIAPE_SYSPP_wrtDIAPP_std | Standard deviation across cardiac cycles of the average duration of the ABP cardiac cycle systolic phase peak with respect to diastolic phase peak, with subtracted diastolic phase end point |

*PPG* photoplethysmography, *ABP* arterial blood pressure.

**Fig. 5.**
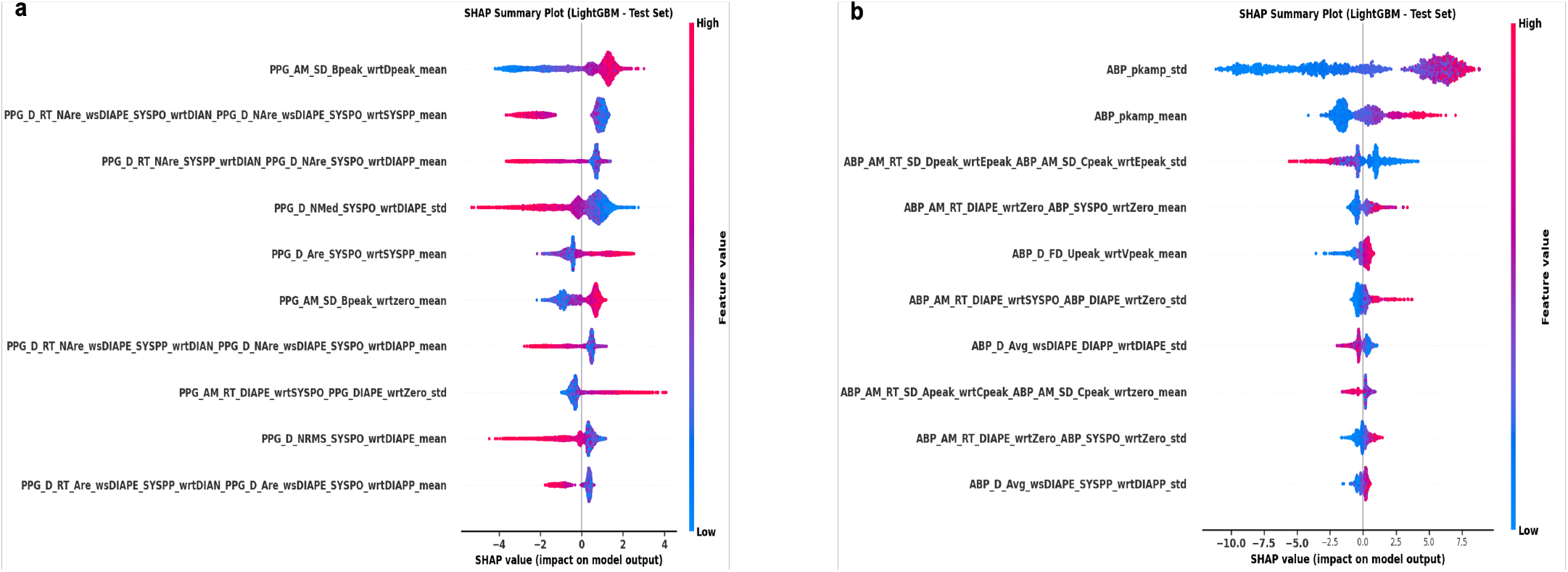
Shapley Additive exPlanation (SHAP) summary plots showing feature contributions to the PPGAge and ABPAge aging clocks. **a**, Contributions of PPG features to the PPGAge clock. **b**, Contributions of ABP features to the ABPAge clock.

In the PPGAge clock, the most influential feature contributing to age prediction was the PPG cardiac cycle second derivative Bpeak amplitude with respect to the Dpeak (Figure 5a). In the ABPAge clock, the feature with the greatest impact was the amplitude of the first peak in the Fast Fourier Transform (FFT) of the ABP cardiac cycle (Figure 5b). Figure 6 provides a visual representation of these two features. To examine how PPG and ABP features vary across Age-Gap groups (delayed, normal, and accelerated aging), Fig. 7 displays box plots of the features ranked 1st, 5th, and 10th by SHAP importance for PPGAge (Fig. 7a–c) and ABPAge (Fig. 7d–f). Pairwise comparisons between Age-Gap groups were significant for each of the features shown in Fig. 7 (p < 0.001, two-sided t-test), indicating differences across the three Age-Gap groups.

**Fig. 6.**
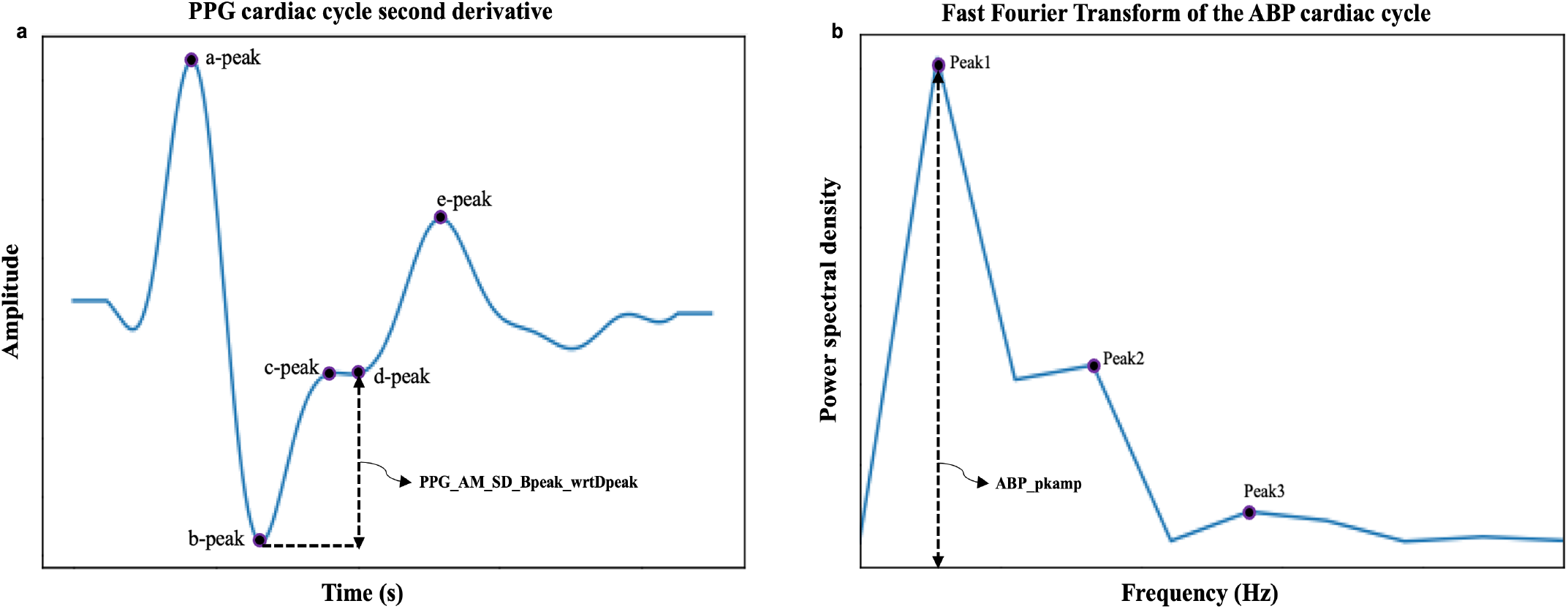
PPG feature contributing most to the PPGAge aging clock and ABP feature contributing most to the ABPAge aging clock. **a**, Second derivative of the PPG cardiac cycle with key peaks (a-peak, b-peak, c-peak, d-peak, and e-peak), highlighting the feature PPG_AM_SD_Bpeak_wrtDpeak (the amplitude of the b-peak relative to the d-peak in the second derivative), which contributed most to the PPGAge aging clock. **b**, Fast Fourier transform (FFT) of the ABP cardiac cycle, showing the feature ABP_pkamp (the amplitude of the first peak in the FFT of the ABP cardiac cycle), which contributed most to the ABPAge aging clock.

**Fig. 7.**
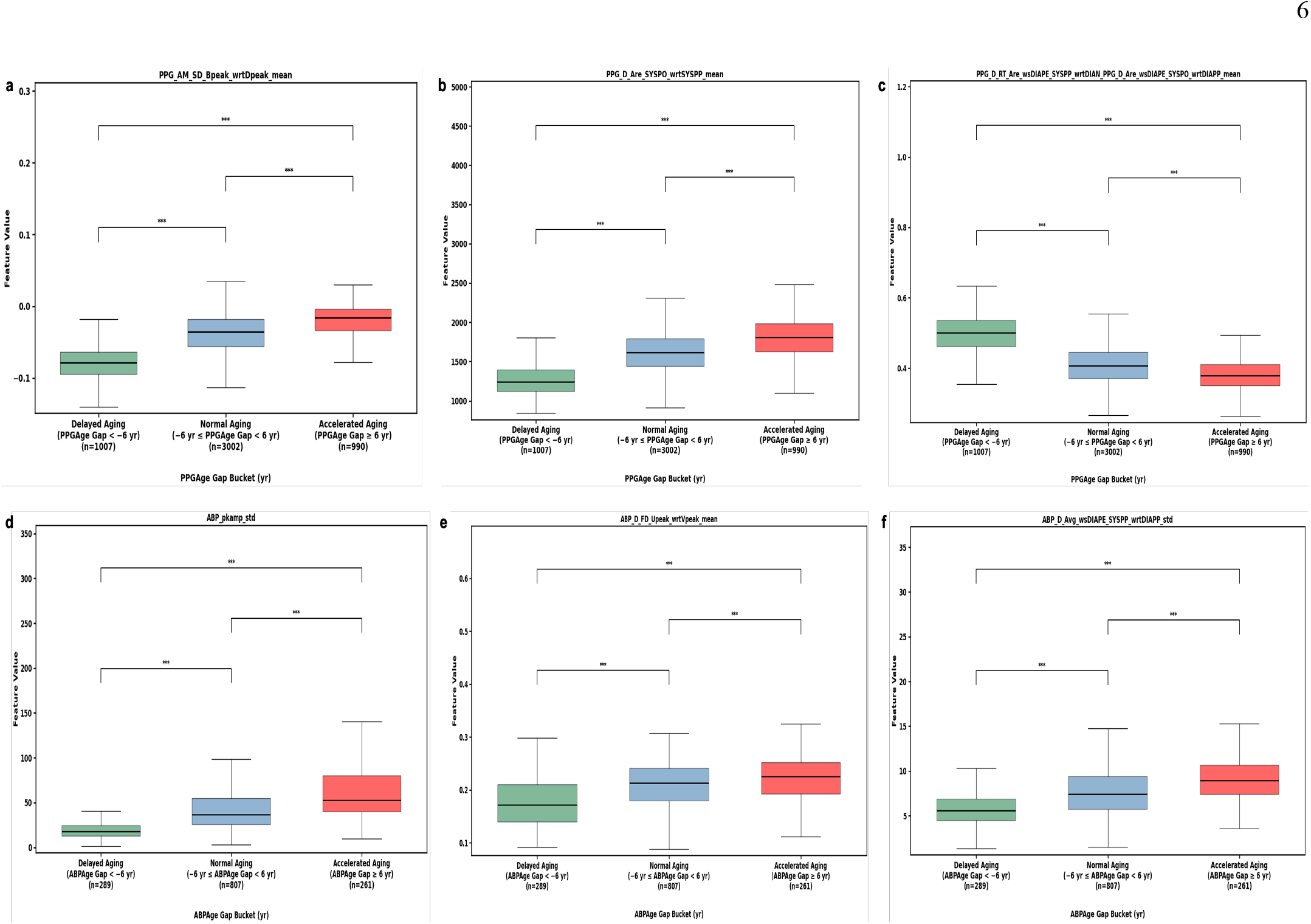
Box plots illustrating PPG and ABP waveform features across three Age-Gap groups: delayed, normal, and accelerated aging. **a–c**, PPG waveform features ranked 1st, 5th, and 10th by SHAP importance for the PPGAge clock (Fig. 5a), respectively. **d–f**, ABP waveform features ranked 1st, 5th, and 10th by SHAP importance for the ABPAge clock (Fig. 5b), respectively. Boxplots display significant differences between Age-Gap groups, denoted by asterisks (***), indicating a highly significant statistical difference (p < 0.001) based on a two-sided t-test.

### Age-Gap associates with cardiovascular disease

A clear association between Age-Gap and CVD was observed for both the PPGAge and ABPAge aging clocks, as illustrated by the spline curves in Fig. 8a and b, respectively. Moreover, Table 3 shows the associations of accelerated and delayed aging (Age-Gap groups, see Methods for the further details) with CVD for the PPGAge and ABPAge aging clocks, with normal aging serving as the reference category. The analyses were adjusted for chronological age and sex.

**Table 3.** Odds ratios for the association of accelerated and delayed aging defined by the PPGAge and ABPAge aging clocks with cardiovascular disease.

| **Aging clock** | **Event** | **Accelerated aging** |  | **Delayed aging** |  |
|---|---|---|---|---|---|
|  |  | **OR (95% CI)** | ***p-value*** | **OR (95% CI)** | ***p-value*** |
| PPGAge | CVD | 1.87 (1.59-2.21) | *p<0.001* | 0.68 (0.58-0.80) | *p<0.001* |
| ABPAge | CVD | 2.28 (1.51-3.43) | *p<0.001* | 0.82 (0.59-1.14) | *p=0.234* |

Odds ratios (ORs) and 95% confidence intervals (CIs) were estimated using logistic regression models. Results are adjusted for chronological age and sex. Normal aging was used as the reference category. *CVD* cardiovascular disease.

**Fig. 8.**
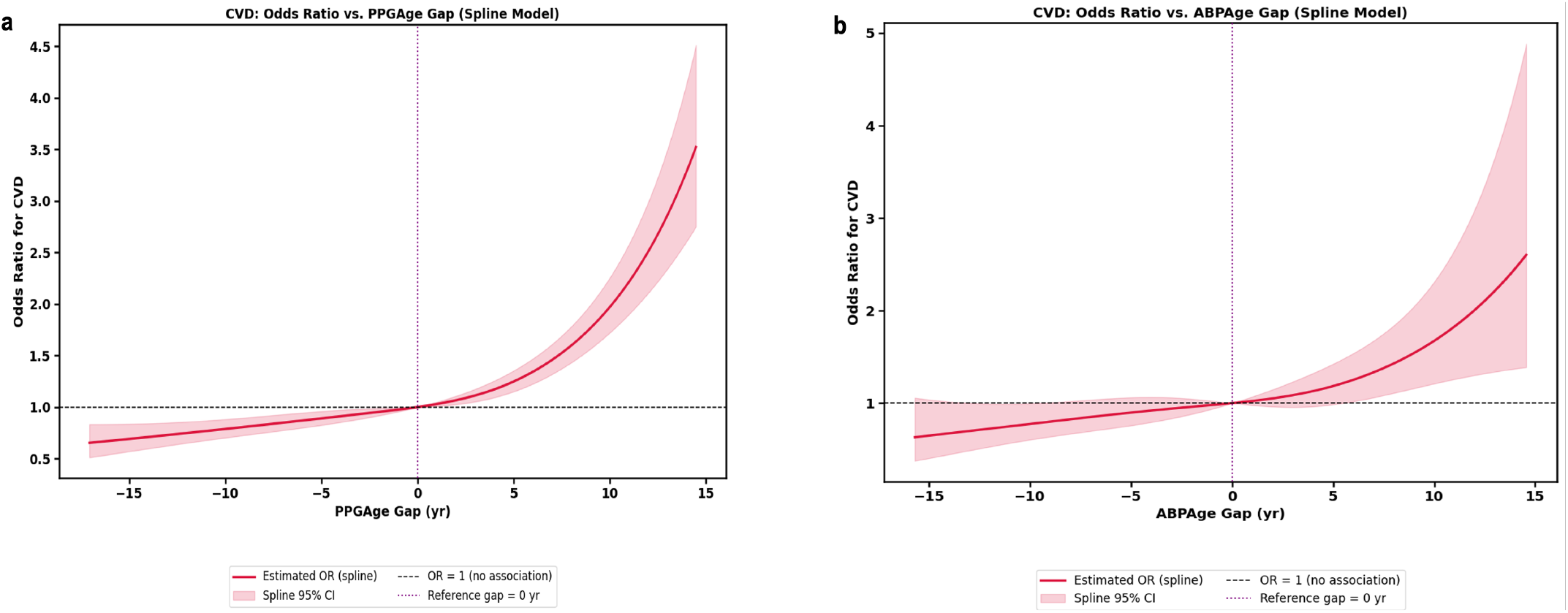
Spline curves showing the relationship between Age-Gap and Odds ratio for CVD. **a**, PPGAge Gap vs. odds ratio for CVD. **b**, ABPAge Gap vs. odds ratio for CVD.

Accelerated aging was significantly associated with higher odds of CVD for PPGAge (odds ratio (OR), 1.87 [95% CI, 1.59-2.21], *p<0*.*001*) and ABPAge (OR, 2.28 [95% CI, 1.51-3.43], *p<0*.*001*). Delayed aging was linked to reduced odds of having CVD for PPGAge (OR, 0.68 [95% CI, 0.58-0.80], *p<0*.*001*). For ABPAge, delayed aging did not show a significant association with CVD (OR, 0.82 [95% CI, 0.59-1.14], *p=0*.*234*).

When separated by sex (Table 4), accelerated aging based on the PPGAge clock was associated with higher odds of CVD in both females (OR, 1.76 [95% CI, 1.43-2.16], *p<0*.*001*) and males (OR, 2.08 [95% CI, 1.57-2.77], *p<0*.*001*). Delayed aging was associated with lower odds of CVD in both females (OR, 0.68 [95% CI, 0.53-0.87], *p=0*.*002*) and males (OR, 0.69 [95% CI, 0.56-0.85], *p<0*.*001*). For the ABPAge clock, accelerated aging was significantly associated with elevated odds of CVD in both females (OR, 2.50 [95% CI, 1.45-4.30], *p<0*.*001*) and males (OR, 2.03 [95% CI, 1.09-3.80], *p=0*.*027*), although, we did not find significant association between delayed aging and CVD in either females (OR, 0.96 [95% CI, 0.59-1.59], *p=0*.*887*) or males (OR, 0.71 [95% CI, 0.46-1.09], *p=0*.*121*) for ABPAge clock.

**Table 4.** Odds ratios for the association of accelerated and delayed aging defined by the PPGAge and ABPAge aging clocks with cardiovascular disease, separated by sex.

| **Aging clock** | **Event** | **Accelerated aging** |  |  |  | **Delayed aging** |  |  |  |
|---|---|---|---|---|---|---|---|---|---|
|  |  | **Females** |  | **Males** |  | **Females** |  | **Males** |  |
|  |  | **OR (95% CI)** | ***p-value*** | **OR (95% CI)** | ***p-value*** | **OR (95% CI)** | ***p-value*** | **OR (95% CI)** | ***p-value*** |
| PPGAge | CVD | 1.76 (1.43-2.16) | *p<0.001* | 2.08 (1.57-2.77) | *p<0.001* | 0.68 (0.53-0.87) | *P=0.002* | 0.69 (0.56-0.85) | *p<0.001* |
| ABPAge | CVD | 2.50 (1.45-4.30) | *p<0.001* | 2.03 (1.09-3.80) | p=0.027 | 0.96 (0.59-1.59) | *p=0.887* | 0.71 (0.46-1.09) | *P=0.121* |

Odds ratios (ORs) and 95% confidence intervals (CIs) were estimated using logistic regression models. Results are presented separately for males and females, with adjustments made for chronological age. Normal aging was used as the reference category. *CVD* cardiovascular disease.

We also observed a strong association between Age-Gap– defined aging groups (accelerated and delayed aging) and CVD diagnostic rates for both the PPGAge and ABPAge aging clocks, with the strongest associations observed in the 18–50-year-old age group among both female and male patients (Fig. 9). For PPGAge, the baseline rate of CVD diagnosis among 18– 50-year-old women was 31.50% (95% CI 28.60-34.60). However, among women with accelerated aging in this age group, that rate increased to 49.01% (95% CI 43.08-54.94), or 1.56 (95% CI 1.37-1.75) times the average rate; we refer to this ratio as the relative rate of diagnosis (see Methods for further details). Among women with delayed aging, that rate drops to 21.53% (95% CI 16.27-27.27), or a 0.68 (95% CI 0.52-0.87) relative rate of diagnosis (Fig. 9 (a, b)). A similar pattern was observed among 18–50-year-old men: the baseline rate was 39.10% (95% CI 35.40-42.60), rising to 61.07% (95% CI 53.02-69.13), or 1.56 (95% CI 1.36-1.77) times the average rate with accelerated aging and falling to 21.88% (95% CI 16.96-27.23) or a 0.56 (95% CI 0.43-0.70) times the average rate with delayed aging (Fig. 9 (c, d)).

**Fig. 9.**
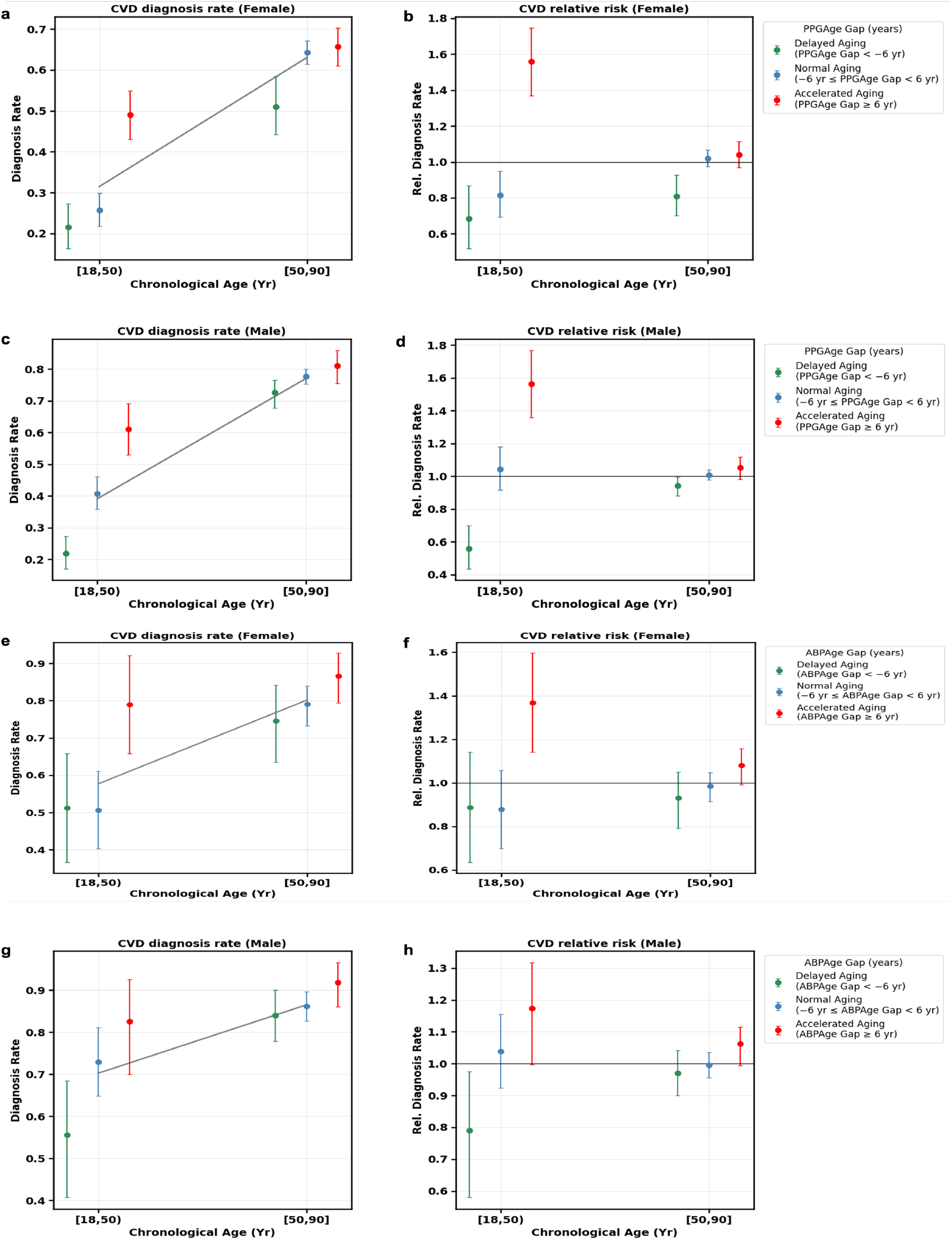
PPGAge Gap and ABPAge Gap groups and CVD diagnosis rates across chronological age brackets. CVD diagnosis rate (left panel of each pair) and relative diagnostic rate (right panel), stratified by Age-Gap group (delayed, normal and accelerated aging) across chronological age brackets ([18,50) and [50,90]). **(a, b)** PPGAge Gap: diagnosis rate and relative diagnostic rate for female (n = 2,462) and **(c, d)** male (n = 2,537) participants. **(e, f)** ABPAge Gap: diagnosis rate and relative diagnostic rate for female (n = 559) and **(g, h)** male (n = 798) participants. Age-Gap is predicted minus chronological age. The grey line (left panels) shows the overall diagnosis rate in each age bracket; the black line (right panels) marks a relative rate of 1. All intervals are 95% confidence intervals computed with 1000 bootstrap samples.

This association held for ABPAge as well. Among 18-50 year old women the baseline rate of CVD diagnosis was 57.70% (95% CI 50.00-65.40), increasing to 78.95% (95% CI 65.79-92.11) or 1.37 (95% CI 1.14-1.60) times the average rate with accelerated aging and decreasing to 51.22% (95% CI 36.59-65.85) or a 0.89 (95% CI 0.63-1.14) times the average rate with delayed aging (Fig. 9 (e, f). Among 18–50-year-old men, the baseline rate was 70.20% (95% CI 63.90-76.60), jumps to 82.50% (95% CI 70.00-92.50), or 1.17 (95% CI 1.00-1.32) times the average rate with accelerated aging and drops to 55.56% (95% CI 40.74-68.52) or a 0.79 (95% CI 0.58-0.98) times the average rate with delayed aging Fig. 9 (g, h).

## Discussion

In this study, we showed that ML-based aging clocks, PPGAge, and ABPAge developed using PPG features and ABP features, respectively, are highly predictive of individual’s age across a broad age range (18 to 90 years). Furthermore, we demonstrated physiological waveform (PPG- or ABP)-based aging clocks capture important information about cardiovascular health by showing an association between the difference between aging clock-predicted age and chronological age (referred to as the PPGAge Gap for the PPGAge clock and the ABPAge Gap for the ABPAge clock) and CVD.

We found that, both aging clocks achieved similar age prediction performance, with mean absolute errors (MAEs) of 5.90 and 5.83 years, respectively. Compared with existing physiological waveform-based aging clocks derived from ECG signals (MAE: 6.89-8 years^17,33-35^), EEG signals (MAE: 5.96-7.6 years^39,40,37^), and PSGs signals (MAE: 5.96 years^41^), their performance compares favourably. Recently, wearable device-based PPG aging clock was introduced by Miller et al.^13^ which achieved the MAE of 2.4 for healthy and 3.2 years for general groups. However, direct comparison with our study is not feasible, as their model was trained on wearable-derived PPG data from healthy participants and has yet to be evaluated on clinical PPG waveforms. Nie et al.^77^ presented a deep learning-based model to estimate AI-PPG age using clinical PPG waveforms for cardiovascular health assessment; however, they reported a moderate correlation between predicted AI-PPG age and chronological age (*r < 0*.*55*). In comparison, our study achieved a higher Pearson correlation (*r=0*.*92* for both PPGAge and ABPAge aging clocks) between aging clock predicted age and chronological age. As noted by Nie et al.^77^, closer alignment between predicted and chronological age may improve the interpretability in practical applications.

Moreover, our analysis shows that, for both PPGAge and ABPAge aging clocks, accelerated aging is associated with significantly higher odds of CVD (PPGAge: odds ratio (OR), 1.87 [95% CI, 1.59-2.21], *p<0*.*001* and ABPAge: OR, 2.28 [95% CI, 1.51-3.43], *p<0*.*001*) in chronological age and sex adjusted models. We also show that delayed aging is significantly associated with lower odds of CVD for PPGAge aging clock (OR, 0.68 [95% CI, 0.58-0.80], *p<0*.*001*). The strong association between Age-Gap groups (accelerated aging and delayed aging) and CVD suggests that physiological waveform-based aging clocks may provide information on cardiovascular health beyond what can be inferred from chronological age alone.

Feature extraction is a critical step in the analysis of physiological waveforms, as it enables the identification of clinically significant features and improves model interpretability and explainability^78^, key factors that support clinician trust^79^. The feature extraction method must capture relevant information even when waveform morphology changes due to disease, normal physiological variation, or aging. For example, a key feature of the cardiac cycle in these waveforms, the dicrotic notch, becomes less pronounced with aging ^5-7,56,57,60-62,80,81^.

Therefore, we used a signal processing-based feature extraction tool that not only provides a comprehensive set of more than 840 features per cardiac cycle, including time-domain, frequency-domain, and statistical characteristics from PPG and ABP waveforms, but also captures key features even when they are subtle or not clearly visible in the waveform, such as the dicrotic notch^50^. We applied the explainability method SHAP^71^ to identify the important features driving the age prediction. Previous studies have shown that features extracted from the second derivative of the PPG waveform are associated with aging^82-89^. Aligned with this, we found that the most important feature contributing to the PPGAge aging clock was derived from the second derivative of the PPG (see Fig. 5a). In the ABPAge aging clock, a frequency-domain feature contributed most to the model output (see Fig. 5b). Overall, these results demonstrate that physiological waveforms contain valuable information not only in raw signals but also in their derivatives and frequency-domain characteristics, providing important insights into individual’s age prediction.

In recent years, advancements in non-invasive PPG-based wearable technology and the development of continuous, non-invasive ABP monitoring devices have enabled the continuous acquisition of PPG and ABP waveforms, respectively, both in clinical settings and in daily life^90-99^. Our developed aging clocks could potentially be integrated into such devices as biomarkers of aging. This integration could contribute to improved patient outcomes and personalized healthcare monitoring, and help enhance the transparency and comprehensibility of these devices for medical professionals and end users by providing information on which physiological features contributed to the output.

Our study has several limitations. First, the aging clock models were developed and evaluated using a single centre clinical dataset, which may limit their generalizability. Future work will focus on validating the performance of these models on diverse, multicentre datasets. Additionally, we plan to assess their sensitivity over time through validation on a longitudinal dataset. Second, due to the unavailability of healthy subjects in our dataset, we trained the aging clocks using perioperative data. However, an ideal approach would involve developing the aging clocks on a healthy control group and then evaluating their performance on perioperative patients. This approach would allow us to better examine the Age-Gap between predicted and chronological age, which could provide a more accurate estimate of biological age and serve as a more reliable aging biomarker. Third, we did not evaluate the associations between Age-Gap (PPGAge Gap or ABPAge Gap) and individual types of CVD. In future work, we will focus on examining these associations across specific CVD subtypes to determine whether the relationships with Age-Gap differ by CVD type. Such analyses may help identify CVD subtypes that may be more strongly associated with accelerated aging. Fourth, while we identified the physiological waveform (PPG and ABP) features that most contributed to the aging clock’s output, our understanding of the underlying physiological mechanisms remains limited, particularly regarding why these features have a greater influence than others. A better understanding of the physiological insights behind these features, from a clinical perspective, could lead to more robust and interpretable aging clocks.

In conclusion, despite these limitations, our results demonstrate that physiological waveform-based aging clocks can predict an individual’s age and that the Age-Gap, defined as the difference between predicted and chronological age, is associated with CVD and may play a crucial role in the assessment of individual’s cardiovascular health. This study represents a significant step toward understanding human healthy aging through physiological waveforms.

## Methods

### Description of dataset

Our study uses a large perioperative dataset (MLORD) containing physiological waveforms from 17,327 patients who underwent surgeries at the David Geffen School of Medicine, University of California, Los Angeles (UCLA) between 2019 and 2022^73^. The MLORD dataset collection protocol was approved by the UCLA Institutional Review Board (IRB# 19– 000354; NIH research funding: R01HL14469). The dataset comprises both clinical and physiological waveform data. Clinical data were extracted from the electronic health record (EHR) platforms Epic (Verona, WI, USA) and Surgical Information Systems (Alpharetta, GA, USA), while the physiological waveform data were collected directly in the operating rooms using the Bernoulli data acquisition system (Cardiopulmonary, New Haven, CT, USA). The physiological waveform data includes more than 72,264 hours of recordings, approximately 7.6 TB in size, and contains digital waveforms such as ECG, ABP, PPG, and central venous pressure waveforms. A detailed description of the MLORD dataset can be found in Kim at el. ^73^. In this study, out of 17,327 patients, 16,663 with available PPG waveforms and 4,521 with available ABP waveforms were used for the analysis. Both PPG and ABP waveforms were sampled at 256 Hz.

Among 16,663 patients with PPG waveforms, 9,964 had CVD and 6,699 were non-CVD. Of 4,521 patients with ABP waveforms, 3,637 had CVD and 884 were non-CVD. The American Heart Association (AHA) definition was used as the ground truth for CVD classification. Patients were labelled as having CVD if they had ischemic heart disease, cerebrovascular disease, arrhythmia, valvular disease, or heart failure.

### PPG and ABP waveform feature extraction

To extract features from PPG and ABP waveforms, we used our recently developed feature extraction tool^50^, which operates in two stages. In the first stage, the tool identifies the temporal locations of key landmarks within each cardiac cycle, including the systolic phase onset, systolic phase peak, dicrotic notch, diastolic phase peak, and diastolic phase endpoint (see Fig. 1), using the Iterative Envelope Mean (IEM) method. The IEM method decomposes the waveform into stationary and non-stationary components, and the non-stationary component is subsequently used for landmark identification. In the second stage, it extracts more than 840 features per cardiac cycle based on these identified landmarks, including time-domain, frequency-domain, and statistical features.

We selected this tool for feature extraction from PPG and ABP waveforms for three main reasons:

1. Its ability to detect key landmarks within a cardiac cycle not only when they are clearly visible but also when they become less distinct. As discussed earlier, with aging, these landmarks, particularly the dicrotic notch, gradually diminish or disappear^7,56,80,81^. Accurate identification of these features is therefore essential, especially for reliable downstream feature extraction.
2. Its capability to extract a comprehensive set of more than 840 features per cardiac cycle from both PPG and ABP waveforms, encompassing time-domain, frequency-domain, and statistical characteristics.
3. To investigate the clinical applicability of the features extracted by our tool in the development of aging clocks.

In this study, the tool extracted 843 features per cardiac cycle from PPG waveforms and 846 features per cardiac cycle from ABP waveforms. These cardiac cycle features were then summarized at the patient level by calculating the mean and standard deviation of each feature across all cardiac cycles. This resulted in 843 averaged PPG features and 843 PPG standard deviation features, giving a total of 1,686 PPG features per patient. Similarly, 846 averaged ABP features and 846 ABP standard deviation features were obtained, resulting in a total of 1,692 ABP features per patient. Moreover, we refer readers to our previous article for the complete working process of the feature extraction tool and a detailed description of each extracted PPG and ABP feature ^50^.

### Aging clocks development

We developed two aging clocks: PPGAge and ABPAge based on PPG features and ABP features, respectively. For each clock, five ML models, LightGBM^66^, XGBoost^67^, Random Forest^68^, Lasso^69^, and Elastic net^70^, were trained on 70% of data and tested on the remaining held out 30% of data. The training and testing sets were as follows: for PPGAge, 11,664 patients were used for training, and the test set included 4,999 patients; for ABPAge, 3,164 patients were used for training, and the test set included 1,357 patients. Feature selection was performed on the training set using mutual information (MI)^100,101^ followed by an elbow detection method^102^, selecting 387 of 1,686 PPG features and 217 of 1,692 ABP features for model development (Fig. 10 (a), PPG features; Fig. 10 (b), ABP features). MI between each feature and chronological age was computed on the training set, and features were ranked by descending MI; to prevent overfitting, control computational complexity, and improve generalizability, we then retained only the top-ranked features (K) using geometric elbow detection, defined as the point of maximum perpendicular distance from the line connecting the first and last points of the sorted MI curve. Detailed descriptions of the selected 387 PPG features and 217 ABP features are provided in Supplementary Table 1. For each model, hyperparameters were tuned using randomized search with 5-fold cross-validation, performed within the training set (70% of the data) only; the held-out test set (30%) was reserved exclusively for final model evaluation and was not used during hyperparameter selection. The best-performing hyperparameter combination for each model, was fixed and used to refit the model on the full training set prior to evaluation on the test set. The selected hyperparameters for each model are presented in Table 5. All models were developed using the Python programming language (version 3.12.2). LightGBM and XGBoost were implemented using their respective standalone Python libraries, whereas the remaining models (Random Forest, Lasso, and Elastic Net) were implemented using the scikit-learn library.

**Table 5.**
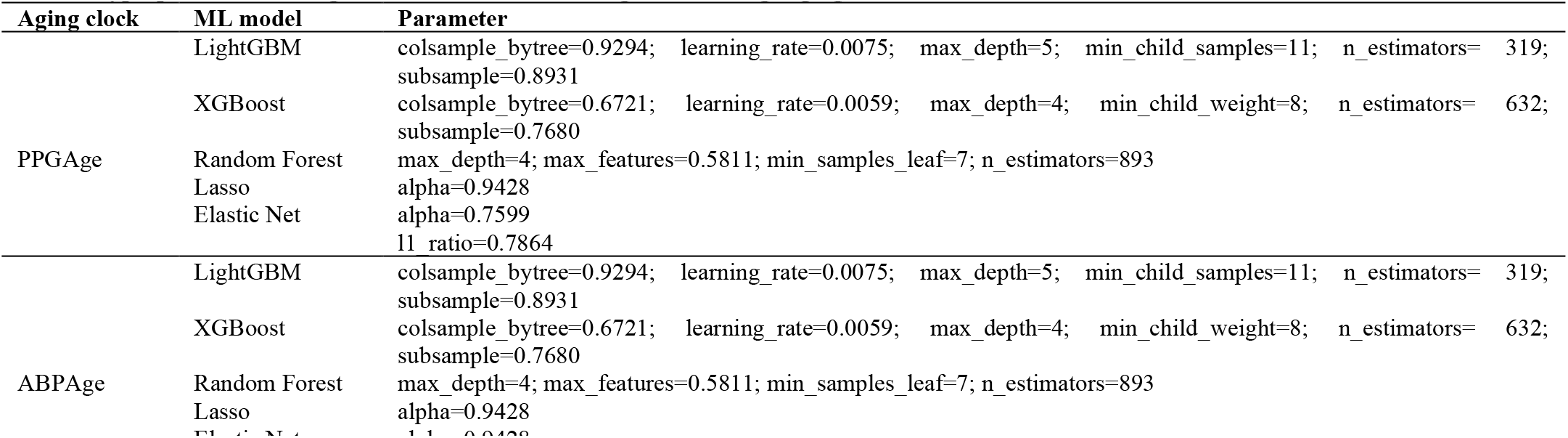
Hyperparameter settings for ML models for PPGAge and ABPAge aging clocks.

| Aging clock | ML model | Parameter |
|---|---|---|
| PPGAge | LightGBM | colsample_bytree=0.9294; learning_rate=0.0075; max_depth=5; min_child_samples=11; n_estimators= 319; subsample=0.8931 |
|  | XGBoost | colsample_bytree=0.6721; learning_rate=0.0059; max_depth=4; min_child_weight=8; n_estimators= 632; subsample=0.7680 |
|  | Random Forest | max_depth=4; max_features=0.5811; min_samples_leaf=7; n_estimators=893 |
|  | Lasso | alpha=0.9428 |
|  | Elastic Net | alpha=0.7599<br>l1_ratio=0.7864 |
| ABPAge | LightGBM | colsample_bytree=0.9294; learning_rate=0.0075; max_depth=5; min_child_samples=11; n_estimators= 319; subsample=0.8931 |
|  | XGBoost | colsample_bytree=0.6721; learning_rate=0.0059; max_depth=4; min_child_weight=8; n_estimators= 632; subsample=0.7680 |
|  | Random Forest | max_depth=4; max_features=0.5811; min_samples_leaf=7; n_estimators=893 |
|  | Lasso | alpha=0.9428 |
|  | [illegible] | [illegible] |

**Fig. 10.**
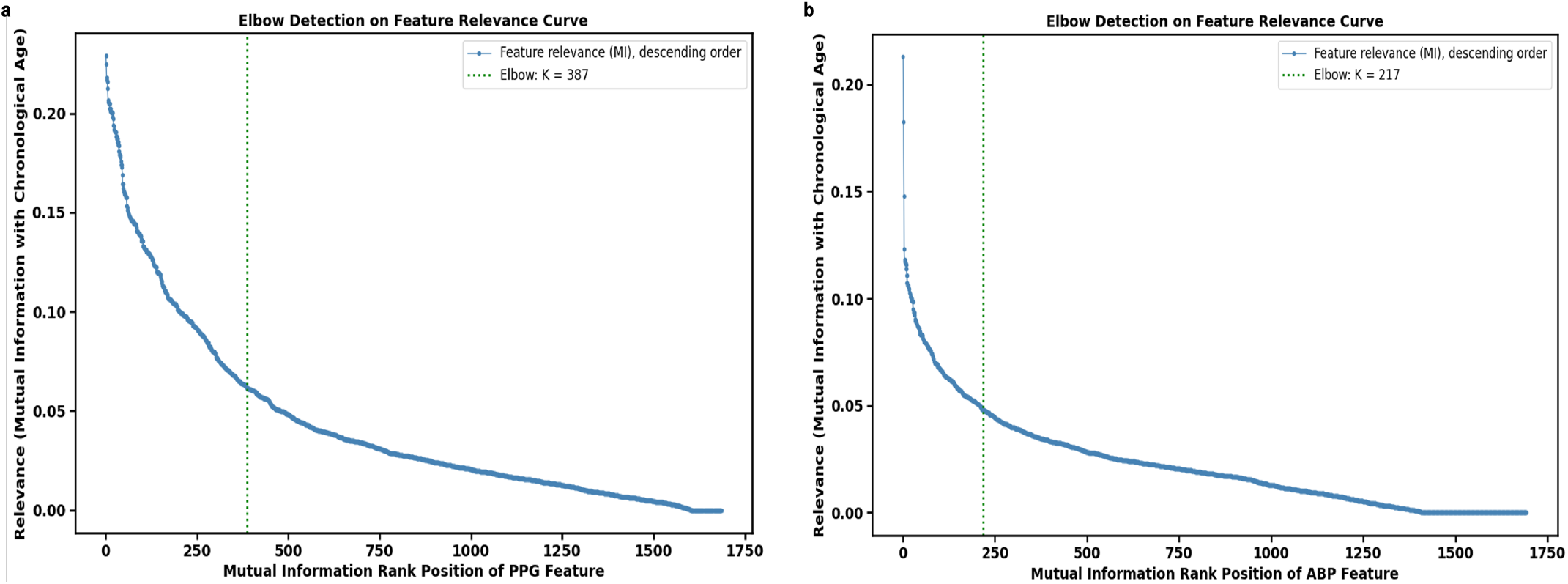
Feature selection by mutual information with elbow-based cutoff for PPG and ABP waveform features. **a** Photoplethysmography (PPG)-derived features were ranked from most to least informative based on their mutual information with chronological age (blue curve). An elbow detection method was used to select the optimal number of features (K = 387), identified as the point of maximum perpendicular distance from the line connecting the first and last points of the relevance curve (green dashed line), representing the point beyond which additional features contributed minimal relevance. **b** The same ranking and cutoff procedure was applied to arterial blood pressure (ABP)-derived features, selecting the top K = 217 features.

Predicted ages from the ML models using PPG features and ABP features are denoted as PPGAge and ABPAge, respectively, with the corresponding differences from chronological age referred to as the PPGAge Gap and ABPAge Gap. Many studies have observed that model predicted age is affected by age-related bias, with ages of younger individuals generally overestimated and ages of older individuals generally underestimated^13,103-107^. To account for this age dependence, we follow Miller et al.^13^, Tian et al.^104^, and Smith et al.^105^, by applying a second-step correction to the PPGAge Gap and ABPAge Gap. Specifically, as suggested by Miller et al.^13^ we regress the raw PPGAge Gap and ABPAge Gap on chronological age using a spline model, and use the residuals from this regression as the age-adjusted PPGAge Gap and ABPAge Gap, respectively.

### Quantitative evaluators

The performance of the both aging clocks PPGAge and ABPAge was evaluated using: Pearson correlation coefficient (*r*), coefficient of determination (*R*^*2*^), mean absolute error (MAE), root mean square error (RMSE)^74-76^, and Bland-Altman plots^108^. A Pearson correlation coefficient provides the linear association between chronological age and predicted age. Its value ranges from −1 to +1, where −1 represents a perfect negative correlation, 0 indicates no linear correlation, and +1 represents a perfect positive correlation^74^. The coefficient of determination measures how well the predicted ages match the chronological ages^75^. It quantifies the proportion of variance in the dependent variable (i.e., chronological age) that is explained by the model and provides an overall measure of model fit^76^. Its value ranges from 0 to 1, where 1 represents a perfect fit^75^. MAE provides the average magnitude of errors made by the model during age predictions. On the other hand, RMSE, by squaring the differences, penalizes larger errors more than MAE, making it more sensitive to outliers. The values of MAE and RMSE closer to 0 indicate more accurate predictions by the model^75,76^. The detailed equations for all four metrics are shown in Eqs. (1)–(4).

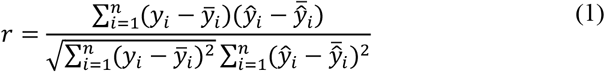

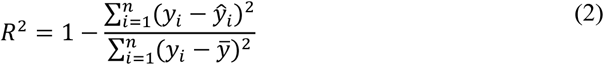

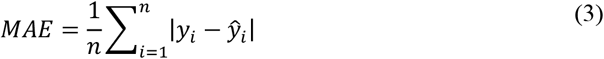

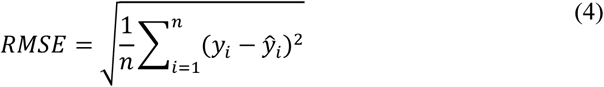

where *y* denotes chronological age, *ŷ* denotes predicted age, 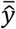 represents the mean chronological age, *ŷ*) represents the mean predicted age, and *n* denotes the overall sample size.

The Bland-Altman plots were used to evaluate the agreement between chronological age and predicted age.

### Interpretability analyses

We employed the SHapley Additive exPlanations (SHAP) method^71,72^ to the best-performing ML model for each aging clock to assess the contributions of individual PPG and ABP features to the PPGAge and ABPAge clocks, respectively. SHAP is a feature-based interpretability method that can be applied to ML models to better understand their predictions. The basic idea of SHAP is to assign a Shapley value to each feature, reflecting its contribution to the model’s output for a given data point^71,72^. We performed SHAP analysis using the Python SHAP package. The overall workflow for the development of aging clocks is illustrated in Fig. 11, where (a– e) and (a–b, f–h) show the pipelines for PPGAge aging clock and ABPAge aging clock, respectively.

**Fig. 11.**
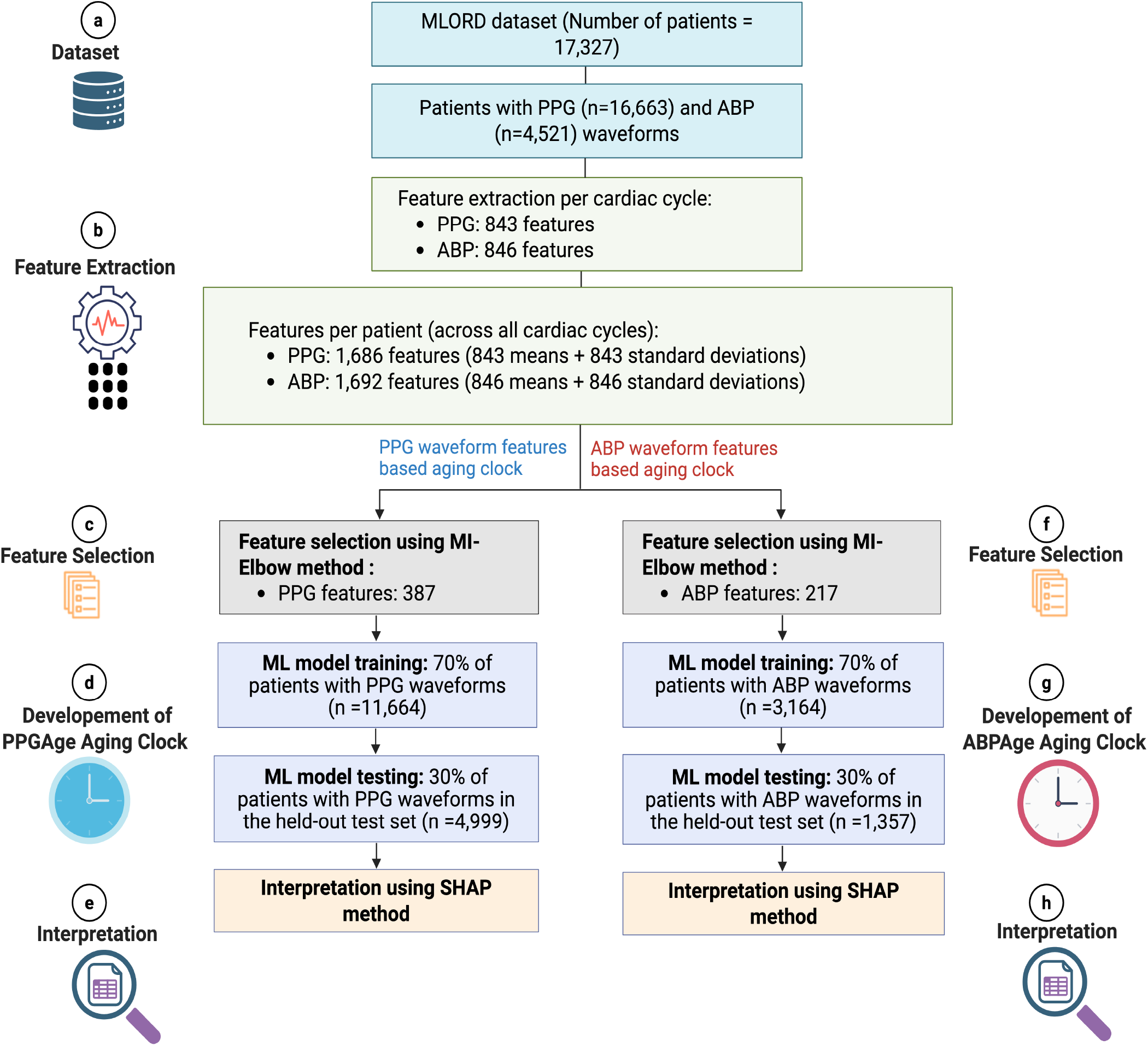
Overall workflow. **(a-h)**, Technical roadmap for the development of Machine leaning (ML) based aging clocks (PPGAge and ABPAge). Workflow includes: **a**, dataset, **b**, feature extraction, (**c, f)** feature selection for aging clocks, (**d, g)** development of aging clocks, **(f, i)**, SHAP interpretability.

### Definitions of Accelerated, Normal, and Delayed Aging Groups

For each aging clock, individuals were classified into three Age-Gap groups: accelerated aging, normal aging, and delayed aging, based on the Age-Gap (PPGAge Gap or ABPAge Gap), defined as the difference between the age predicted by the aging clock and the individual’s chronological age. A positive Age-Gap indicates that an individual is identified as older by the aging clock compared to their chronological age, whereas a negative Age-Gap reflects that an individual is recognized as younger by the aging clock compared to their chronological age. To define the Age-Gap groups, we followed the approach described in Ref. 10 and Ref. 109, using the model’s mean absolute error (MAE, rounded up to the nearest whole year) as the classification threshold. For the PPGAge and ABPAge aging clocks, the best-performing ML model (LightGBM) achieved held-out test-set MAEs of 5.90 and 5.83 years, respectively. Both MAEs rounded up to 6 years; therefore, a threshold of 6 years was applied to both aging clocks. Individuals with an Age-Gap ≥ +6 years were classified as having accelerated aging, whereas those with an Age-Gap < −6 years were classified as having delayed aging. Individuals with an Age-Gap within the threshold range were classified as having normal aging (−6 years ≤ Age-Gap < +6 years).

### Cardiovascular disease diagnosis rate

For each aging clock *c*, individuals were grouped by sex, *s* ∈ {male, female}, and within each sex, by chronological age group, *a* ∈ {[18, 50), [50, 90]}. The average CVD diagnosis rate^13^ was then calculated for each aging clock *c*, age group *a*, and sex *s* as (Eq. (5)):

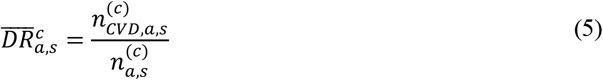

where 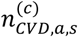 is the number of individuals diagnosed with CVD within the (*a, s*) group of clock *c*′*s* analysis cohort, and 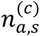 is the total number of individuals in that group.

Within each (*a, s*) group, individuals were further divided by their clock-specific Age-Gap group *q* (accelerated aging, normal aging, and delayed aging), and the CVD diagnosis rate was computed as (Eq. (6)):

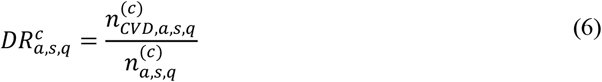

We then also computed the relative diagnosis rate (risk relative to average)^13^ within each group (*a, s, q*) using Eq. (7).

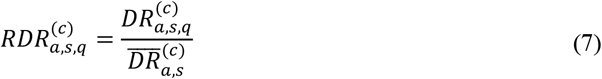

### Statistical analyses

To graphically illustrate the association between Age-Gap and CVD, polynomial smoothing splines curves were used^16^. To assess the association between aging groups and CVD, odds ratios (ORs) were calculated using logistic regression models, with predictors being the defined aging groups (“accelerated aging” or “delayed aging,” with “normal aging” as the reference category)^10^. The primary model was adjusted for chronological age and sex; sex-stratified models were adjusted for chronological age only. To assess the distribution of the PPG and ABP features for the PPGAge and ABPAge clocks, respectively, across aging groups, box plots were used, and pairwise comparisons between aging groups were assessed by Welch’s two-sided t-test. Statistical significance was defined as *p* < 0.05 for all analyses. All 95% confidence intervals reported in this study were estimated using a 1,000-iteration bootstrap procedure^110^. All statistical analyses were performed using the Python programming language (version 3.12.2).

## Data Availability

The interested parties may reach out to the first author at or the corresponding author at to request access to the MLORD dataset.

## Code availability

The code used in this study is not publicly available but may be provided to qualified researchers upon reasonable request to the first or corresponding author.

## Acknowledgements

This work was supported by the National Institutes of Health (NIH): R01EB029751 and R01HL144692.

## Author contributions

R.P. contributed to study design, data analysis, conceptualization, and manuscript preparation. A.B. contributed to study design, supervision of the study, review & editing of the manuscript, and conceptualization. M.C. contributed to study design, supervision of the study, review & editing of the manuscript, funding acquisition, and conceptualization. All authors have read and approved the final version of the manuscript.

