## Supplementary Table 1 for "Physiological Waveform Features Based Aging Clocks Associate with Cardiovascular Disease": Supplementary_material.pdf

### Detailed descriptions of the PPG features (n=387) used to develop the PPGAge aging clock

| PPG Features | Descriptions |
|---|---|
| PPG_D_RT_SYSPP_wrtDIAN_PPG_SYSPO_wrtSYSPP_mean | The ratio of the PPG cardiac cycle systolic phase peak duration with respect to dicrotic notch to the PPG systolic phase onset duration with respect to systolic phase peak, mean across cardiac cycles |
| PPG_D_RT_SYSPO_wrtDIAN_PPG_SYSPO_wrtSYSPP_mean | The ratio of the PPG cardiac cycle systolic phase onset duration with respect to dicrotic notch to the PPG cardiac cycle systolic phase onset duration with respect to systolic phase peak, mean across cardiac cycles |
| PPG_D_RT_NAre_wsDIAPE_SYSPP_wrtDIAN_PPG_D_NAre_wsDIAPE_SYSPO_wrtDIAPP_mean | The ratio of normalized area of systolic phase peak with respect to dicrotic notch, with subtracted diastolic phase end point to the area under the normalized PPG cardiac cycle systolic phase onset with respect to diastolic phase peak, with subtracted diastolic phase end point, mean across cardiac cycles |
| PPG_AM_SD_Bpeak_wrtDpeak_mean | PPG cardiac cycle second derivative Bpeak amplitude with respect to Dpeak, mean across cardiac cycles |
| PPG_D_RT_SYSPP_wrtDIAN_PPG_SYSPO_wrtDIAN_mean | The ratio of the PPG cardiac cycle systolic phase peak duration with respect to dicrotic notch to the PPG cardiac cycle systolic phase onset duration with respect to dicrotic notch, mean across cardiac cycles |
| PPG_D_RT_NAre_SYSPP_wrtDIAN_PPG_D_NAre_SYSPO_wrtDIAPP_mean | The ratio of the area under the normalized PPG cardiac cycle systolic phase peak with respect to dicrotic notch to the area under the normalized PPG cardiac cycle systolic phase onset with respect to diastolic phase peak, mean across cardiac cycles |
| PPG_D_RT_NAre_wsDIAPE_SYSPO_wrtDIAN_PPG_D_NAre_wsDIAPE_SYSPO_wrtSYSPP_mean | The ratio of the area under the normalized PPG cardiac cycle systolic phase onset with respect to dicrotic notch duration, with subtracted diastolic phase end point to the area under the normalized PPG cardiac cycle systolic phase onset with respect to systolic phase peak, with subtracted diastolic phase end point, mean across cardiac cycles |

| | |
|---|---|
| PPG_AM_RT_SD_Bpeak_wrtDpeak_PPG_AM_SD_Apeak_wrtzero_mean | The ratio of the PPG cardiac cycle second derivative Bpeak amplitude with respect to D peak to the PPG cardiac cycle second derivative Apeak amplitude with respect to zero, mean across cardiac cycles |
| PPG_D_RT_Are_SYSPP_wrtDIAN_PPG_D_Are_SYSPO_wrtDIAN_mean | The ratio of the area under the PPG cardiac cycle systolic phase peak with respect to dicrotic notch to the area under the PPG cardiac cycle systolic phase onset with respect to dicrotic notch, mean across cardiac cycles |
| PPG_D_RT_NAre_SYSPP_wrtDIAN_PPG_D_NAre_SYSPO_wrtDIAN_mean | The ratio of the area under the normalized PPG cardiac cycle systolic phase peak with respect to dicrotic notch to the area under the normalized PPG cardiac cycle systolic phase onset with respect to dicrotic notch, mean across cardiac cycles |
| PPG_D_RT_NAre_wsDIAPE_SYSPP_wrtDIAN_PPG_D_NAre_wsDIAPE_SYSPO_wrtDIAN_mean | The ratio of the area under the normalized PPG cardiac cycle systolic phase peak with respect to dicrotic notch, with subtracted diastolic phase end point to the area under the normalized PPG cardiac cycle systolic phase onset with respect to dicrotic notch, with subtracted diastolic phase end point, mean across cardiac cycles |
| PPG_D_RT_NAre_SYSPP_wrtDIAN_PPG_D_NAre_SYSPO_wrtSYSPP_mean | The ratio of the area under the normalized PPG cardiac cycle systolic phase peak with respect to dicrotic notch to the area under the normalized PPG cardiac cycle systolic phase onset with respect to systolic phase peak, mean across cardiac cycles |
| PPG_D_RT_NAre_SYSPO_wrtDIAN_PPG_D_NAre_SYSPO_wrtSYSPP_mean | The ratio of the area under the normalized PPG cardiac cycle systolic phase onset with respect to dicrotic notch to the area under the normalized PPG cardiac cycle systolic phase onset with respect to systolic phase peak, mean across cardiac cycles |
| PPG_D_RT_SYSPP_wrtDIAN_PPG_SYSPO_wrtDIAPP_mean | The ratio of the PPG cardiac cycle systolic phase peak duration with respect to dicrotic notch to the PPG cardiac cycle systolic phase onset duration with respect to diastolic phase peak, mean across cardiac cycles |
| PPG_W_SRP10_mean | PPG cardiac cycle systolic rise phase width at 10% of the cardiac cycle, mean across cardiac cycles |

| | |
|---|---|
| PPG_AM_SD_Bpeak_wrtCpeak_mean | PPG cardiac cycle second derivative Bpeak amplitude with respect to Cpeak, mean across cardiac cycles |
| PPG_D_RT_NAre_wsDIAPE_SYSPP_wrtDIAN_PPG_D_NAre_wsDIAPE_SYSPO_wrtSYSPP_mean | The ratio of the area under the normalized PPG cardiac cycle systolic phase peak with respect to dicrotic notch, with subtracted diastolic phase end point to the area under the normalized PPG cardiac cycle systolic phase onset with respect to systolic phase peak, with subtracted diastolic phase end point, mean across cardiac cycles |
| PPG_D_RT_Are_SYSPO_wrtDIAN_PPG_D_Are_SYSPO_wrtSYSPP_mean | The ratio of the area under the PPG cardiac cycle systolic phase onset with respect to dicrotic notch to the area under the PPG cardiac cycle systolic phase onset with respect to systolic phase peak, mean across cardiac cycles |
| PPG_D_RT_Are_SYSPP_wrtDIAN_PPG_D_Are_SYSPO_wrtSYSPP_mean | The ratio of the area under the PPG cardiac cycle systolic phase peak with respect to dicrotic notch to the area under the PPG cardiac cycle systolic phase onset with respect to systolic phase peak, mean across cardiac cycles |
| PPG_D_RT_Are_wsDIAPE_SYSPP_wrtDIAN_PPG_D_Are_wsDIAPE_SYSPO_wrtDIAN_mean | The ratio of the area under the PPG cardiac cycle systolic phase peak with respect to dicrotic notch, with subtracted diastolic phase end point to the area under the PPG cardiac cycle systolic phase onset with respect to dicrotic notch, with subtracted diastolic phase end point, mean across cardiac cycles |
| PPG_D_RT_Are_wsDIAPE_SYSPP_wrtDIAN_PPG_D_Are_wsDIAPE_SYSPO_wrtDIAN_mean | The ratio of the area under the PPG cardiac cycle systolic phase peak with respect to dicrotic notch, with subtracted diastolic phase end point to the area under the PPG cardiac cycle systolic phase onset with respect to diastolic phase peak, with subtracted diastolic phase end point, mean across cardiac cycles |
| PPG_D_RT_Are_wsDIAPE_SYSPP_wrtDIAN_PPG_D_Are_wsDIAPE_SYSPO_wrtSYSPP_mean | The ratio of the area under the PPG cardiac cycle systolic phase peak with respect to dicrotic notch, with subtracted diastolic phase end point to the area under the PPG cardiac cycle systolic phase onset with respect to systolic phase peak, with subtracted diastolic phase end point, mean across cardiac cycles |

| | |
|---|---|
| PPG_AM_SD_Bpeak_wrtzero_mean | PPG cardiac cycle second derivative Bpeak amplitude with respect to zero, mean across cardiac cycles |
| PPG_D_NAre_SYSPO_wrtSYSPP_mean | Area under the normalized PPG cardiac cycle systolic phase onset with respect to systolic phase peak, mean across cardiac cycles |
| PPG_AM_RT_SD_Bpeak_wrtzero_PPG_AM_SD_Apeak_wrtzero_mean | The ratio of the PPG cardiac cycle second derivative Bpeak amplitude with respect to zero to the PPG cardiac cycle second derivative Apeak amplitude with respect to zero, mean across cardiac cycles |
| PPG_AM_RT_SD_Apeak_wrtBpeak_PPG_AM_SD_Apeak_wrtzero_mean | The ratio of the APPG cardiac cycle second derivative Apeak amplitude with respect to Bpeak to the PPG cardiac cycle second derivative Apeak amplitude with respect to zero, mean across cardiac cycles |
| PPG_AM_RT_SD_Apeak_wrtDpeak_PPG_AM_SD_Bpeak_wrtzero_mean | The ratio of the PPG cardiac cycle second derivative Apeak amplitude with respect to Dpeak to the PPG cardiac cycle second derivative Bpeak amplitude with respect to zero, mean across cardiac cycles |
| PPG_D_RT_Are_wsDIAPE_SYSPO_wrtDIAN_PPG_D_Are_wsDIAPE_SYSPO_wrtSYSPP_mean | The ratio of the area under the PPG cardiac cycle systolic phase onset with respect to dicrotic notch, with subtracted diastolic phase end point to the area under the PPG cardiac cycle systolic phase onset with respect to systolic phase peak, with subtracted diastolic phase end point, mean across cardiac cycles |
| PPG_D_RT_Are_SYSPP_wrtDIAN_PPG_D_Are_SYSPO_wrtDIAPP_mean | The ratio of the area under the PPG cardiac cycle systolic phase peak with respect to dicrotic notch to the area under the PPG cardiac cycle systolic phase onset with respect to diastolic phase peak, mean across cardiac cycles |
| PPG_AM_RT_SD_Bpeak_wrtDpeak_PPG_AM_SD_Apeak_wrtEpeak_mean | The ratio of the PPG cardiac cycle second derivative Bpeak amplitude with respect to Dpeak to the PPG cardiac cycle second derivative Apeak amplitude with respect to Epeak, mean across cardiac cycles |
| PPG_AM_RT_SD_Bpeak_wrtDpeak_PPG_AM_SD_Apeak_wrtBpeak_mean | The ratio of the PPG cardiac cycle second derivative Bpeak amplitude with respect to Dpeak to the PPG cardiac cycle second derivative Apeak amplitude with respect to Bpeak , mean across cardiac cycles |

| | |
|---|---|
| PPG_AM_RT_SD_Apeak_wrtDpeak_PPG_AM_SD_Apeak_wrtBpeak_mean | The ratio of the PPG cardiac cycle second derivative Apeak amplitude with respect to Dpeak to the PPG cardiac cycle second derivative Apeak amplitude with respect to Bpeak, mean across cardiac cycles |
| PPG_D_NAre_wsDIAPE_SYSPO_wrtSYSPP_mean | Area under the normalized PPG cardiac cycle systolic phase onset with respect to systolic phase peak, with subtracted diastolic phase end point, mean across cardiac cycles |
| PPG_D_Are_SYSPO_wrtSYSPP_mean | Area under the PPG cardiac cycle systolic phase onset with respect to systolic phase peak, mean across cardiac cycles |
| PPG_D_Are_wsDIAPE_SYSPO_wrtSYSPP_mean | Area under the PPG cardiac cycle systolic phase onset with respect to systolic phase peak, with subtracted diastolic phase end point, mean across cardiac cycles |
| PPG_W_SRP50_mean | PPG cardiac cycle systolic rise phase width at 50% of the cardiac cycle, mean across cardiac cycles |
| PPG_W_SRP33_mean | PPG cardiac cycle systolic rise phase width at 33% of the cardiac cycle, mean across cardiac cycles |
| PPG_AM_RT_SD_Apeak_wrtBpeak_PPG_AM_SD_Bpeak_wrtzero_mean | The ratio of the PPG cardiac cycle second derivative Apeak amplitude with respect to Bpeak to the PPG cardiac cycle second derivative Bpeak amplitude with respect to zero, mean across cardiac cycles |
| PPG_AM_RT_SD_Bpeak_wrtDpeak_PPG_AM_SD_Apeak_wrtDpeak_mean | The ratio of the PPG cardiac cycle second derivative Bpeak amplitude with respect to Dpeak to the PPG cardiac cycle second derivative Apeak amplitude with respect to Dpeak, mean across cardiac cycles |
| PPG_AM_RT_SD_Dpeak_wrtEpeak_PPG_AM_SD_Bpeak_wrtCpeak_mean | The ratio of the PPG cardiac cycle second derivative Dpeak amplitude with respect to Epeak to the PPG cardiac cycle second derivative Bpeak amplitude with respect to Cpeak, mean across cardiac cycles |
| PPG_Slope_SYSPO_SYSPP_mean | Mean across cardiac cycles of the slope between the systolic phase onset and systolic phase peak of the PPG cardiac cycle |
| PPG_W_SRP25_mean | PPG cardiac cycle systolic rise phase width at 25% of the cardiac cycle, mean across cardiac cycles |

| | |
|---|---|
| PPG_AM_RT_SD_Bpeak_wrtDpeak_PPG_AM_SD_Bpeak_wrtzero_mean | The ratio of the PPG cardiac cycle second derivative Bpeak amplitude with respect to Dpeak to the PPG cardiac cycle second derivative Bpeak amplitude with respect to zero, mean across cardiac cycles |
| PPG_AM_RT_SD_Dpeak_wrtzero_PPG_AM_SD_Bpeak_wrtzero_mean | The ratio of the PPG cardiac cycle second derivative Dpeak amplitude with respect to zero to the PPG cardiac cycle second derivative Bpeak amplitude with respect to zero, mean across cardiac cycles |
| PPG_D_SYSPO_wrtSYSPP_mean | Duration of the PPG cardiac cycle systolic phase onset with respect to systolic phase peak, mean across cardiac cycles |
| PPG_AM_SD_Bpeak_wrtEpeak_mean | PPG cardiac cycle second derivative Bpeak amplitude with respect to Epeak, mean across cardiac cycles |
| PPG_D_RT_NAre_SYSPP_wrtDIAN_PPG_D_NAre_SYSPO_wrtDIAPE_mean | The ratio of the area under the normalized PPG cardiac cycle systolic phase peak with respect to dicrotic notch to the area under the normalized PPG cardiac cycle systolic phase onset with respect to diastolic phase end point, mean across cardiac cycles |
| PPG_W_SRP66_mean | PPG cardiac cycle systolic rise phase width at 66% of the cardiac cycle<br>, mean across cardiac cycles |
| PPG_AM_SD_Apeak_wrtBpeak_mean | PPG cardiac cycle second derivative Apeak amplitude with respect to Bpeak, mean across cardiac cycles |
| PPG_D_RT_SYSPP_wrtDIAPP_PPG_SYSPO_wrtDIAPP_mean | The ratio of the PPG cardiac cycle systolic phase peak duration with respect to diastolic phase peak to the PPG cardiac cycle systolic phase onset duration with respect to diastolic phase peak, mean across cardiac cycles |
| PPG_D_RT_NAre_wsDIAPE_SYSPP_wrtDIAN_PPG_D_NAre_wsDIAPE_SYSPO_wrtDIAPE_mean | The ratio of normalized area of systolic phase peak with respect to dicrotic notch, with subtracted diastolic phase end point to the area under the normalized PPG cardiac cycle systolic phase onset with respect to diastolic phase end point, with subtracted diastolic phase end point, mean across cardiac cycles |
| PPG_D_RT_SYSPP_wrtDIAPP_PPG_SYSPO_wrtSYSPP_mean | The ratio of the PPG cardiac cycle systolic phase peak duration with respect to diastolic phase peak to the PPG cardiac cycle systolic phase |

| | |
|---|---|
| | onset duration with respect to systolic phase peak, mean across cardiac cycles |
| PPG_VAge1_mean | PPG cardiac cycle second derivative feature defined as: (Bpeak − Cpeak − Dpeak − Epeak)/Apeak, with all peak amplitudes measured relative to the zero baseline and the ratio averaged across cardiac cycles |
| PPG_AM_RT_SD_Bpeak_wrtDpeak_PPG_AM_SD_Apeak_wrtCpeak_mean | The ratio of the PPG cardiac cycle second derivative Bpeak amplitude with respect to Dpeak to the PPG cardiac cycle second derivative Apeak amplitude with respect to Cpeak, mean across cardiac cycles |
| PPG_D_RT_SYSPO_wrtDIAPP_PPG_SYSPO_wrtSYSPP_mean | The ratio of the PPG cardiac cycle systolic phase onset duration with respect to diastolic phase peak to the PPG cardiac cycle systolic phase onset duration with respect to systolic phase peak, mean across cardiac cycles |
| PPG_AM_RT_SD_Apeak_wrtCpeak_PPG_AM_SD_Bpeak_wrtzero_mean | The ratio of the PPG cardiac cycle second derivative Apeak amplitude with respect to Cpeak to the PPG cardiac cycle second derivative Bpeak amplitude with respect to zero, mean across cardiac cycles |
| PPG_AM_RT_SD_Bpeak_wrtEpeak_PPG_AM_SD_Apeak_wrtDpeak_mean | The ratio of the PPG cardiac cycle second derivative Bpeak amplitude with respect to Epeak to the PPG cardiac cycle second derivative Apeak amplitude with respect to Dpeak, mean across cardiac cycles |
| PPG_D_RT_Are_wsDIAPE_SYSPO_wrtDIAPP_PPG_D_Are_wsDIAPE_SYSPO_wrtSYSPP_mean | The ratio of the area under the PPG cardiac cycle systolic phase onset with respect to diastolic phase peak, with subtracted diastolic phase end point to the area under the PPG cardiac cycle systolic phase onset with respect to systolic phase peak, with subtracted diastolic phase end point, mean across cardiac cycles |
| PPG_NAM_RT_DIAN_wrtZero_PPG_SYSPP_wrtSYSPO_mean | The ratio of the normalized PPG cardiac cycle dicrotic notch with respect to zero to the normalized PPG cardiac cycle systolic phase peak amplitude with respect to systolic phase onset, mean across cardiac cycles |
| PPG_D_RT_NAre_wsDIAPE_SYSPP_wrtDIAPP_PPG_D_NAre_wsDIAPE_SYSPO_wrtDIAPP_mean | The ratio of the area under the normalized PPG cardiac cycle systolic phase peak with respect to diastolic phase peak, with subtracted diastolic phase end point to the area under the normalized PPG cardiac cycle systolic phase |

| | |
|---|---|
| | onset with respect to diastolic phase peak, with subtracted diastolic phase end point, mean across cardiac cycles |
| PPG_D_RT_Are_wsDIAPE_SYSPP_wrtDIAPP_PPG_D_Are_wsDIAPE_SYSPO_wrtSYSPP_mean | The ratio of the area under the PPG cardiac cycle systolic phase peak with respect to diastolic phase peak, with subtracted diastolic phase end point to the area under the PPG cardiac cycle systolic phase onset with respect to systolic phase peak, with subtracted diastolic phase end point, mean across cardiac cycles |
| PPG_D_RT_NAre_DIAN_wrtDIAPP_PPG_D_NAre_SYSPP_wrtDIAPP_mean | The ratio of the area under the normalized PPG cardiac cycle dicrotic notch with respect to diastolic phase peak to the area under the normalized PPG cardiac cycle systolic phase peak with respect to diastolic phase peak, mean across cardiac cycles |
| PPG_W_SRP75_mean | PPG cardiac cycle systolic rise phase width at 75% of the cardiac cycle, mean across cardiac cycles |
| PPG_D_RT_NAre_SYSPP_wrtDIAPP_PPG_D_NAre_SYSPO_wrtDIAPP_mean | The ratio of the area under the normalized PPG cardiac cycle systolic phase peak with respect to diastolic phase peak to the area under the normalized PPG cardiac cycle systolic phase onset with respect to diastolic phase peak, mean across cardiac cycles |
| PPG_NAM_SYSPP_wrtDIAN_mean | Normalized PPG cardiac cycle systolic phase peak amplitude with respect to dicrotic notch, mean across cardiac cycles |
| PPG_AM_RT_SD_Bpeak_wrtEpeak_PPG_AM_SD_Apeak_wrtCpeak_mean | The ratio of the PPG cardiac cycle second derivative Bpeak amplitude with respect to Epeak to the PPG cardiac cycle second derivative Apeak amplitude with respect to Cpeak, mean across cardiac cycles |
| PPG_D_RT_NAre_DIAN_wrtDIAPP_PPG_D_NAre_SYSPP_wrtDIAN_mean | The ratio of the area under the normalized PPG cardiac cycle dicrotic notch with respect to diastolic phase peak to the area under the normalized PPG cardiac cycle systolic phase peak with respect to dicrotic notch, mean across cardiac cycles |
| PPG_D_RT_NAre_SYSPP_wrtDIAPP_PPG_D_NAre_SYSPP_wrtDIAN_mean | The ratio of the area under the normalized PPG cardiac cycle systolic phase peak with respect to diastolic phase peak to the area under the normalized PPG cardiac cycle systolic phase |

| | |
|---|---|
| | peak with respect to dicrotic notch, mean across cardiac cycles |
| PPG_D_RT_Are_DIAN_wrtDIAPP_PPG_D_Are_SYSPP_wrtDIAPP_mean | The ratio of the area under the PPG cardiac cycle dicrotic notch with respect to diastolic phase peak to the area under the PPG cardiac cycle systolic phase peak with respect to diastolic phase peak, mean across cardiac cycles |
| PPG_D_RT_Are_SYSPP_wrtDIAPP_PPG_D_Are_SYSPO_wrtSYSPP_mean | The ratio of the area under the PPG cardiac cycle systolic phase peak with respect to diastolic phase peak to the area under the PPG cardiac cycle systolic phase onset with respect to systolic phase peak, mean across cardiac cycles |
| PPG_D_RT_Are_SYSPO_wrtDIAPP_PPG_D_Are_SYSPO_wrtSYSPP_mean | The ratio of the area under the PPG cardiac cycle systolic phase onset with respect to diastolic phase peak to the area under the PPG cardiac cycle systolic phase onset with respect to systolic phase peak, mean across cardiac cycles |
| PPG_D_RT_NAre_wsDIAPE_SYSPP_wrtDIAPP_PPG_D_NAre_wsDIAPE_SYSPO_wrtSYSPP_mean | The ratio of the area under the normalized APPG cardiac cycle systolic phase peak with respect to diastolic phase peak, with subtracted diastolic phase end point to the area under the normalized PPG cardiac cycle systolic phase onset with respect to systolic phase peak, with subtracted diastolic phase end point, mean across cardiac cycles |
| PPG_D_RT_NAre_wsDIAPE_DIAN_wrtDIAPP_PPG_D_NAre_wsDIAPE_SYSPP_wrtDIAN_mean | The ratio of the area under the normalized PPG cardiac cycle dicrotic notch with respect to diastolic phase peak, with subtracted diastolic phase end point to the area under the normalized PPG cardiac cycle systolic phase peak with respect to dicrotic notch, with subtracted diastolic phase end point, mean across cardiac cycles |
| PPG_D_RT_Are_wsDIAPE_SYSPP_wrtDIAPP_PPG_D_Are_wsDIAPE_SYSPP_wrtDIAN_mean | The ratio of the area under the PPG cardiac cycle systolic phase peak with respect to diastolic phase peak, with subtracted diastolic phase end point to the area under the PPG cardiac cycle systolic phase peak with respect to dicrotic notch, with subtracted diastolic phase end point, mean across cardiac cycles |

| | |
|---|---|
| PPG_D_RT_NAre_SYSPO_wrtDIAPP_PPG_D_NAre_SYSPO_wrtSYSPP_mean | The ratio of the area under the normalized PPG cardiac cycle systolic phase onset with respect to diastolic phase peak to the area under the normalized PPG cardiac cycle systolic phase onset with respect to systolic phase peak, mean across cardiac cycles |
| PPG_D_RT_NAre_SYSPP_wrtDIAPP_PPG_D_NAre_SYSPO_wrtSYSPP_mean | The ratio of the area under the normalized PPG cardiac cycle systolic phase peak with respect to diastolic phase peak to the area under the normalized PPG cardiac cycle systolic phase onset with respect to systolic phase peak, mean across cardiac cycles |
| PPG_D_RT_NAre_wsDIAPE_SYSPO_wrtDIAPP_PPG_D_NAre_wsDIAPE_SYSPO_wrtSYSPP_mean | The ratio of the area under the normalized PPG cardiac cycle systolic phase onset with respect to diastolic phase peak, with subtracted diastolic phase end point to the area under the normalized PPG cardiac cycle systolic phase onset with respect to systolic phase peak, with subtracted diastolic phase end point, mean across cardiac cycles |
| PPG_D_RT_NAre_wsDIAPE_SYSPP_wrtDIAPP_PPG_D_NAre_wsDIAPE_SYSPP_wrtDIAN_mean | The ratio of the area under the normalized PPG cardiac cycle systolic phase peak with respect to diastolic phase peak, with subtracted diastolic phase end point to the area under the normalized PPG cardiac cycle systolic phase peak with respect to dicrotic notch, with subtracted diastolic phase end point, mean across cardiac cycles |
| PPG_AM_RT_SD_Bpeak_wrtCpeak_PPG_AM_SD_Apeak_wrtDpeak_mean | The ratio of the PPG cardiac cycle second derivative Bpeak amplitude with respect to Cpeak to the PPG cardiac cycle second derivative Apeak amplitude with respect to Dpeak, mean across cardiac cycles |
| PPG_D_RT_SYSPP_wrtDIAN_PPG_SYSPO_wrtDIAPE_mean | The ratio of the PPG cardiac cycle systolic phase peak duration with respect to dicrotic notch to the PPG cardiac cycle systolic phase onset duration with respect to diastolic phase end point, mean across cardiac cycles |
| PPG_D_RT_Are_SYSPP_wrtDIAPP_PPG_D_Are_SYSPP_wrtDIAN_mean | The ratio of the area under the PPG cardiac cycle systolic phase peak with respect to diastolic phase peak to the area under the PPG cardiac cycle systolic phase peak with respect to dicrotic notch, mean across cardiac cycles |

| | |
|---|---|
| PPG_D_RT_Are_DIAN_wrtDIAPP_PPG_D_Are_SYSPP_wrtDIAN_mean | The ratio of the area under the PPG cardiac cycle dicrotic notch with respect to diastolic phase peak to the area under the PPG cardiac cycle systolic phase peak with respect to dicrotic notch, mean across cardiac cycles |
| PPG_D_RT_NAre_wsDIAPE_DIAN_wrtDIAPP_PPG_D_NAre_wsDIAPE_SYSPP_wrtDIAPP_mean | The ratio of the area under the normalized PPG cardiac cycle dicrotic notch with respect to diastolic phase peak, with subtracted diastolic phase end point to the area under the normalized PPG cardiac cycle systolic phase peak with respect to diastolic phase peak, with subtracted diastolic phase end point, mean across cardiac cycles |
| PPG_D_RT_Are_wsDIAPE_DIAN_wrtDIAPP_PPG_D_Are_wsDIAPE_SYSPP_wrtDIAN_mean | The ratio of the area under the PPG cardiac cycle dicrotic notch with respect to diastolic phase end peak, with subtracted diastolic phase end point to the area under the PPG cardiac cycle systolic phase peak with respect to dicrotic notch, with subtracted diastolic phase end point, mean across cardiac cycles |
| PPG_AM_RT_SD_Bpeak_wrtDpeak_PPG_AM_SD_Bpeak_wrtCpeak_mean | The ratio of the PPG cardiac cycle second derivative Bpeak amplitude with respect to Dpeak to the PPG cardiac cycle second derivative Bpeak amplitude with respect to Cpeak, mean across cardiac cycles |
| PPG_AM_RT_SD_Cpeak_wrtDpeak_PPG_AM_SD_Bpeak_wrtCpeak_mean | The ratio of the PPG cardiac cycle second derivative Cpeak amplitude with respect to Dpeak to the PPG cardiac cycle second derivative Bpeak amplitude with respect to Cpeak, mean across cardiac cycles |
| PPG_AM_RT_SD_Dpeak_wrtEpeak_PPG_AM_SD_Bpeak_wrtEpeak_mean | The ratio of the APPG cardiac cycle second derivative Dpeak amplitude with respect to Epeak to the PPG cardiac cycle second derivative Bpeak amplitude with respect to Epeak, mean across cardiac cycles |
| PPG_AM_RT_SD_Dpeak_wrtzero_PPG_AM_SD_Apeak_wrtzero_mean | The ratio of the PPG cardiac cycle second derivative Dpeak amplitude with respect to zero to the PPG cardiac cycle second derivative Apeak amplitude with respect to zero, mean across cardiac cycles |
| PPG_AM_RT_SD_Apeak_wrtDpeak_PPG_AM_SD_Apeak_wrtzero_mean | The ratio of the PPG cardiac cycle second derivative Apeak amplitude with respect to Dpeak to the PPG cardiac cycle second |

| | |
|---|---|
| | derivative Apeak amplitude with respect to zero, mean across cardiac cycles |
| PPG_AM_RT_SD_Bpeak_wrtCpeak_PPG_AM_SD_Apeak_wrtzero_mean | The ratio of the PPG cardiac cycle second derivative Bpeak amplitude with respect to Cpeak to the PPG cardiac cycle second derivative Apeak amplitude with respect to zero, mean across cardiac cycles |
| PPG_AM_RT_SD_Bpeak_wrtEpeak_PPG_AM_SD_Apeak_wrtzero_mean | The ratio of the PPG cardiac cycle second derivative Bpeak amplitude with respect to Epeak to the PPG cardiac cycle second derivative Apeak amplitude with respect to zero, mean across cardiac cycles |
| PPG_Vage2_mean | PPG cardiac cycle second derivative feature defined as: (Bpeak − Epeak)/Apeak, with all peak amplitudes measured relative to the zero baseline and the ratio averaged across cardiac cycles |
| PPG_AM_RT_SD_Apeak_wrtEpeak_PPG_AM_SD_Bpeak_wrtzero_mean | The ratio of the PPG cardiac cycle second derivative Apeak amplitude with respect to Epeak to The ratio of the PPG cardiac cycle second derivative Bpeak amplitude with respect to zero, mean across cardiac cycles |
| PPG_NAM_RT_DIAN_wrtZero_PPG_SYSPP_wrtDIAPE_mean | The ratio of the normalized PPG cardiac cycle dicrotic notch with respect to zero to the normalized PPG cardiac cycle systolic phase peak amplitude with respect to diastolic phase end point, mean across cardiac cycles |
| PPG_AM_RT_DIAN_wrtSYSPO_PPG_SYSPP_wrtDIAPE_mean | The ratio of thePPG cardiac cycle dicrotic notch amplitude with respect to systolic phase onset to the PPG cardiac cycle systolic phase peak amplitude with respect to diastolic phase end point, mean across cardiac cycles |
| PPG_AM_DIAN_wrtZero_mean | PPG cardiac cycle dicrotic notch amplitude with respect to zero, mean across cardiac cycles |
| PPG_NAM_RT_DIAN_wrtSYSPO_PPG_SYSPP_wrtDIAPE_mean | The ratio of the normalized PPG cardiac cycle dicrotic notch with respect to systolic phase onset to the normalized PPG cardiac cycle systolic phase peak amplitude with respect to diastolic phase end point, mean across cardiac cycles |
| PPG_D_RT_Are_wsDIAPE_SYSPP_wrtDIAPP_PPG_D_Are_wsDIAPE_SYSPO_wrtDIAPP_mean | The ratio of the area under the PPG cardiac cycle systolic phase peak with respect to diastolic phase peak, with subtracted diastolic phase end point to the area under the PPG |

| | |
|---|---|
| | cardiac cycle systolic phase onset with respect to diastolic phase peak, with subtracted diastolic phase end point, mean across cardiac cycles |
| PPG_D_RT_SD_Cpeak_wrtDpeak_PPG_D_SD_Bpeak_wrtCpeak_mean | The ratio of the PPG cardiac cycle second derivative Cpeak duration with respect to Dpeak to the PPG cardiac cycle second derivative Bpeak duration with respect to Cpeak, mean across cardiac cycles |
| PPG_D_RT_Are_SYSPP_wrtDIAPP_PPG_D_Are_SYSPO_wrtDIAPP_mean | The ratio of the area under the PPG cardiac cycle systolic phase peak with respect to diastolic phase peak to the area under the PPG cardiac cycle systolic phase onset with respect to diastolic phase peak, mean across cardiac cycles |
| PPG_AM_RT_SD_Bpeak_wrtEpeak_PPG_AM_SD_Bpeak_wrtCpeak_mean | The ratio of the PPG cardiac cycle second derivative Bpeak amplitude with respect to Epeak to the PPG cardiac cycle second derivative Bpeak amplitude with respect to Cpeak, mean across cardiac cycles |
| PPG_AM_RT_SD_Cpeak_wrtEpeak_PPG_AM_SD_Bpeak_wrtCpeak_mean | The ratio of the PPG cardiac cycle second derivative Cpeak amplitude with respect to Epeak to the PPG cardiac cycle second derivative Bpeak amplitude with respect to Cpeak, mean across cardiac cycles |
| PPG_D_RT_Are_wsDIAPE_SYSPP_wrtDIAN_PPG_D_Are_wsDIAPE_SYSPO_wrtDIAPE_mean | The ratio of the area under the PPG cardiac cycle systolic phase peak with respect to dicrotic notch, with subtracted diastolic phase end point to the area under the PPG cardiac cycle systolic phase onset with respect to diastolic phase end point, with subtracted diastolic phase end point, mean across cardiac cycles |
| PPG_NAM_DIAN_wrtZero_mean | Normalized PPG cardiac cycle dicrotic notch amplitude with respect to zero, mean across cardiac cycles |
| PPG_D_RT_SD_Cpeak_wrtDpeak_PPG_D_SD_Bpeak_wrtCpeak_std | The ratio of the PPG cardiac cycle second derivative Cpeak duration with respect to Dpeak to the PPG cardiac cycle second derivative Bpeak duration with respect to Cpeak, standard deviation across cardiac cycles |
| PPG_D_NAre_DIAN_wrtDIAPP_mean | Area under the normalized PPG cardiac cycle dicrotic notch with respect to diastolic phase peak, mean across cardiac cycles |

| | |
|---|---|
| PPG_AM_RT_SD_Bpeak_wrtCpeak_PPG_AM_SD_Apeak_wrtCpeak_mean | The ratio of the PPG cardiac cycle second derivative Bpeak amplitude with respect to Cpeak to the PPG cardiac cycle second derivative Apeak amplitude with respect to Cpeak, mean across cardiac cycles |
| PPG_D_RT_Are_wsDIAPE_DIAN_wrtDIAPP_PPG_D_Are_wsDIAPE_SYSPP_wrtDIAPP_mean | The ratio of the area under the PPG cardiac cycle dicrotic notch with respect to diastolic phase peak, with subtracted diastolic phase end point to the area under the PPG cardiac cycle systolic phase peak with respect to diastolic phase peak, with subtracted diastolic phase end point, mean across cardiac cycles |
| PPG_D_RT_DIAN_wrtDIAPP_PPG_SYSPP_wrtDIAN_mean | The ratio of the PPG cardiac cycle dicrotic notch duration with respect to diastolic phase peak to the PPG cardiac cycle systolic phase peak duration with respect to dicrotic notch, mean across cardiac cycles |
| PPG_AM_RT_SD_Bpeak_wrtEpeak_PPG_AM_SD_Bpeak_wrtDpeak_std | The ratio of the PPG cardiac cycle second derivative Bpeak amplitude with respect to Epeak to the PPG cardiac cycle second derivative Bpeak amplitude with respect to Dpeak, standard deviation across cardiac cycles |
| PPG_AM_RT_SD_Dpeak_wrtEpeak_PPG_AM_SD_Bpeak_wrtDpeak_std | The ratio of the PPG cardiac cycle second derivative Dpeak amplitude with respect to Epeak to the PPG cardiac cycle second derivative Bpeak amplitude with respect to Dpeak, standard deviation across cardiac cycles |
| PPG_D_RT_SD_Bpeak_wrtDpeak_PPG_D_SD_Bpeak_wrtCpeak_mean | The ratio of the PPG cardiac cycle second derivative Bpeak duration with respect to Dpeak to the PPG cardiac cycle second derivative Bpeak duration with respect to Cpeak, mean across cardiac cycles |
| PPG_NAM_RT_DIAN_wrtZero_PPG_SYSPP_wrtZero_mean | The ratio of the normalized PPG cardiac cycle dicrotic notch with respect to zero to the normalized PPG cardiac cycle systolic phase peak amplitude with respect to zero, mean across cardiac cycles |
| PPG_NAM_RT_SYSPP_wrtDIAN_PPG_SYSPP_wrtZero_mean | The ratio of the normalized PPG cardiac cycle systolic phase peak amplitude with respect to dicrotic notch to the normalized PPG cardiac cycle systolic phase peak amplitude with respect to zero, mean across cardiac cycles |
| PPG_AM_RT_DIAPP_wrtDIAN_PPG_SYSPP_wrtDIAN_mean | The ratio of the PPG cardiac cycle diastolic phase peak amplitude with respect to dicrotic |

| | |
|---|---|
| | notch to the PPG cardiac cycle systolic phase peak amplitude with respect to dicrotic notch, mean across cardiac cycles |
| PPG_AM_RT_SYSPP_wrtDIAPP_PPG_SYSPP_wrtDIAN_mean | The ratio of the PPG cardiac cycle systolic phase peak amplitude with respect to diastolic phase peak to the PPG cardiac cycle systolic phase peak amplitude with respect to dicrotic notch, mean across cardiac cycles |
| PPG_AM_RT_SD_Bpeak_wrtEpeak_PPG_AM_SD_Dpeak_wrtzero_mean | The ratio of the PPG cardiac cycle second derivative Bpeak amplitude with respect to Epeak to the PPG cardiac cycle second derivative Dpeak amplitude with respect to zero, mean across cardiac cycles |
| PPG_AM_RT_SYSPP_wrtDIAN_PPG_SYSPP_wrtZero_mean | The ratio of the PPG cardiac cycle systolic phase peak amplitude with respect to dicrotic notch to the PPG cardiac cycle systolic phase peak amplitude with respect to zero, mean across cardiac cycles |
| PPG_AM_RT_DIAN_wrtZero_PPG_SYSPP_wrtZero_mean | The ratio of the PPG cardiac cycle dicrotic notch amplitude with respect to zero to the PPG cardiac cycle systolic phase peak amplitude with respect to zero, mean across cardiac cycles |
| PPG_AM_RT_SD_Dpeak_wrtEpeak_PPG_AM_SD_Bpeak_wrtzero_mean | The ratio of the PPG cardiac cycle second derivative Dpeak amplitude with respect to Epeak to the PPG cardiac cycle second derivative Bpeak amplitude with respect to zero, mean across cardiac cycles |
| PPG_D_RT_Are_SYSPP_wrtDIAN_PPG_D_Are_SYSPO_wrtDIAPE_mean | The ratio of the area under the PPG cardiac cycle systolic phase peak with respect to dicrotic notch to the area under the PPG cardiac cycle systolic phase onset with respect to diastolic phase end point, mean across cardiac cycles |
| PPG_D_RT_SYSPP_wrtDIAPP_PPG_SYSPP_wrtDIAN_mean | The ratio of the PPG cardiac cycle systolic phase peak duration with respect to diastolic phase peak to the PPG cardiac cycle systolic phase peak duration with respect to dicrotic notch, mean across cardiac cycles |
| PPG_NAM_RT_DIAPP_wrtDIAN_PPG_SYSPP_wrtDIAN_mean | The ratio of the normalized PPG cardiac cycle diastolic phase peak amplitude with respect to dicrotic notch to the normalized PPG cardiac cycle systolic phase peak amplitude with |

| | |
|---|---|
| | respect to dicrotic notch, mean across cardiac cycles |
| PPG_NAM_RT_SYSPP_wrtDIAPP_PPG_SYSPP_wrtDIAN_mean | The ratio of the normalized PPG cardiac cycle systolic phase peak amplitude with respect to diastolic phase peak to the normalized PPG cardiac cycle systolic phase peak amplitude with respect to systolic dicrotic notch, mean across cardiac cycles |
| PPG_W_SRP50_ODP50_mean | Sum of the PPG cardiac cycle systolic rise phase width and overall decay phase width at 50% of the cardiac cycle, mean across cardiac cycles |
| PPG_D_RT_SD_Bpeak_wrtDpeak_PPG_D_SD_Bpeak_wrtCpeak_std | The ratio of the PPG cardiac cycle second derivative Bpeak duration with respect to Dpeak to the PPG cardiac cycle second derivative Bpeak duration with respect to Cpeak, standard deviation across cardiac cycles |
| PPG_D_NAre_wsDIAPE_SYSPO_wrtDIAPP_mean | Area under the normalized PPG cardiac cycle systolic phase onset with respect to diastolic phase peak, with subtracted diastolic phase end point, mean across cardiac cycles |
| PPG_NAM_RT_DIAN_wrtDIAPE_PPG_SYSPP_wrtDIAPE_mean | The ratio of the normalized PPG cardiac cycle dicrotic notch with respect to diastolic phase end point to the normalized PPG cardiac cycle systolic phase peak amplitude with respect to diastolic phase end point, mean across cardiac cycles |
| PPG_AM_RT_DIAN_wrtDIAPE_PPG_SYSPP_wrtDIAPE_mean | The ratio of the PPG cardiac cycle dicrotic notch amplitude with respect to diastolic phase end point to the PPG cardiac cycle systolic phase peak amplitude with respect to diastolic phase end point, mean across cardiac cycles |
| PPG_D_NAre_SYSPO_wrtDIAPP_mean | Area under the normalized PPG cardiac cycle systolic phase onset with respect to diastolic phase end point, mean across cardiac cycles |
| PPG_D_NAre_wsDIAPE_DIAN_wrtDIAPP_mean | Area under the normalized PPG cardiac cycle dicrotic notch with respect to diastolic phase peak, with subtracted diastolic phase end point, mean across cardiac cycles |
| PPG_D_SYSPP_wrtDIAN_mean | Duration of the PPG cardiac cycle systolic phase peak with respect to dicrotic notch, mean across cardiac cycles |
| PPG_D_RT_SD_Cpeak_wrtDpeak_PPG_D_SD_Bpeak_wrtDpeak_mean | The ratio of the PPG cardiac cycle second derivative Cpeak duration with respect to Dpeak to the PPG cardiac cycle second |

| | |
|---|---|
| | derivative Bpeak duration with respect to Dpeak, mean across cardiac cycles |
| PPG_AM_FD_Upeak_wrtVpeak_mean | PPG cardiac cycle first derivative Upeak amplitude with respect to Vpeak, mean across cardiac cycles |
| PPG_AM_RT_SD_Bpeak_wrtCpeak_PPG_AM_SD_Apeak_wrtEpeak_mean | The ratio of the PPG cardiac cycle second derivative Bpeak amplitude with respect to Cpeak to the PPG cardiac cycle second derivative Apeak amplitude with respect to Epeak, mean across cardiac cycles |
| PPG_D_NAre_wsDIAPE_SYSPO_wrtDIAPE_mean | Area under the normalized PPG cardiac cycle systolic phase onset with respect to diastolic phase end point, with subtracted diastolic phase end point, mean across cardiac cycles |
| PPG_NAM_RT_DIAN_wrtDIAPE_PPG_SYSPP_wrtSYSPO_mean | The ratio of the normalized PPG cardiac cycle dicrotic notch with respect to diastolic phase end point to the normalized PPG cardiac cycle systolic phase peak amplitude with respect to systolic phase onset, mean across cardiac cycles |
| PPG_D_RT_DIAN_wrtDIAPP_PPG_SYSPP_wrtDIAPP_mean | The ratio of the PPG cardiac cycle dicrotic notch duration with respect to diastolic phase peak to the PPG cardiac cycle systolic phase peak duration with respect to diastolic phase peak, mean across cardiac cycles |
| PPG_AM_RT_DIAN_wrtDIAPE_PPG_SYSPP_wrtSYSPO_mean | The ratio of the PPG cardiac cycle dicrotic notch amplitude with respect to diastolic phase end point to the PPG cardiac cycle systolic phase peak amplitude with respect to systolic phase onset, mean across cardiac cycles |
| PPG_D_RT_NAre_wsDIAPE_DIAN_wrtDIAPP_PPG_D_NAre_wsDIAPE_SYSPP_wrtDIAPE_mean | The ratio of the area under the normalized PPG cardiac cycle dicrotic notch with respect to diastolic phase peak, with subtracted diastolic phase end point to the area under the normalized PPG cardiac cycle systolic phase peak with respect to diastolic phase end point, with subtracted diastolic phase end point, mean across cardiac cycles |
| PPG_NAM_RT_SYSPP_wrtDIAN_PPG_SYSPP_wrtSYSPO_mean | The ratio of the normalized PPG cardiac cycle systolic phase peak amplitude with respect to dicrotic notch to the normalized PPG cardiac cycle systolic phase peak amplitude with respect to systolic phase onset, mean across cardiac cycles |

| | |
|---|---|
| PPG_AM_RT_SD_Bpeak_wrtCpeak_PPG_AM_SD_Apeak_wrtBpeak_mean | The ratio of the PPG cardiac cycle second derivative Bpeak amplitude with respect to Cpeak to the PPG cardiac cycle second derivative Apeak amplitude with respect to Bpeak, mean across cardiac cycles |
| PPG_NAM_RT_DIAN_wrtSYSPO_PPG_SYSPP_wrtSYSPO_mean | The ratio of the normalized PPG cardiac cycle dicrotic notch with respect to systolic phase onset to the normalized PPG cardiac cycle systolic phase peak amplitude with respect to systolic phase onset, mean across cardiac cycles |
| PPG_AM_RT_SD_Apeak_wrtCpeak_PPG_AM_SD_Apeak_wrtBpeak_mean | The ratio of the PPG cardiac cycle second derivative Apeak amplitude with respect to Cpeak to the PPG cardiac cycle second derivative Apeak amplitude with respect to Bpeak, mean across cardiac cycles |
| PPG_AM_SYSPP_wrtDIAN_mean | PPG cardiac cycle systolic phase peak amplitude with respect to dicrotic notch, mean across cardiac cycles |
| PPG_AM_FD_Upeak_wrtzero_mean | PPG cardiac cycle first derivative U peak amplitude with respect to zero, mean across cardiac cycles |
| PPG_AM_SD_Apeak_wrtzero_mean | PPG cardiac cycle second derivative Apeak amplitude with respect to zero, mean across cardiac cycles |
| PPG_D_NAre_SYSPO_wrtDIAPE_mean | Area under the normalized PPG cardiac cycle systolic phase onset with respect to diastolic phase end point, mean across cardiac cycles |
| PPG_AM_RT_DIAN_wrtSYSPO_PPG_SYSPP_wrtSYSPO_mean | The ratio of the PPG cardiac cycle dicrotic notch amplitude with respect to systolic phase onset to the PPG cardiac cycle systolic phase peak amplitude with respect to systolic phase onset, mean across cardiac cycles |
| PPG_AM_RT_SYSPP_wrtDIAN_PPG_SYSPP_wrtSYSPO_mean | The ratio of the PPG cardiac cycle systolic phase peak amplitude with respect to dicrotic notch to the PPG cardiac cycle systolic phase peak amplitude with respect to systolic phase onset, mean across cardiac cycles |
| PPG_D_RT_NAre_DIAN_wrtDIAPP_PPG_D_NAre_SYSPP_wrtDIAPE_mean | The ratio of the area under the normalized PPG cardiac cycle dicrotic notch with respect to diastolic phase peak to the area under the normalized PPG cardiac cycle systolic phase peak with respect to diastolic phase end point, mean across cardiac cycles |

| | |
|---|---|
| PPG_D_Are_DIAN_wrtDIAPP_mean | Area under the PPG cardiac cycle dicrotic notch with respect to diastolic phase peak, mean across cardiac cycles |
| PPG_D_Are_wsDIAPE_DIAN_wrtDIAPP_mean | Area under the PPG cardiac cycle dicrotic notch with respect to diastolic phase peak, with subtracted diastolic phase end point, mean across cardiac cycles |
| PPG_AM_RT_SD_Bpeak_wrtDpeak_PPG_AM_SD_Cpeak_wrtzero_std | The ratio of the PPG cardiac cycle second derivative Bpeak amplitude with respect to Dpeak to the PPG cardiac cycle second derivative Cpeak amplitude with respect to zero, standard deviation across cardiac cycles |
| PPG_W_SRP33_ODP33_mean | Sum of the PPG cardiac cycle systolic rise phase width and overall decay phase width at 33% of the cardiac cycle, mean across cardiac cycles |
| PPG_AM_RT_SD_Bpeak_wrtCpeak_PPG_AM_SD_Bpeak_wrtzero_mean | The ratio of the PPG cardiac cycle second derivative Bpeak amplitude with respect to Cpeak to the PPG cardiac cycle second derivative Bpeak amplitude with respect to zero, mean across cardiac cycles |
| PPG_AM_RT_SD_Cpeak_wrtzero_PPG_AM_SD_Bpeak_wrtzero_mean | The ratio of the PPG cardiac cycle second derivative Cpeak amplitude with respect to zero to the PPG cardiac cycle second derivative Bpeak amplitude with respect to zero, mean across cardiac cycles |
| PPG_W_SRP66_ODP66_mean | Sum of the PPG cardiac cycle systolic rise phase width and overall decay phase width at 66% of the cardiac cycle, mean across cardiac cycles |
| PPG_D_RT_Are_SYSPP_wrtDIAPE_PPG_D_Are_SYSPO_wrtDIAPE_mean | The ratio of the area under the PPG cardiac cycle systolic phase peak with respect to diastolic phase end point to the area under the PPG cardiac cycle systolic phase onset with respect to diastolic phase end point, mean across cardiac cycles |
| PPG_NAM_RT_DIAN_wrtDIAPE_PPG_SYSPP_wrtDIAN_mean | The ratio of the normalized PPG cardiac cycle dicrotic notch with respect to diastolic phase end point to the normalized PPG cardiac cycle systolic phase peak amplitude with respect to dicrotic notch, mean across cardiac cycles |
| PPG_NAM_RT_SYSPP_wrtDIAPE_PPG_SYSPP_wrtDIAN_mean | The ratio of the normalized PPG cardiac cycle systolic phase peak amplitude with respect to diastolic phase end point to the normalized PPG cardiac cycle systolic phase peak |

| | |
|---|---|
| | amplitude with respect to dicrotic notch, mean across cardiac cycles |
| PPG_RT_ODP75_W_SRP75_ODP75_W_mean | The ratio of the PPG cardiac cycle overall decay phase width at 75% of the cardiac cycle to the sum of the PPG cardiac cycle systolic rise phase width and overall decay phase width at 75% of the cardiac cycle, mean across cardiac cycles |
| PPG_D_RT_Are_wsDIAPE_SYSPP_wrtDIAPE_PPG_D_Are_wsDIAPE_SYSPO_wrtDIAPE_mean | The ratio of the area under the PPG cardiac cycle systolic phase peak with respect to diastolic phase end point, with subtracted diastolic phase end point to the area under the PPG cardiac cycle systolic phase onset with respect to diastolic phase end point, with subtracted diastolic phase end point, mean across cardiac cycles |
| PPG_AM_RT_DIAPP_wrtDIAN_PPG_SYSPP_wrtDIAPP_mean | The ratio of the PPG cardiac cycle diastolic phase peak amplitude with respect to dicrotic notch to the PPG cardiac cycle systolic phase peak amplitude with respect to diastolic phase peak, mean across cardiac cycles |
| PPG_RT_SRP75_W_SRP75_ODP75_W_mean | The ratio of the PPG cardiac cycle systolic rise phase width at 75% of the cardiac cycle to the sum of the PPG cardiac cycle systolic rise phase width and overall decay phase width at 75% of the cardiac cycle, mean across cardiac cycles |
| PPG_AM_RT_SD_Bpeak_wrtEpeak_PPG_AM_SD_Bpeak_wrtDpeak_mean | The ratio of the PPG cardiac cycle second derivative Bpeak amplitude with respect to Epeak to the PPG cardiac cycle second derivative Bpeak amplitude with respect to Dpeak, mean across cardiac cycles |
| PPG_AM_RT_SD_Dpeak_wrtEpeak_PPG_AM_SD_Bpeak_wrtDpeak_mean | The ratio of the PPG cardiac cycle second derivative Dpeak amplitude with respect to Epeak to the PPG cardiac cycle second derivative Bpeak amplitude with respect to Dpeak, mean across cardiac cycles |
| PPG_NAM_RT_DIAN_wrtZero_PPG_SYSPP_wrtDIAN_mean | The ratio of the normalized PPG cardiac cycle dicrotic notch with respect to zero to the normalized PPG cardiac cycle systolic phase peak amplitude with respect to dicrotic notch, mean across cardiac cycles |
| PPG_NAM_RT_DIAN_wrtDIAPE_PPG_SYSPP_wrtZero_mean | The ratio of the normalized PPG cardiac cycle dicrotic notch with respect to diastolic phase end point to the normalized PPG cardiac cycle |

| | |
|---|---|
| | systolic phase peak amplitude with respect to zero, mean across cardiac cycles |
| PPG_AM_RT_SD_Bpeak_wrtDpeak_PPG_AM_SD_Dpeak_wrtzero_mean | The ratio of the PPG cardiac cycle second derivative Bpeak amplitude with respect to Dpeak to the PPG cardiac cycle second derivative Dpeak amplitude with respect to zero, mean across cardiac cycles |
| PPG_AM_RT_SD_Cpeak_wrtEpeak_PPG_AM_SD_Bpeak_wrtDpeak_mean | The ratio of the PPG cardiac cycle second derivative Cpeak amplitude with respect to Epeak to the PPG cardiac cycle second derivative Bpeak amplitude with respect to Dpeak, mean across cardiac cycles |
| PPG_D_RT_SD_Bpeak_wrtEpeak_PPG_D_SD_Bpeak_wrtCpeak_std | The ratio of the PPG cardiac cycle second derivative Bpeak duration with respect to Epeak to the PPG cardiac cycle second derivative Bpeak duration with respect to Cpeak, standard deviation across cardiac cycles |
| PPG_AM_RT_DIAN_wrtDIAPE_PPG_SYSPP_wrtDIAN_mean | The ratio of the PPG cardiac cycle dicrotic notch amplitude with respect to diastolic phase end point to the PPG cardiac cycle systolic phase peak amplitude with respect to dicrotic notch, mean across cardiac cycles |
| PPG_AM_RT_SYSPP_wrtDIAPE_PPG_SYSPP_wrtDIAN_mean | The ratio of the PPG cardiac cycle systolic phase peak amplitude with respect to diastolic phase end point to the PPG cardiac cycle systolic phase peak amplitude with respect to dicrotic notch, mean across cardiac cycles |
| PPG_D_RT_SD_Cpeak_wrtEpeak_PPG_D_SD_Bpeak_wrtCpeak_std | The ratio of the PPG cardiac cycle second derivative Cpeak duration with respect to Epeak to the PPG cardiac cycle second derivative Bpeak duration with respect to Cpeak, standard deviation across cardiac cycles |
| PPG_AM_RT_SD_Cpeak_wrtEpeak_PPG_AM_SD_Bpeak_wrtCpeak_std | The ratio of the PPG cardiac cycle second derivative Cpeak amplitude with respect to Epeak to the PPG cardiac cycle second derivative Bpeak amplitude with respect to Cpeak, standard deviation across cardiac cycles |
| PPG_AM_RT_SD_Bpeak_wrtEpeak_PPG_AM_SD_Bpeak_wrtCpeak_std | The ratio of the PPG cardiac cycle second derivative Bpeak amplitude with respect to Epeak to the PPG cardiac cycle second derivative Bpeak amplitude with respect to Cpeak, standard deviation across cardiac cycles |
| PPG_NAM_RT_DIAPP_wrtDIAN_PPG_SYSPP_wrtDIAPP_mean | The ratio of the normalized PPG cardiac cycle diastolic phase peak amplitude with respect to |

| | |
|---|---|
| | dicrotic notch to the normalized PPG cardiac cycle systolic phase peak amplitude with respect to diastolic phase peak, mean across cardiac cycles |
| PPG_D_RT_NAre_wsDIAPE_SYSPP_wrtDIAPE_PPG_D_NAre_wsDIAPE_SYSPO_wrtDIAPE_mean | The ratio of the area under the normalized PPG cardiac cycle systolic phase peak with respect to diastolic phase end point, with subtracted diastolic phase end point to the area under the normalized PPG cardiac cycle systolic phase onset with respect to diastolic phase endpoint, with subtracted diastolic phase end point, mean across cardiac cycles |
| PPG_AM_RT_SD_Dpeak_wrtEpeak_PPG_AM_SD_Dpeak_wrtzero_mean | The ratio of the PPG cardiac cycle second derivative Dpeak amplitude with respect to Epeak to the PPG cardiac cycle second derivative Dpeak amplitude with respect to zero, mean across cardiac cycles |
| PPG_AM_RT_SD_Epeak_wrtzero_PPG_AM_SD_Dpeak_wrtzero_mean | The ratio of the PPG cardiac cycle second derivative Epeak amplitude with respect to zero to the PPG cardiac cycle second derivative Dpeak amplitude with respect to zero, mean across cardiac cycles |
| PPG_D_RT_SYSPP_wrtDIAPE_PPG_SYSPO_wrtSYSPP_mean | The ratio of the PPG cardiac cycle systolic phase peak duration with respect to diastolic phase end point to the PPG cardiac cycle systolic phase onset duration with respect to systolic phase peak, mean across cardiac cycles |
| PPG_D_RT_NAre_SYSPP_wrtDIAPE_PPG_D_NAre_SYSPO_wrtDIAPE_mean | The ratio of the area under the normalized PPG cardiac cycle systolic phase peak with respect to diastolic phase end point to the area under the normalized PPG cardiac cycle systolic phase onset with respect to diastolic phase end point, mean across cardiac cycles |
| PPG_NAM_RT_DIAPP_wrtDIAN_PPG_SYSPP_wrtDIAPE_mean | The ratio of the normalized PPG cardiac cycle diastolic phase peak amplitude with respect to dicrotic notch to the normalized PPG cardiac cycle systolic phase peak amplitude with respect to diastolic phase end point, mean across cardiac cycles |
| PPG_NAM_RT_DIAN_wrtSYSPO_PPG_SYSPP_wrtDIAN_mean | The ratio of the normalized PPG cardiac cycle dicrotic notch with respect to systolic phase onset to the normalized PPG cardiac cycle systolic phase peak amplitude with respect to dicrotic notch, mean across cardiac cycles |

| | |
|---|---|
| PPG_AM_RT_SD_Bpeak_wrtCpeak_PPG_AM_SD_Dpeak_wrtzero_mean | The ratio of the PPG cardiac cycle second derivative Bpeak amplitude with respect to Cpeak to the PPG cardiac cycle second derivative Dpeak amplitude with respect to zero, mean across cardiac cycles |
| PPG_AM_RT_SD_Apeak_wrtBpeak_PPG_AM_SD_Dpeak_wrtzero_mean | The ratio of the PPG cardiac cycle second derivative Apeak amplitude with respect to Bpeak to the PPG cardiac cycle second derivative Dpeak amplitude with respect to zero, mean across cardiac cycles |
| PPG_AM_RT_DIAPP_wrtDIAN_PPG_SYSPP_wrtDIAPE_mean | The ratio of the PPG cardiac cycle diastolic phase peak amplitude with respect to dicrotic notch to the PPG cardiac cycle systolic phase peak amplitude with respect to diastolic phase end point, mean across cardiac cycles |
| PPG_D_RT_SYSPO_wrtDIAPE_PPG_SYSPO_wrtSYSPP_mean | The ratio of the PPG cardiac cycle systolic phase onset duration with respect to diastolic phase end point to the PPG cardiac cycle systolic phase onset duration with respect to systolic phase peak, mean across cardiac cycles |
| PPG_AM_RT_DIAN_wrtSYSPO_PPG_SYSPP_wrtDIAN_mean | The ratio of the PPG cardiac cycle dicrotic notch amplitude with respect to systolic phase onset to the PPG cardiac cycle systolic phase peak amplitude with respect to dicrotic notch, mean across cardiac cycles |
| PPG_NAM_DIAN_wrtDIAPE_mean | Normalized PPG cardiac cycle dicrotic notch amplitude with respect to diastolic phase end point, mean across cardiac cycles |
| PPG_NAM_DIAN_wrtSYSPO_mean | Normalized PPG cardiac cycle dicrotic notch amplitude with respect to systolic phase onset, mean across cardiac cycles |
| PPG_D_RT_SD_Apeak_wrtCpeak_PPG_D_SD_Apeak_wrtBpeak_mean | The ratio of the PPG cardiac cycle second derivative Apeak duration with respect to Cpeak to the PPG cardiac cycle second derivative Apeak duration with respect to Bpeak, mean across cardiac cycles |
| PPG_AM_RT_SD_Cpeak_wrtEpeak_PPG_AM_SD_Bpeak_wrtEpeak_mean | The ratio of the PPG cardiac cycle second derivative Cpeak amplitude with respect to Epeak to the PPG cardiac cycle second derivative Bpeak amplitude with respect to Epeak, mean across cardiac cycles |
| PPG_AM_DIAN_wrtSYSPO_mean | PPG cardiac cycle dicrotic notch amplitude with respect to systolic phase onset, mean across cardiac cycles |

| | |
|---|---|
| PPG_D_NAvg_SYSPP_wrtDIAN_mean | PPG cardiac cycle dicrotic notch amplitude with respect to systolic phase onset, mean across cardiac cycles |
| PPG_AM_RT_DIAN_wrtDIAPE_PPG_SYSPP_wrtZero_mean | The ratio of the PPG cardiac cycle dicrotic notch amplitude with respect to diastolic phase end point to the PPG cardiac cycle systolic phase peak amplitude with respect to zero, mean across cardiac cycles |
| PPG_D_RT_SD_Bpeak_wrtCpeak_PPG_D_SD_Apeak_wrtCpeak_mean | The ratio of the PPG cardiac cycle second derivative Bpeak duration with respect to Cpeak to the PPG cardiac cycle second derivative Apeak duration with respect to Cpeak, mean across cardiac cycles |
| PPG_AM_RT_DIAN_wrtSYSPO_PPG_SYSPP_wrtZero_mean | The ratio of the PPG cardiac cycle dicrotic notch amplitude with respect to systolic phase onset to the PPG cardiac cycle systolic phase peak amplitude with respect to zero, mean across cardiac cycles |
| PPG_AM_RT_SD_Cpeak_wrtEpeak_PPG_AM_SD_Bpeak_wrtzero_mean | The ratio of the PPG cardiac cycle second derivative Cpeak amplitude with respect to Epeak to the PPG cardiac cycle second derivative Bpeak amplitude with respect to zero, mean across cardiac cycles |
| PPG_D_RT_Are_wsDIAPE_SYSPP_wrtDIAPE_PPG_D_Are_wsDIAPE_SYSPO_wrtSYSPP_mean | The ratio of the area under the PPG cardiac cycle systolic phase peak with respect to diastolic phase end point, with subtracted diastolic phase end point to the area under the PPG cardiac cycle systolic phase onset with respect to systolic phase peak, with subtracted diastolic phase end point, mean across cardiac cycles |
| PPG_D_RT_SD_Bpeak_wrtCpeak_PPG_D_SD_Apeak_wrtBpeak_mean | The ratio of the PPG cardiac cycle second derivative Bpeak duration with respect to Cpeak to the PPG cardiac cycle second derivative Apeak duration with respect to Bpeak, mean across cardiac cycles |
| PPG_W_SRP75_ODP75_mean | Sum of the PPG cardiac cycle systolic rise phase width and overall decay phase width at 75% of the cardiac cycle, mean across cardiac cycles |
| PPG_D_RT_NAre_DIAN_wrtDIAPP_PPG_D_NAre_SYSPO_wrtDIAPP_mean | The ratio of the area under the normalized PPG cardiac cycle dicrotic notch with respect to diastolic phase peak to the area under the normalized PPG cardiac cycle systolic phase |

| | |
|---|---|
| | onset with respect to diastolic phase peak, mean across cardiac cycles |
| PPG_D_RT_Are_SYSPP_wrtDIAPE_PPG_D_Are_SYSPO_wrtSYSPP_mean | The ratio of the area under the PPG cardiac cycle systolic phase peak with respect to diastolic phase end point to the area under the PPG cardiac cycle systolic phase onset with respect to systolic phase peak, mean across cardiac cycles |
| PPG_D_RT_Are_SYSPO_wrtDIAPE_PPG_D_Are_SYSPO_wrtSYSPP_mean | The ratio of the area under the PPG cardiac cycle systolic phase onset with respect to diastolic phase end point to the area under the PPG cardiac cycle systolic phase onset with respect to systolic phase peak, mean across cardiac cycles |
| PPG_NAM_RT_DIAN_wrtSYSPO_PPG_SYSPP_wrtZero_mean | The ratio of the normalized PPG cardiac cycle dicrotic notch with respect to systolic phase onset to the normalized PPG cardiac cycle systolic phase peak amplitude with respect to zero, mean across cardiac cycles |
| PPG_D_RT_NAre_wsDIAPE_DIAN_wrtDIAPP_PPG_D_NAre_wsDIAPE_SYSPO_wrtDIAPP_mean | The ratio of the area under the normalized PPG cardiac cycle dicrotic notch with respect to diastolic phase peak, with subtracted diastolic phase end point to the area under the normalized PPG cardiac cycle systolic phase onset with respect to diastolic phase peak, with subtracted diastolic phase end point, mean across cardiac cycles |
| PPG_AM_RT_DIAN_wrtZero_PPG_SYSPP_wrtSYSPO_mean | The ratio of the PPG cardiac cycle dicrotic notch amplitude with respect to zero to the PPG cardiac cycle systolic phase peak amplitude with respect to systolic phase onset, mean across cardiac cycles |
| PPG_D_RT_Are_wsDIAPE_DIAN_wrtDIAPP_PPG_D_Are_wsDIAPE_SYSPP_wrtDIAPE_mean | The ratio of the area under the PPG cardiac cycle dicrotic notch with respect to diastolic phase peak, with subtracted diastolic phase end point to the area under the PPG cardiac cycle systolic phase peak with respect to diastolic phase end point, with subtracted diastolic phase end point, mean across cardiac cycles |
| PPG_D_RT_SYSPP_wrtDIAPE_PPG_SYSPP_wrtDIAN_mean | The ratio of the PPG cardiac cycle systolic phase peak duration with respect to diastolic phase end point to the PPG cardiac cycle systolic |

| | |
|---|---|
| | phase peak duration with respect to dicrotic notch, mean across cardiac cycles |
| PPG_D_RT_DIAN_wrtDIAPE_PPG_SYSPP_wrtDIAN_mean | The ratio of the PPG cardiac cycle dicrotic notch duration with respect to diastolic phase end point to the PPG cardiac cycle systolic phase peak duration with respect to dicrotic notch, mean across cardiac cycles |
| PPG_D_NMed_SYSPO_wrtDIAPE_mean | Median duration of the normalized PPG cardiac cycle systolic phase onset with respect to diastolic phase end point, mean across cardiac cycles |
| PPG_W_SRP25_ODP25_mean | Sum of the PPG cardiac cycle systolic rise phase width and overall decay phase width at 25% of the cardiac cycle, mean across cardiac cycles |
| PPG_AM_RT_DIAN_wrtZero_PPG_SYSPP_wrtDIAN_mean | The ratio of the PPG cardiac cycle dicrotic notch amplitude with respect to zero to the PPG cardiac cycle systolic phase peak amplitude with respect to dicrotic notch, mean across cardiac cycles |
| PPG_AM_RT_DIAPP_wrtDIAN_PPG_SYSPP_wrtSYSPO_mean | The ratio of the PPG cardiac cycle diastolic phase peak amplitude with respect to dicrotic notch to the PPG cardiac cycle systolic phase peak amplitude with respect to systolic phase onset, mean across cardiac cycles |
| PPG_NAM_RT_DIAPP_wrtDIAN_PPG_SYSPP_wrtSYSPO_mean | The ratio of the normalized PPG cardiac cycle diastolic phase peak amplitude with respect to dicrotic notch to the normalized PPG cardiac cycle systolic phase peak amplitude with respect to systolic phase onset, mean across cardiac cycles |
| PPG_D_RT_SD_Cpeak_wrtEpeak_PPG_D_SD_Bpeak_wrtCpeak_mean | The ratio of the PPG cardiac cycle second derivative Cpeak duration with respect to Epeak to the PPG cardiac cycle second derivative Bpeak duration with respect to Cpeak, mean across cardiac cycles |
| PPG_D_RT_NAre_DIAN_wrtDIAPP_PPG_D_NAre_SYSPO_wrtDIAPE_mean | The ratio of the area under the normalized PPG cardiac cycle dicrotic notch peak with respect to diastolic phase peak to the area under the normalized PPG cardiac cycle systolic phase onset with respect to diastolic phase end point, mean across cardiac cycles |
| PPG_RT_ODP10_W_SRP10_W_mean | The ratio of the PPG cardiac cycle overall decay phase width at 10% of the cardiac cycle to the PPG cardiac cycle systolic rise phase width at |

| | |
|---|---|
| | 10% of the cardiac cycle, mean across cardiac cycles |
| PPG_D_NRMS_DIAN_wrtDIAPE_mean | Root mean square duration of the normalized PPG cardiac cycle dicrotic notch with respect to diastolic phase end point, mean across cardiac cycles |
| PPG_NAM_RT_DIAPP_wrtDIAN_PPG_SYSPP_wrtZero_mean | The ratio of the normalized PPG cardiac cycle diastolic phase peak amplitude with respect to dicrotic notch to the normalized PPG cardiac cycle systolic phase peak amplitude with respect to zero, mean across cardiac cycles |
| PPG_D_RT_NAre_wsDIAPE_SYSPO_wrtDIAPP_PPG_D_NAre_wsDIAPE_SYSPO_wrtDIAN_mean | The ratio of the area under the normalized PPG cardiac cycle systolic phase onset with respect to diastolic phase peak, with subtracted diastolic phase end point to the area under the normalized PPG cardiac cycle systolic phase onset with respect to dicrotic notch, with subtracted diastolic phase end point, mean across cardiac cycles |
| PPG_D_RT_SD_Bpeak_wrtEpeak_PPG_D_SD_Bpeak_wrtCpeak_mean | The ratio of the PPG cardiac cycle second derivative Bpeak duration with respect to Epeak to the PPG cardiac cycle second derivative Bpeak duration with respect to Cpeak, mean across cardiac cycles |
| PPG_D_RT_NAre_wsDIAPE_SYSPP_wrtDIAPE_PPG_D_NAre_wsDIAPE_SYSPO_wrtSYSPP_mean | The ratio of the area under the normalized PPG cardiac cycle systolic phase peak with respect to diastolic phase end point, with subtracted diastolic phase end point to the area under the normalized PPG cardiac cycle systolic phase onset with respect to systolic phase peak, with subtracted diastolic phase end point, mean across cardiac cycles |
| PPG_D_RT_NAre_wsDIAPE_SYSPO_wrtDIAPE_PPG_D_NAre_wsDIAPE_SYSPO_wrtSYSPP_mean | The ratio of the area under the normalized PPG cardiac cycle systolic phase onset with respect to diastolic phase end point, with subtracted diastolic phase end point to the area under the normalized PPG cardiac cycle systolic phase onset with respect to systolic phase peak, with subtracted diastolic phase end point, mean across cardiac cycles |
| PPG_D_NRMS_SYSPP_wrtDIAN_mean | Root mean square duration of the normalized PPG cardiac cycle systolic phase peak with respect of dicrotic notch, mean across cardiac cycles |

| | |
|---|---|
| PPG_RT_ODP10_W_SRP10_ODP10_W_mean | The ratio of the PPG cardiac cycle overall decay phase width at 10% of the cardiac cycle to the sum of the PPG cardiac cycle systolic rise phase width and overall decay phase width at 10% of the cardiac cycle, mean across cardiac cycles |
| PPG_D_RT_Are_wsDIAPE_SYSPO_wrtDIAPE_PPG_D_Are_wsDIAPE_SYSPO_wrtSYSPP_mean | The ratio of the area under the PPG cardiac cycle systolic phase onset with respect to diastolic phase end point, with subtracted diastolic phase end point to the area under the PPG cardiac cycle systolic phase onset with respect to systolic phase peak, with subtracted diastolic phase end point, mean across cardiac cycles |
| PPG_D_NAvg_SYSPO_wrtSYSPP_mean | Average duration of the normalized PPG cardiac cycle systolic phase onset with respect to systolic phase peak, mean across cardiac cycles |
| PPG_D_NMed_SYSPO_wrtSYSPP_mean | Median duration of the normalized PPG cardiac cycle systolic phase onset with respect to systolic phase peak, mean across cardiac cycles |
| PPG_AM_RT_SD_Cpeak_wrtEpeak_PPG_AM_SD_Bpeak_wrtDpeak_std | The ratio of the PPG cardiac cycle second derivative Cpeak amplitude with respect to Epeak to the PPG cardiac cycle second derivative Bpeak amplitude with respect to Dpeak, standard deviation across cardiac cycles |
| PPG_D_RT_NAre_DIAN_wrtDIAPP_PPG_D_NAre_SYSPO_wrtDIAN_mean | The ratio of the area under the normalized PPG cardiac cycle dicrotic notch with respect to diastolic phase peak to the area under the normalized PPG cardiac cycle systolic phase onset with respect to dicrotic notch, mean across cardiac cycles |
| PPG_RT_SRP10_W_SRP10_ODP10_W_mean | The ratio of the PPG cardiac cycle systolic rise phase width at 10% of the cardiac cycle to the sum of the PPG cardiac cycle systolic rise phase width and overall decay phase width at 10% of the cardiac cycle, mean across cardiac cycles |
| PPG_D_RT_NAre_SYSPO_wrtDIAPP_PPG_D_NAre_SYSPO_wrtDIAN_mean | The ratio of the area under the normalized PPG cardiac cycle systolic phase onset with respect to diastolic phase peak to the area under the normalized PPG cardiac cycle systolic phase onset with respect to dicrotic notch, mean across cardiac cycles |
| PPG_D_RT_NAre_wsDIAPE_DIAN_wrtDIAPP_PPG_D_NAre_wsDIAPE_SYSPO_wrtDIAN_mean | The ratio of the area under the normalized PPG cardiac cycle dicrotic notch with respect to |

| | |
|---|---|
| | diastolic phase peak, with subtracted diastolic phase end point to the area under the normalized PPG cardiac cycle systolic phase onset with respect to dicrotic notch, with subtracted diastolic phase end point, mean across cardiac cycles |
| PPG_AM_RT_SD_Dpeak_wrtEpeak_PPG_AM_SD_Apeak_wrtBpeak_mean | The ratio of the PPG cardiac cycle second derivative Dpeak amplitude with respect to Epeak to the PPG cardiac cycle second derivative Apeak amplitude with respect to Bpeak, mean across cardiac cycles |
| PPG_pkamp_mean | Mean across cardiac cycles of the amplitude of the first peak observed in the FFT of the PPG cardiac cycle |
| PPG_NAM_DIAPP_wrtDIAN_mean | Normalized PPG cardiac cycle diastolic phase peak amplitude with respect to dicrotic notch, mean across cardiac cycles |
| PPG_D_RT_Are_DIAN_wrtDIAPP_PPG_D_Are_SYSPP_wrtDIAPE_mean | The ratio of the area under the PPG cardiac cycle dicrotic notch with respect to diastolic phase peak to the area under the PPG cardiac cycle systolic phase peak with respect to diastolic phase end point, mean across cardiac cycles |
| PPG_NAM_RT_DIAN_wrtSYSPO_PPG_SYSPP_wrtDIAPP_mean | The ratio of the normalized PPG cardiac cycle dicrotic notch with respect to systolic phase onset to the normalized PPG cardiac cycle systolic phase peak amplitude with respect to diastolic phase peak, mean across cardiac cycles |
| PPG_RT_ODP75_W_SRP75_W_mean | The ratio of the PPG cardiac cycle overall decay phase width at 75% of the cardiac cycle to the PPG cardiac cycle systolic rise phase width at 75% of the cardiac cycle, mean across cardiac cycles |
| PPG_D_RT_NAre_SYSPP_wrtDIAPE_PPG_D_NAre_SYSPO_wrtSYSPP_mean | The ratio of the area under the normalized PPG cardiac cycle systolic phase peak with respect to diastolic phase end point to the area under the normalized PPG cardiac cycle systolic phase onset with respect to systolic phase peak, mean across cardiac cycles |
| PPG_D_RT_NAre_SYSPO_wrtDIAPE_PPG_D_NAre_SYSPO_wrtSYSPP_mean | The ratio of the area under the normalized PPG cardiac cycle systolic phase onset with respect to diastolic phase end point to the area under the normalized PPG cardiac cycle systolic phase |

| | |
|---|---|
| | onset with respect to systolic phase peak, mean across cardiac cycles |
| PPG_D_NRMS_SYSPO_wrtDIAPE_mean | Root mean square duration of the normalized PPG cardiac cycle systolic phase onset with respect to diastolic phase end point, mean across cardiac cycles |
| PPG_RT_SRP66_W_SRP66_ODP66_W_mean | The ratio of the PPG cardiac cycle systolic rise phase width at 66% of the cardiac cycle to the sum of the PPG cardiac cycle systolic rise phase width and overall decay phase width at 66% of the cardiac cycle, mean across cardiac cycles |
| PPG_AM_DIAN_wrtDIAPE_mean | PPG cardiac cycle dicrotic notch amplitude with respect to diastolic phase end point, mean across cardiac cycles |
| PPG_D_RT_SYSPP_wrtDIAPE_PPG_SYSPO_wrtDIAPE_mean | The ratio of the PPG cardiac cycle systolic phase peak duration with respect to diastolic phase end point to the PPG cardiac cycle systolic phase onset duration with respect to diastolic phase end point, mean across cardiac cycles |
| PPG_AM_RT_DIAN_wrtZero_PPG_SYSPP_wrtDIAPE_mean | The ratio of the PPG cardiac cycle dicrotic notch amplitude with respect to zero to the PPG cardiac cycle systolic phase peak amplitude with respect to diastolic phase end point, mean across cardiac cycles |
| PPG_D_NAre_wsDIAPE_SYSPO_wrtDIAN_mean | Area under the normalized PPG cardiac cycle systolic phase onset with respect to dicrotic notch, with subtracted diastolic phase end point, mean across cardiac cycles |
| PPG_RT_ODP66_W_SRP66_ODP66_W_mean | The ratio of the PPG cardiac cycle overall decay phase width at 66% of the cardiac cycle to the sum of the PPG cardiac cycle systolic rise phase width and overall decay phase width at 66% of the cardiac cycle, mean across cardiac cycles |
| PPG_D_NAre_SYSPO_wrtDIAN_mean | Area under the normalized PPG cardiac cycle systolic phase onset with respect to dicrotic notch, mean across cardiac cycles |
| PPG_D_Are_wsDIAPE_SYSPO_wrtDIAPP_mean | Area under the PPG cardiac cycle systolic phase onset with respect to diastolic phase peak, with subtracted diastolic phase end point, mean across cardiac cycles |
| PPG_D_RT_Are_wsDIAPE_DIAN_wrtDIAPP_PPG_D_Are_wsDIAPE_SYSPO_wrtDIAN_mean | The ratio of the area under the PPG cardiac cycle dicrotic notch with respect to diastolic phase peak, with subtracted diastolic phase end point to the area under the PPG cardiac |

| | |
|---|---|
| | cycle systolic phase onset with respect to dicrotic notch, with subtracted diastolic phase end point, mean across cardiac cycles |
| PPG_AM_DIAPP_wrtDIAN_mean | PPG cardiac cycle diastolic phase peak amplitude with respect to dicrotic notch, mean across cardiac cycles |
| PPG_D_Med_SYSPO_wrtDIAPE_mean | Median duration of the PPG cardiac cycle systolic phase onset with respect to diastolic phase end point, mean across cardiac cycles |
| PPG_AM_RT_DIAN_wrtDIAPE_PPG_SYSPP_wrtDIAPP_mean | The ratio of the PPG cardiac cycle dicrotic notch amplitude with respect to diastolic phase end point to the PPG cardiac cycle systolic phase peak amplitude with respect to diastolic phase peak, mean across cardiac cycles |
| PPG_D_RT_Are_DIAN_wrtDIAPP_PPG_D_Are_SYSPO_wrtDIAN_mean | The ratio of the area under the/PPG cardiac cycle dicrotic notch with respect to diastolic phase peak to the area under the PPG cardiac cycle systolic phase onset with respect to dicrotic notch, mean across cardiac cycles |
| PPG_D_RT_Are_SYSPO_wrtDIAPP_PPG_D_Are_SYSPO_wrtDIAN_mean | The ratio of the area under the PPG cardiac cycle systolic phase onset with respect to diastolic phase peak to the area under the PPG cardiac cycle systolic phase onset with respect to dicrotic notch, mean across cardiac cycles |
| PPG_D_Are_SYSPO_wrtDIAPP_mean | Area under the PPG cardiac cycle systolic phase onset with respect to diastolic phase peak, mean across cardiac cycles |
| PPG_AM_RT_DIAN_wrtSYSPO_PPG_SYSPP_wrtDIAPP_mean | The ratio of the PPG cardiac cycle dicrotic notch amplitude with respect to systolic phase onset to the PPG cardiac cycle systolic phase peak amplitude with respect to diastolic phase peak, mean across cardiac cycles |
| PPG_D_RT_NAre_wsDIAPE_DIAN_wrtDIAPP_PPG_D_NAre_wsDIAPE_SYSPO_wrtDIAPE_mean | The ratio of the area under the normalized PPG cardiac cycle dicrotic notch with respect to diastolic phase peak, with subtracted diastolic phase end point to the area under the normalized PPG cardiac cycle systolic phase onset with respect to diastolic phase end point, mean across cardiac cycles |
| PPG_D_RT_Are_wsDIAPE_SYSPO_wrtDIAPP_PPG_D_Are_wsDIAPE_SYSPO_wrtDIAN_mean | The ratio of the area under the PPG cardiac cycle systolic phase onset with respect to diastolic phase peak, with subtracted diastolic phase end point to the area under the PPG cardiac cycle systolic phase onset with respect |

| | |
|---|---|
| | to dicrotic notch, with subtracted diastolic phase end point, mean across cardiac cycles |
| PPG_D_RT_SD_Bpeak_wrtCpeak_PPG_D_SD_Apeak_wrtEpeak_mean | The ratio of the PPG cardiac cycle second derivative Bpeak duration with respect to Cpeak to the PPG cardiac cycle second derivative Apeak duration with respect to Epeak, mean across cardiac cycles |
| PPG_AM_SD_Apeak_wrtEpeak_mean | PPG cardiac cycle second derivative Apeak amplitude with respect to Epeak, mean across cardiac cycles |
| PPG_AM_RT_DIAPP_wrtDIAN_PPG_SYSPP_wrtZero_mean | The ratio of the PPG cardiac cycle diastolic phase peak amplitude with respect to dicrotic notch to the PPG cardiac cycle systolic phase peak amplitude with respect to zero, mean across cardiac cycles |
| PPG_D_RT_DIAN_wrtDIAPE_PPG_SYSPP_wrtDIAPE_mean | The ratio of the PPG cardiac cycle dicrotic notch duration with respect to diastolic phase end point to the PPG cardiac cycle systolic phase peak duration with respect to diastolic phase end point, mean across cardiac cycles |
| PPG_D_RT_NAre_wsDIAPE_SYSPP_wrtDIAPE_PPG_D_NAre_wsDIAPE_SYSPP_wrtDIAN_mean | The ratio of the area under the normalized PPG cardiac cycle systolic phase peak with respect to diastolic phase end point, with subtracted diastolic phase end point to the area under the normalized PPG cardiac cycle systolic phase peak with respect to dicrotic notch, with subtracted diastolic phase end point, mean across cardiac cycles |
| PPG_D_RT_Are_wsDIAPE_DIAN_wrtDIAPP_PPG_D_Are_wsDIAPE_SYSPO_wrtDIAPP_mean | The ratio of the area under the PPG cardiac cycle dicrotic notch with respect to diastolic phase peak, with subtracted diastolic phase end point to the area under the PPG cardiac cycle systolic phase onset with respect to diastolic phase peak, with subtracted diastolic phase end point, mean across cardiac cycles |
| PPG_AM_RT_FD_Vpeak_wrtWpeak_PPG_AM_FD_Wpeak_wrtzero_std | The ratio of the PPG cardiac cycle first derivative Vpeak amplitude with respect to Wpeak to the PPG cardiac cycle first derivative Wpeak amplitude with respect to zero, standard deviation across cardiac cycles |
| PPG_D_NMed_DIAN_wrtDIAPP_mean | Median duration of the normalized PPG cardiac cycle dicrotic notch with respect to diastolic phase peak, mean across cardiac cycles |

| | |
|---|---|
| PPG_NAM_RT_DIAN_wrtDIAPE_PPG_SYSPP_wrtDIAPP_mean | The ratio of the normalized PPG cardiac cycle dicrotic notch with respect to diastolic phase end point to the normalized PPG cardiac cycle systolic phase peak amplitude with respect to diastolic phase peak, mean across cardiac cycles |
| PPG_AM_RT_FD_Upeak_wrtVpeak_PPG_AM_FD_Wpeak_wrtzero_mean | The ratio of the PPG cardiac cycle first derivative Upeak amplitude with respect to Vpeak to the PPG cardiac cycle first derivative W peak amplitude with respect to zero, mean across cardiac cycles |
| PPG_D_RT_NAre_DIAN_wrtDIAPE_PPG_D_NAre_SYSPP_wrtDIAPE_mean | The ratio of the area under the normalized PPG cardiac cycle dicrotic notch with respect to diastolic phase end point to the area under the normalized PPG cardiac cycle systolic phase peak with respect to diastolic phase end point, mean across cardiac cycles |
| PPG_D_RT_SD_Apeak_wrtEpeak_PPG_D_SD_Apeak_wrtBpeak_mean | The ratio of the PPG cardiac cycle second derivative Apeak duration with respect to Epeak to the PPG cardiac cycle second derivative Apeak duration with respect to Bpeak, mean across cardiac cycles |
| PPG_AM_RT_SD_Cpeak_wrtDpeak_PPG_AM_SD_Bpeak_wrtDpeak_std | The ratio of the PPG cardiac cycle second derivative Cpeak amplitude with respect to Dpeak to the PPG cardiac cycle second derivative Bpeak amplitude with respect to Dpeak, standard deviation across cardiac cycles |
| PPG_D_RT_NAre_SYSPP_wrtDIAPE_PPG_D_NAre_SYSPP_wrtDIAN_mean | The ratio of the area under the normalized PPG cardiac cycle systolic phase peak with respect to diastolic phase end point to the area under the normalized PPG cardiac cycle systolic phase peak with respect to dicrotic notch, mean across cardiac cycles |
| PPG_D_RT_NAre_DIAN_wrtDIAPE_PPG_D_NAre_SYSPP_wrtDIAN_mean | The ratio of the area under the normalized PPG cardiac cycle dicrotic notch with respect to diastolic phase end point to the area under the normalized PPG cardiac cycle systolic phase peak with respect to dicrotic notch, mean across cardiac cycles |
| PPG_D_RT_NAre_wsDIAPE_DIAN_wrtDIAPE_PPG_D_NAre_wsDIAPE_SYSPP_wrtDIAN_mean | The ratio of the area under the normalized PPG cardiac cycle dicrotic notch with respect to diastolic phase end point, with subtracted diastolic phase end point to the area under the normalized PPG cardiac cycle systolic phase |

| | |
|---|---|
| | peak with respect to dicrotic notch, with subtracted diastolic phase end point, mean across cardiac cycles |
| PPG_NAM_RT_DIAN_wrtZero_PPG_SYSPP_wrtDIAPP_mean | The ratio of the normalized PPG cardiac cycle dicrotic notch with respect to zero to the normalized PPG cardiac cycle systolic phase peak amplitude with respect to diastolic phase peak, mean across cardiac cycles |
| PPG_D_NAvg_SYSPO_wrtDIAPE_mean | Average duration of the normalized PPG cardiac cycle systolic phase onset with respect to diastolic phase end point, mean across cardiac cycles |
| PPG_D_Avg_SYSPP_wrtDIAN_mean | Average duration of the PPG cardiac cycle systolic phase peak with respect to dicrotic notch, mean across cardiac cycles |
| PPG_D_RMS_SYSPP_wrtDIAN_mean | Root mean square duration of the PPG cardiac cycle systolic phase peak with respect to dicrotic notch, mean across cardiac cycles |
| PPG_NAM_RT_SYSPP_wrtDIAN_PPG_SYSPO_wrtZero_std | The ratio of the normalized PPG cardiac cycle systolic phase peak amplitude with respect to dicrotic notch to the normalized PPG cardiac cycle systolic phase onset amplitude with respect to zero, standard deviation across cardiac cycles |
| PPG_D_RT_Are_DIAN_wrtDIAPP_PPG_D_Are_SYSPO_wrtDIAPP_mean | The ratio of the area under the PPG cardiac cycle dicrotic notch with respect to diastolic phase peak to the area under the PPG cardiac cycle systolic phase onset with respect to diastolic phase peak, mean across cardiac cycles |
| PPG_D_RT_NAre_wsDIAPE_DIAN_wrtDIAPE_PPG_D_NAre_wsDIAPE_SYSPP_wrtDIAPE_mean | The ratio of the area under the normalized PPG cardiac cycle dicrotic notch with respect to diastolic phase end point, with subtracted diastolic phase end point to the area under the normalized PPG cardiac cycle systolic phase peak with respect to diastolic phase end point, with subtracted diastolic phase end point, mean across cardiac cycles |
| PPG_D_NMed_SYSPP_wrtDIAPP_mean | Median duration of the normalized PPG cardiac cycle systolic phase peak with respect to diastolic phase peak, mean across cardiac cycles |
| PPG_AM_RT_SD_Apeak_wrtDpeak_PPG_AM_SD_Dpeak_wrtzero_mean | The ratio of the PPG cardiac cycle second derivative Apeak amplitude with respect to |

| | |
|---|---|
| | Dpeak to the PPG cardiac cycle second derivative Dpeak amplitude with respect to zero, mean across cardiac cycles |
| PPG_D_NAvg_DIAN_wrtDIAPE_mean | Average duration of the normalized PPG cardiac cycle dicrotic notch with respect to diastolic phase end point, mean across cardiac cycles |
| PPG_RT_ODP66_W_SRP66_W_mean | The ratio of the PPG cardiac cycle overall decay phase width at 66% of the cardiac cycle to the PPG cardiac cycle systolic rise phase width at 66% of the cardiac cycle, mean across cardiac cycles |
| PPG_D_SD_Bpeak_wrtCpeak_mean | Duration of the PPG cardiac cycle second derivative Bpeak duration with respect to Cpeak, mean across cardiac cycles |
| PPG_AM_RT_SD_Bpeak_wrtEpeak_PPG_AM_SD_Cpeak_wrtzero_std | The ratio of the PPG cardiac cycle second derivative Bpeak amplitude with respect to Epeak to the PPG cardiac cycle second derivative Cpeak amplitude with respect to zero, standard deviation across cardiac cycles |
| PPG_AM_RT_DIAPE_wrtSYSPO_PPG_DIAPE_wrtZero_std | The ratio of the PPG cardiac cycle diastolic phase end point amplitude with respect to systolic phase onset to the PPG diastolic phase end point with respect to zero, standard deviation across cardiac cycles |
| PPG_RT_ODP25_W_SRP25_W_mean | The ratio of the PPG cardiac cycle overall decay phase width at 25% of the cardiac cycle to the PPG cardiac cycle systolic rise phase width at 25% of the cardiac cycle, mean across cardiac cycles |
| PPG_D_NMed_SYSPP_wrtDIAPE_mean | Median duration of the normalized PPG cardiac cycle systolic phase peak with respect to diastolic phase end point, mean across cardiac cycles |
| PPG_AM_RT_SD_Bpeak_wrtCpeak_PPG_AM_SD_Cpeak_wrtzero_std | The ratio of the PPG cardiac cycle second derivative Bpeak amplitude with respect to Cpeak to the PPG cardiac cycle second derivative Cpeak amplitude with respect to zero, standard deviation across cardiac cycles |
| PPG_D_RT_SD_Bpeak_wrtCpeak_PPG_D_SD_Apeak_wrtDpeak_mean | The ratio of the PPG cardiac cycle second derivative Bpeak duration with respect to Cpeak to the PPG cardiac cycle second derivative Apeak duration with respect to Dpeak, mean across cardiac cycles |

| | |
|---|---|
| PPG_AM_RT_FD_Upeak_wrtVpeak_PPG_AM_FD_Wpeak_wrtzero_std | The ratio of the PPG cardiac cycle first derivative Upeak amplitude with respect to Vpeak to the PPG cardiac cycle first derivative W peak amplitude with respect to zero, standard deviation across cardiac cycles |
| PPG_NAM_RT_SYSPP_wrtDIAPP_PPG_SYSPP_wrtDIAN_std | The ratio of the normalized PPG cardiac cycle systolic phase peak amplitude with respect to diastolic phase peak to the normalized PPG cardiac cycle systolic phase peak amplitude with respect to systolic dicrotic notch, standard deviation across cardiac cycles |
| PPG_NAM_RT_DIAPP_wrtDIAN_PPG_SYSPP_wrtDIAN_std | The ratio of the normalized PPG cardiac cycle diastolic phase peak amplitude with respect to dicrotic notch to the normalized PPG cardiac cycle systolic phase peak amplitude with respect to dicrotic notch, standard deviation across cardiac cycles |
| PPG_AM_RT_SD_Dpeak_wrtEpeak_PPG_AM_SD_Bpeak_wrtCpeak_std | The ratio of the PPG cardiac cycle second derivative Dpeak amplitude with respect to Epeak to the PPG cardiac cycle second derivative Bpeak amplitude with respect to Cpeak, standard deviation across cardiac cycles |
| PPG_AM_RT_FD_Upeak_wrtWpeak_PPG_AM_FD_Wpeak_wrtzero_mean | The ratio of the PPG cardiac cycle first derivative Upeak amplitude with respect to Wpeak to the PPG cardiac cycle first derivative Wpeak amplitude with respect to zero, mean across cardiac cycles |
| PPG_AM_RT_DIAPP_wrtDIAN_PPG_DIAPP_wrtZero_std | The ratio of the PPG cardiac cycle diastolic phase peak amplitude with respect to dicrotic notch to the PPG diastolic phase peak with respect to zero, standard deviation across cardiac cycles |
| PPG_AM_RT_SYSPP_wrtDIAPP_PPG_SYSPP_wrtDIAN_std | The ratio of the PPG cardiac cycle systolic phase peak amplitude with respect to diastolic phase peak to the PPG cardiac cycle systolic phase peak amplitude with respect to dicrotic notch , standard deviation across cardiac cycles |
| PPG_AM_RT_DIAPP_wrtDIAN_PPG_SYSPP_wrtDIAN_std | The ratio of the PPG cardiac cycle diastolic phase peak amplitude with respect to dicrotic notch to the PPG cardiac cycle systolic phase peak amplitude with respect to dicrotic notch, standard deviation across cardiac cycles |

| | |
|---|---|
| PPG_D_NMed_SYSPP_wrtDIAN_mean | Median duration of the normalized PPG cardiac cycle systolic phase peak with respect to dicrotic notch, mean across cardiac cycles |
| PPG_D_NAvg_DIAN_wrtDIAPP_mean | Average duration of the normalized PPG cardiac cycle dicrotic notch with respect to diastolic phase peak, mean across cardiac cycles |
| PPG_D_RT_SD_Bpeak_wrtEpeak_PPG_D_SD_Apeak_wrtBpeak_mean | The ratio of the PPG cardiac cycle second derivative Bpeak duration with respect to Epeak to the PPG cardiac cycle second derivative Apeak duration with respect to Bpeak, mean across cardiac cycles |
| PPG_AM_RT_SD_Apeak_wrtCpeak_PPG_AM_SD_Dpeak_wrtzero_mean | The ratio of the PPG cardiac cycle second derivative Apeak amplitude with respect to Cpeak to the PPG cardiac cycle second derivative Dpeak amplitude with respect to zero, mean across cardiac cycles |
| PPG_D_NAre_DIAN_wrtDIAPE_mean | Area under the normalized PPG cardiac cycle dicrotic notch with respect to diastolic phase end point, mean across cardiac cycles |
| PPG_AM_FD_Upeak_wrtWpeak_mean | PPG cardiac cycle first derivative Upeak amplitude with respect to Wpeak, mean across cardiac cycles |
| PPG_D_RT_SD_Bpeak_wrtDpeak_PPG_D_SD_Apeak_wrtBpeak_mean | The ratio of the PPG cardiac cycle second derivative Bpeak duration with respect to Dpeak to the PPG cardiac cycle second derivative Apeak duration with respect to Bpeak, mean across cardiac cycles |
| PPG_D_NRMS_DIAN_wrtDIAPP_mean | Root mean square duration of the normalized PPG cardiac cycle dicrotic notch with respect to diastolic phase peak, mean across cardiac cycles |
| PPG_RT_ODP25_W_SRP25_ODP25_W_mean | The ratio of the PPG cardiac cycle overall decay phase width at 25% of the cardiac cycle to the sum of the PPG cardiac cycle systolic rise phase width and overall decay phase width at 25% of the cardiac cycle, mean across cardiac cycles |
| PPG_AM_RT_DIAPE_wrtSYSPO_PPG_DIAPE_wrtZero_mean | The ratio of the PPG cardiac cycle diastolic phase end point amplitude with respect to systolic phase onset to the PPG diastolic phase end point with respect to zero, mean across cardiac cycles |
| PPG_AM_RT_DIAPP_wrtDIAPE_PPG_SYSPP_wrtDIAN_mean | The ratio of the PPG cardiac cycle diastolic phase peak amplitude with respect to diastolic |

| | |
|---|---|
| | phase end point to the PPG cardiac cycle systolic phase peak amplitude with respect to dicrotic notch, mean across cardiac cycles |
| PPG_RT_SRP25_W_SRP25_ODP25_W_mean | The ratio of the PPG cardiac cycle systolic rise phase width at 25% of the cardiac cycle to the sum of the PPG cardiac cycle systolic rise phase width and overall decay phase width at 25% of the cardiac cycle, mean across cardiac cycles |
| PPG_AM_RT_SD_Bpeak_wrtEpeak_PPG_AM_SD_Apeak_wrtBpeak_mean | The ratio of the PPG cardiac cycle second derivative Bpeak amplitude with respect to Epeak to the PPG cardiac cycle second derivative Apeak amplitude with respect to Bpeak, mean across cardiac cycles |
| PPG_AM_RT_SD_Apeak_wrtEpeak_PPG_AM_SD_Apeak_wrtBpeak_mean | The ratio of the PPG cardiac cycle second derivative Apeak amplitude with respect to Epeak to the PPG cardiac cycle second derivative Apeak amplitude with respect to Bpeak, mean across cardiac cycles |
| PPG_D_NAvg_wsDIAPE_DIAN_wrtDIAPE_mean | Average duration of the normalized PPG cardiac cycle dicrotic notch with respect to diastolic phase end point, with subtracted diastolic phase end point, mean across cardiac cycles |
| PPG_D_NAre_wsDIAPE_DIAN_wrtDIAPE_mean | Area under the normalized PPG cardiac cycle dicrotic notch with respect to diastolic phase end point, with subtracted diastolic phase end point, mean across cardiac cycles |
| PPG_D_Med_SYSPO_wrtDIAPP_mean | Median duration of the PPG cardiac cycle systolic phase onset with respect to diastolic phase peak, mean across cardiac cycles |
| PPG_D_Med_SYSPO_wrtSYSPP_mean | Median duration of the PPG cardiac cycle systolic phase onset with respect to systolic phase peak, mean across cardiac cycles |
| PPG_D_SD_Apeak_wrtBpeak_std | Duration of the PPG cardiac cycle second derivative Apeak duration with respect to Bpeak, standard deviation across cardiac cycles |
| PPG_AM_RT_SD_Cpeak_wrtDpeak_PPG_AM_SD_Bpeak_wrtDpeak_mean | The ratio of the PPG cardiac cycle second derivative Cpeak amplitude with respect to Dpeak to the PPG cardiac cycle second derivative Bpeak amplitude with respect to Dpeak, mean across cardiac cycles |
| PPG_NAM_RT_DIAPP_wrtZero_PPG_SYSPP_wrtDIAN_mean | The ratio of the normalized PPG cardiac cycle diastolic phase peak with respect to zero to the normalized PPG cardiac cycle systolic phase |

| | |
|---|---|
| | peak amplitude with respect to dicrotic notch, mean across cardiac cycles |
| PPG_D_RT_Are_DIAN_wrtDIAPP_PPG_D_Are_SYSPO_wrtDIAPE_mean | The ratio of the area under the PPG cardiac cycle dicrotic notch with respect to diastolic phase peak to the area under the PPG cardiac cycle systolic phase onset with respect to diastolic phase end point, mean across cardiac cycles |
| PPG_D_RT_Are_wsDIAPE_DIAN_wrtDIAPP_PPG_D_Are_wsDIAPE_SYSPO_wrtDIAPE_mean | The ratio of the area under the PPG cardiac cycle dicrotic notch with respect to diastolic phase peak, with subtracted diastolic phase end point to the area under the PPG cardiac cycle systolic phase onset with respect to diastolic phase end point, with subtracted diastolic phase end point, mean across cardiac cycles |
| PPG_D_RT_SD_Apeak_wrtDpeak_PPG_D_SD_Apeak_wrtBpeak_mean | The ratio of the PPG cardiac cycle second derivative Apeak duration with respect to Dpeak to the PPG cardiac cycle second derivative Apeak duration with respect to Bpeak, mean across cardiac cycles |
| PPG_W_SRP10_ODP10_mean | Sum of the PPG cardiac cycle systolic rise phase width and overall decay phase width at 10% of the cardiac cycle, mean across cardiac cycles |
| PPG_AM_RT_FD_Vpeak_wrtWpeak_PPG_AM_FD_Wpeak_wrtzero_mean | The ratio of the PPG cardiac cycle first derivative Vpeak amplitude with respect to Wpeak to the PPG cardiac cycle first derivative Wpeak amplitude with respect to zero, mean across cardiac cycles |
| PPG_AM_RT_SD_Bpeak_wrtDpeak_PPG_AM_SD_Bpeak_wrtCpeak_std | The ratio of the PPG cardiac cycle second derivative Bpeak amplitude with respect to Dpeak to the PPG cardiac cycle second derivative Bpeak amplitude with respect to Cpeak, standard deviation across cardiac cycles |
| PPG_AM_RT_SD_Cpeak_wrtDpeak_PPG_AM_SD_Bpeak_wrtCpeak_std | The ratio of the PPG cardiac cycle second derivative Cpeak amplitude with respect to Dpeak to the PPG cardiac cycle second derivative Bpeak amplitude with respect to Cpeak, standard deviation across cardiac cycles |
| PPG_D_NAvg_SYSPO_wrtDIAPP_mean | Average duration of the normalized PPG cardiac cycle systolic phase onset with respect to diastolic phase peak, mean across cardiac cycles |

| | |
|---|---|
| PPG_D_Are_wsDIAPE_SYSPO_wrtDIAPE_mean | Area under the PPG cardiac cycle systolic phase onset with respect to diastolic phase end point, with subtracted diastolic phase end point, mean across cardiac cycles |
| PPG_AM_FD_Vpeak_wrtWpeak_mean | PPG cardiac cycle first derivative Vpeak amplitude with respect to Wpeak, mean across cardiac cycles |
| PPG_D_NMed_SYSPO_wrtDIAPE_std | Median duration of the normalized PPG cardiac cycle systolic phase onset with respect to diastolic phase end point, standard deviation across cardiac cycles |
| PPG_NAM_RT_DIAPP_wrtDIAPE_PPG_SYSPP_wrtDIAN_mean | The ratio of the normalized PPG cardiac cycle diastolic phase peak with respect to diastolic phase end point to the normalized PPG cardiac cycle systolic phase peak amplitude with respect to dicrotic notch, mean across cardiac cycles |
| PPG_D_RT_SD_Cpeak_wrtEpeak_PPG_D_SD_Bpeak_wrtEpeak_mean | The ratio of the PPG cardiac cycle second derivative Cpeak duration with respect to Epeak to the PPG cardiac cycle second derivative Dpeak duration with respect to Epeak, mean across cardiac cycles |
| PPG_D_NMed_SYSPO_wrtDIAPP_mean | Median duration of the normalized PPG cardiac cycle systolic phase onset with respect to diastolic phase peak, mean across cardiac cycles |
| PPG_D_Med_SYSPP_wrtDIAN_mean | Median duration of the PPG cardiac cycle systolic phase peak with respect to dicrotic notch, mean across cardiac cycles |
| PPG_D_SYSPP_wrtDIAPP_std | Duration of the PPG cardiac cycle systolic phase peak with respect to diastolic phase peak, standard deviation across cardiac cycles |
| PPG_NAM_RT_DIAPP_wrtDIAN_PPG_SYSPO_wrtZero_std | The ratio of the normalized PPG cardiac cycle diastolic phase peak amplitude with respect to dicrotic notch to the normalized PPG cardiac cycle systolic phase onset amplitude with respect to zero, standard deviation across cardiac cycles |
| PPG_AM_RT_DIAPP_wrtSYSPO_PPG_SYSPP_wrtDIAN_mean | The ratio of the PPG cardiac cycle diastolic phase peak amplitude with respect to systolic phase onset to the PPG cardiac cycle systolic phase peak amplitude with respect to dicrotic notch, mean across cardiac cycles |

| | |
|---|---|
| PPG_RT_ODP25_W_SRP25_W_std | The ratio of the PPG cardiac cycle overall decay phase width at 25% of the cardiac cycle to the PPG cardiac cycle systolic rise phase width at 25% of the cardiac cycle, standard deviation across cardiac cycles |
| PPG_D_RT_NAre_SYSPP_wrtDIAPP_PPG_D_NAre_SYSPO_wrtDIAPE_mean | The ratio of the area under the normalized PPG cardiac cycle systolic phase peak with respect to diastolic phase peak to the area under the normalized PPG cardiac cycle systolic phase onset with respect to diastolic phase end point, mean across cardiac cycles |
| PPG_Slope_DIAN_DIAPP_mean | Mean across cardiac cycles of the slope between the dicrotic notch and diastolic phase peak of the PPG cardiac cycle |
| PPG_AM_RT_DIAPP_wrtDIAN_PPG_SYSPO_wrtZero_std | The ratio of the PPG cardiac cycle diastolic phase peak amplitude with respect to dicrotic notch to the PPG cardiac cycle systolic phase onset amplitude with respect to zero, standard deviation across cardiac cycles |
| PPG_D_SYSPO_wrtDIAPE_mean | Duration of the PPG cardiac cycle systolic phase onset with respect to diastolic phase end point, mean across cardiac cycles |
| PPG_AM_RT_SD_Apeak_wrtEpeak_PPG_AM_SD_Dpeak_wrtzero_mean | The ratio of the PPG cardiac cycle second derivative Apeak amplitude with respect to Epeak to the PPG cardiac cycle second derivative Dpeak amplitude with respect to zero, mean across cardiac cycles |
| PPG_AM_RT_DIAPP_wrtDIAN_PPG_DIAN_wrtDIAPE_mean | The ratio of the PPG cardiac cycle diastolic phase peak amplitude with respect to dicrotic notch to the PPG cardiac cycle dicrotic notch amplitude with respect to diastolic phase end point, mean across cardiac cycles |
| PPG_AM_RT_DIAPP_wrtDIAPE_PPG_DIAN_wrtDIAPE_mean | The ratio of the PPG cardiac cycle diastolic phase peak amplitude with respect to diastolic phase end point to the PPG cardiac cycle dicrotic notch amplitude with respect to diastolic phase end point, mean across cardiac cycles |
| PPG_NAM_RT_DIAPP_wrtSYSPO_PPG_SYSPP_wrtDIAN_mean | The ratio of the normalized PPG cardiac cycle diastolic phase peak amplitude with respect to systolic phase onset to the normalized PPG cardiac cycle systolic phase peak amplitude with respect to dicrotic notch, mean across cardiac cycles |

| | |
|---|---|
| PPG_AM_RT_SD_Apeak_wrtCpeak_PPG_AM_SD_Apeak_wrtzero_mean | The ratio of the PPG cardiac cycle second derivative Apeak amplitude with respect to Cpeak to the PPG cardiac cycle second derivative Apeak amplitude with respect to zero, mean across cardiac cycles |
| PPG_AM_RT_SD_Cpeak_wrtzero_PPG_AM_SD_Apeak_wrtzero_mean | The ratio of the PPG cardiac cycle second derivative Cpeak amplitude with respect to zero to the PPG cardiac cycle second derivative Apeak amplitude with respect to zero, mean across cardiac cycles |
| PPG_D_RT_SYSPP_wrtDIAPP_PPG_SYSPP_wrtDIAN_std | The ratio of the PPG cardiac cycle systolic phase peak duration with respect to diastolic phase peak to the PPG cardiac cycle systolic phase peak duration with respect to dicrotic notch, standard deviation across cardiac cycles |
| PPG_D_RT_Are_wsDIAPE_DIAN_wrtDIAPP_PPG_D_Are_wsDIAPE_SYSPP_wrtDIAN_std | The ratio of the area under the PPG cardiac cycle dicrotic notch with respect to diastolic phase end peak, with subtracted diastolic phase end point to the area under the PPG cardiac cycle systolic phase peak with respect to dicrotic notch, with subtracted diastolic phase end point, standard deviation across cardiac cycles |
| PPG_D_RT_Are_wsDIAPE_DIAN_wrtDIAPE_PPG_D_Are_wsDIAPE_SYSPP_wrtDIAN_mean | The ratio of the area under the PPG cardiac cycle dicrotic notch with respect to diastolic phase end point, with subtracted diastolic phase end point to the area under the PPG cardiac cycle systolic phase peak with respect to dicrotic notch, with subtracted diastolic phase end point, mean across cardiac cycles |
| PPG_D_RT_DIAPP_wrtDIAPE_PPG_SYSPP_wrtDIAPE_std | The ratio of the PPG cardiac cycle diastolic phase peak duration with respect to diastolic phase end point to the PPG cardiac cycle systolic phase peak duration with respect to diastolic phase end point, standard deviation across cardiac cycles |
| PPG_D_NAvg_SYSPO_wrtDIAN_mean | Average duration of the normalized PPG cardiac cycle systolic phase onset with respect to dicrotic notch, mean across cardiac cycles |
| PPG_D_RT_NAre_wsDIAPE_SYSPP_wrtDIAPP_PPG_D_NAre_wsDIAPE_SYSPO_wrtDIAPE_mean | The ratio of the area under the normalized PPG cardiac cycle systolic phase peak with respect to diastolic phase peak, with subtracted diastolic phase end point to the area under the |

| | |
|---|---|
| | normalized PPG cardiac cycle systolic phase onset with respect to diastolic phase end point, with subtracted diastolic phase end point, mean across cardiac cycles |
| PPG_AM_RT_FD_Upeak_wrtWpeak_PPG_AM_FD_Wpeak_wrtzero_std | The ratio of the PPG cardiac cycle first derivative Upeak amplitude with respect to Wpeak to the PPG cardiac cycle first derivative Wpeak amplitude with respect to zero, standard deviation across cardiac cycles |
| PPG_AM_RT_SD_Bpeak_wrtDpeak_PPG_AM_SD_Epeak_wrtzero_mean | The ratio of the PPG cardiac cycle second derivative Bpeak amplitude with respect to Dpeak to second derviative of PPG cardiac cycle Epeak amplitude with respect to zero, mean across cardiac cycles |
| PPG_D_NAre_DIAN_wrtDIAPP_std | Area under the normalized PPG cardiac cycle dicrotic notch with respect to diastolic phase peak, standard deviation across cardiac cycles |
| PPG_D_RT_Are_wsDIAPE_SYSPP_wrtDIAPP_PPG_D_Are_wsDIAPE_SYSPP_wrtDIAN_std | The ratio of the area under the PPG cardiac cycle systolic phase peak with respect to diastolic phase peak, with subtracted diastolic phase end point to the area under the PPG cardiac cycle systolic phase peak with respect to dicrotic notch, with subtracted diastolic phase end point, standard deviation across cardiac cycles |
| PPG_NAM_RT_DIAPP_wrtDIAN_PPG_DIAPP_wrtDIAPE_mean | The ratio of the normalized PPG cardiac cycle diastolic phase peak amplitude with respect to dicrotic notch to the normalized PPG cardiac cycle diastolic phase peak amplitude with respect to diastolic phase end point, mean across cardiac cycles |
| PPG_D_RT_DIAN_wrtDIAPP_PPG_SYSPP_wrtDIAN_std | The ratio of the PPG cardiac cycle dicrotic notch duration with respect to diastolic phase peak to the PPG cardiac cycle systolic phase peak duration with respect to dicrotic notch, standard deviation across cardiac cycles |
| PPG_D_Are_SYSPO_wrtDIAPE_mean | Area under the PPG cardiac cycle systolic phase onset with respect to diastolic phase end point, mean across cardiac cycles |
| PPG_D_RT_SD_Cpeak_wrtDpeak_PPG_D_SD_Bpeak_wrtEpeak_mean | The ratio of the PPG cardiac cycle second derivative Cpeak duration with respect to Dpeak to the PPG cardiac cycle second derivative Bpeak duration with respect to Epeak, mean across cardiac cycles |

| | |
|---|---|
| PPG_AM_RT_DIAPP_wrtDIAN_PPG_DIAPP_wrtZero_mean | The ratio of the PPG cardiac cycle diastolic phase peak amplitude with respect to dicrotic notch to the PPG diastolic phase peak with respect to zero, mean across cardiac cycles |
| PPG_D_NRMS_SYSPP_wrtDIAPP_mean | Root mean square duration of the normalized PPG cardiac cycle systolic phase peak with respect to diastolic phase peak, mean across cardiac cycles |
| PPG_D_RT_SD_Cpeak_wrtEpeak_PPG_D_SD_Apeak_wrtCpeak_std | The ratio of the PPG cardiac cycle second derivative Cpeak duration with respect to Epeak to the PPG cardiac cycle second derivative Apeak duration with respect to Cpeak, standard deviation across cardiac cycles |
| PPG_D_RT_Are_DIAN_wrtDIAPP_PPG_D_Are_SYSPP_wrtDIAN_std | The ratio of the area under the PPG cardiac cycle dicrotic notch with respect to diastolic phase peak to the area under the PPG cardiac cycle systolic phase peak with respect to dicrotic notch, standard deviation across cardiac cycles |
| PPG_D_RT_Are_SYSPP_wrtDIAPP_PPG_D_Are_SYSPP_wrtDIAN_std | The ratio of the area under the PPG cardiac cycle systolic phase peak with respect to diastolic phase peak to the area under the PPG cardiac cycle systolic phase peak with respect to dicrotic notch, standard deviation across cardiac cycles |
| PPG_NAM_RT_DIAPP_wrtDIAN_PPG_DIAN_wrtZero_mean | The ratio of the normalized PPG cardiac cycle diastolic phase peak amplitude with respect to dicrotic notch to the normalized PPG cardiac cycle dicrotic notch amplitude with respect to zero, mean across cardiac cycles |
| PPG_NAM_RT_DIAPP_wrtZero_PPG_DIAN_wrtZero_mean | The ratio of the normalized PPG cardiac cycle diastolic phase peak amplitude with respect to zero to the normalized PPG cardiac cycle dicrotic notch amplitude with respect to zero, mean across cardiac cycles |
| PPG_D_RT_NAre_wsDIAPE_DIAN_wrtDIAPP_PPG_D_NAre_wsDIAPE_SYSPP_wrtDIAN_std | The ratio of the area under the normalized PPG cardiac cycle dicrotic notch with respect to diastolic phase peak, with subtracted diastolic phase end point to the area under the normalized PPG cardiac cycle systolic phase peak with respect to dicrotic notch, with subtracted diastolic phase end point, standard deviation across cardiac cycles |

| | |
|---|---|
| PPG_D_NAvg_SYSPP_wrtDIAPP_mean | Average duration of the normalized PPG cardiac cycle systolic phase peak with respect to diastolic phase peak, mean across cardiac cycles |
| PPG_D_RMS_SYSPP_wrtDIAPP_std | Root mean square duration of the PPG cardiac cycle systolic phase peak with respect to diastolic phase peak , standard deviation across cardiac cycles |
| PPG_D_SD_Bpeak_wrtEpeak_mean | Duration of the PPG cardiac cycle second derivative Bpeak duration with respect to Epeak, mean across cardiac cycles |
| PPG_AM_RT_SD_Apeak_wrtBpeak_PPG_AM_SD_Cpeak_wrtzero_std | The ratio of the PPG cardiac cycle second derivative Apeak amplitude with respect to Bpeak to the PPG cardiac cycle second derivative Cpeak amplitude with respect to zero, standard deviation across cardiac cycles |
| PPG_D_Avg_wsDIAPE_SYSPP_wrtDIAN_mean | Average duration of the PPG cardiac cycle systolic phase peak with respect to dicrotic notch, with subtracted diastolic phase end point, mean across cardiac cycles |
| PPG_AM_RT_SD_Bpeak_wrtDpeak_PPG_AM_SD_Cpeak_wrtzero_mean | The ratio of the PPG cardiac cycle second derivative Bpeak amplitude with respect to Dpeak to the PPG cardiac cycle second derivative Cpeak amplitude with respect to zero, mean across cardiac cycles |
| PPG_AM_RT_DIAPP_wrtDIAN_PPG_SYSPO_wrtZero_mean | The ratio of the PPG cardiac cycle diastolic phase peak amplitude with respect to dicrotic notch to the PPG cardiac cycle systolic phase onset amplitude with respect to zero, mean across cardiac cycles |
| PPG_NAM_RT_DIAPP_wrtSYSPO_PPG_DIAN_wrtZero_mean | The ratio of the normalized PPG cardiac cycle diastolic phase peak amplitude with respect to systolic phase onset to the normalized PPG cardiac cycle dicrotic notch amplitude with respect to zero, mean across cardiac cycles |
| PPG_D_NAre_wsDIAPE_DIAN_wrtDIAPP_std | Area under the normalized PPG cardiac cycle dicrotic notch with respect to diastolic phase peak, with subtracted diastolic phase end point, standard deviation across cardiac cycles |
| PPG_D_RT_SYSPP_wrtDIAPP_PPG_SYSPO_wrtDIAPP_std | The ratio of the PPG cardiac cycle systolic phase peak duration with respect to diastolic phase peak to the PPG cardiac cycle systolic phase onset duration with respect to diastolic phase peak, standard deviation across cardiac cycles |

### Detailed descriptions of the ABP features (n=217) used to develop the ABPAge aging clock

| ABP Features | Descriptions |
|---|---|
| ABP_pkamp_std | Standard deviation across cardiac cycles of the amplitude of the first peak observed in the FFT of the ABP cardiac cycle |
| ABP_pkamp_mean | Mean across cardiac cycles of the amplitude of the first peak observed in the FFT of the ABP cardiac cycle |
| ABP_D_Avg_wsDIAPE_SYSPO_wrtSYSPP_mean | Average duration of the ABP cardiac cycle systolic phase onset with respect to systolic phase peak, with subtracted diastolic phase end point, mean across cardiac cycles |
| ABP_D_Avg_wsDIAPE_SYSPP_wrtDIAN_std | Average duration of the ABP cardiac cycle systolic phase peak with respect to dicrotic notch, with subtracted diastolic phase end point, standard deviation across cardiac cycles |
| ABP_D_Avg_wsDIAPE_SYSPP_wrtDIAN_mean | Average duration of the ABP cardiac cycle systolic phase peak with respect to dicrotic notch, with subtracted diastolic phase end point, mean across cardiac cycles |
| ABP_D_Avg_wsDIAPE_SYSPO_wrtDIAPP_std | Average duration of the ABP cardiac cycle systolic phase onset with respect to diastolic phase peak, with subtracted diastolic phase end point, standard deviation across cardiac cycles |
| ABP_AM_RT_SYSPP_wrtZero_ABP_SYSPO_wrtZero_mean | The ratio of the ABP cardiac cycle systolic phase peak amplitude with respect to zero to the ABP cardiac cycle systolic phase onset amplitude with respect to zero, mean across cardiac cycles |
| ABP_AM_RT_SYSPP_wrtSYSPO_ABP_SYSPO_wrtZero_mean | The ratio of the ABP cardiac cycle systolic phase peak amplitude with respect to systolic phase onset to the ABP cardiac cycle systolic phase onset amplitude with respect to zero, mean across cardiac cycles |
| ABP_D_Avg_wsDIAPE_SYSPO_wrtDIAN_mean | Average duration of the ABP cardiac cycle systolic phase onset with respect to dicrotic notch, with subtracted diastolic phase end point, mean across cardiac cycles |
| ABP_D_Avg_wsDIAPE_SYSPO_wrtDIAPE_mean | Average duration of the ABP cardiac cycle systolic phase onset with respect to diastolic phase end point, with subtracted diastolic phase end point, mean across cardiac cycles |

| | |
|---|---|
| ABP_D_Avg_wsDIAPE_SYSPO_wrtDIAPP_mean | Average duration of the ABP cardiac cycle systolic phase onset with respect to diastolic phase peak, with subtracted diastolic phase end point, mean across cardiac cycles |
| ABP_D_Avg_wsDIAPE_SYSPO_wrtSYSPP_std | Average duration of the ABP cardiac cycle systolic phase onset with respect to systolic phase peak, with subtracted diastolic phase end point, standard deviation across cardiac cycles |
| ABP_AM_RT_SYSPP_wrtDIAPP_ABP_SYSPO_wrtZero_mean | The ratio of the ABP cardiac cycle systolic phase peak amplitude with respect to diastolic phase peak to the ABP cardiac cycle systolic phase onset amplitude with respect to zero, mean across cardiac cycles |
| ABP_AM_RT_SYSPP_wrtDIAPE_ABP_SYSPO_wrtZero_mean | The ratio of the ABP cardiac cycle systolic phase peak amplitude with respect to diastolic phase end point to the ABP cardiac cycle systolic phase onset amplitude with respect to zero, mean across cardiac cycles |
| ABP_D_Avg_wsDIAPE_SYSPO_wrtDIAPE_std | Average duration of the ABP cardiac cycle systolic phase onset with respect to diastolic phase end point, with subtracted diastolic phase end point, standard deviation across cardiac cycles |
| ABP_D_FD_Upeak_wrtVpeak_mean | Duration of the ABP cardiac cycle first derivative Upeak with respect to Vpeak, mean across cardiac cycles |
| ABP_AM_RT_SYSPP_wrtSYSPO_ABP_SYSPP_wrtZero_mean | The ratio of the ABP cardiac cycle systolic phase peak amplitude with respect to systolic phase onset to the ABP cardiac cycle systolic phase peak amplitude with respect to zero, mean across cardiac cycles |
| ABP_AM_SD_Dpeak_wrtEpeak_std | ABP cardiac cycle second derivative Dpeak amplitude with respect to Epeak, standard deviation across cardiac cycles |
| ABP_D_RMS_SYSPP_wrtDIAN_std | Root mean square duration of the ABP cardiac cycle systolic phase peak with respect to dicrotic notch, standard deviation across cardiac cycles |
| ABP_AM_RT_DIAPE_wrtZero_ABP_SYSPP_wrtSYSPO_mean | The ratio of the ABP cardiac cycle diastolic phase end point amplitude with respect to zero to the ABP cardiac cycle systolic phase peak amplitude with respect to systolic phase onset, mean across cardiac cycles |
| ABP_AM_RT_DIAPP_wrtZero_ABP_DIAN_wrtZero_mean | The ratio of the ABP cardiac cycle diastolic phase peak amplitude with respect to zero to the ABP |

| | |
|---|---|
| | cardiac cycle dicrotic notch amplitude with respect to zero, mean across cardiac cycles |
| ABP_AM_RT_DIAPP_wrtDIAN_ABP_DIAN_wrtZero_mean | The ratio of the ABP cardiac cycle diastolic phase peak amplitude with respect to dicrotic notch to the ABP cardiac cycle dicrotic notch amplitude with respect to zero, mean across cardiac cycles |
| ABP_AM_SD_Epeak_wrtzero_std | ABP cardiac cycle second derivative Epeak amplitude with respect to zero, standard deviation across cardiac cycles |
| ABP_AM_FD_Vpeak_wrtzero_mean | ABP cardiac cycle first derivative Vpeak amplitude with respect to zero, mean across cardiac cycles |
| ABP_AM_RT_DIAPE_wrtZero_ABP_SYSPP_wrtZero_mean | The ratio of the ABP cardiac cycle diastolic phase end point amplitude with respect to zero to the ABP cardiac cycle systolic phase peak amplitude with respect to zero, mean across cardiac cycles |
| ABP_AM_RT_SYSPP_wrtDIAPE_ABP_SYSPP_wrtZero_mean | The ratio of the ABP cardiac cycle systolic phase peak amplitude with respect to diastolic phase end point to the ABP cardiac cycle systolic phase peak amplitude with respect to zero, mean across cardiac cycles |
| ABP_D_Avg_wsDIAPE_SYSPP_wrtDIAPE_std | Average duration of the ABP cardiac cycle systolic phase peak with respect to diastolic phase end point, with subtracted diastolic phase end point, standard deviation across cardiac cycles |
| ABP_D_Avg_wsDIAPE_SYSPO_wrtDIAN_std | Average duration of the ABP cardiac cycle systolic phase onset with respect to dicrotic notch, with subtracted diastolic phase end point, standard deviation across cardiac cycles |
| ABP_AM_SYSPP_wrtSYSPO_mean | ABP cardiac cycle systolic phase peak amplitude with respect to systolic phase onset, mean cardiac cycles |
| ABP_D_Avg_wsDIAPE_SYSPP_wrtDIAPP_std | Average duration of the ABP cardiac cycle systolic phase peak with respect to diastolic phase peak, with subtracted diastolic phase end point, standard deviation across cardiac cycles |
| ABP_D_Avg_wsDIAPE_SYSPP_wrtDIAPE_mean | Average duration of the ABP cardiac cycle systolic phase peak with respect to diastolic phase end point, with subtracted diastolic phase end point, mean across cardiac cycles |
| ABP_AM_RT_DIAPP_wrtDIAN_ABP_SYSPO_wrtZero_mean | The ratio of the ABP cardiac cycle diastolic phase peak amplitude with respect to dicrotic notch to the ABP cardiac cycle systolic phase onset amplitude with respect to zero, mean across cardiac cycles |

| | |
|---|---|
| ABP_AM_RT_DIAPP_wrtZero_ABP_DIAN_wrtDIAPE_mean | The ratio of the ABP cardiac cycle diastolic phase peak amplitude with respect to zero to the ABP cardiac cycle dicrotic notch amplitude with respect to diastolic phase end point, mean across cardiac cycles |
| ABP_AM_RT_DIAPP_wrtZero_ABP_SYSPP_wrtSYSPO_mean | The ratio of the ABP cardiac cycle diastolic phase peak amplitude with respect to zero to the ABP cardiac cycle systolic phase peak amplitude with respect to systolic phase onset, mean across cardiac cycles |
| ABP_AM_SYSPP_wrtDIAPP_mean | ABP/PPG cardiac cycle systolic phase peak amplitude with respect to diastolic phase peak, mean across cardiac cycles |
| ABP_D_Avg_SYSPP_wrtDIAN_std | Average duration of the ABP cardiac cycle systolic phase peak with respect to dicrotic notch, standard deviation across cardiac cycles |
| ABP_AM_SYSPP_wrtSYSPO_std | ABP cardiac cycle systolic phase peak amplitude with respect to systolic phase onset, standard deviation across cardiac cycles |
| ABP_AM_RT_DIAPE_wrtZero_ABP_SYSPP_wrtDIAPE_mean | The ratio of the ABP cardiac cycle diastolic phase end point amplitude with respect to zero to the ABP cardiac cycle systolic phase peak amplitude with respect to diastolic phase end point, mean across cardiac cycles |
| ABP_AM_RT_DIAPP_wrtZero_ABP_DIAN_wrtSYSPO_mean | The ratio of the ABP cardiac cycle diastolic phase peak amplitude with respect to zero to the ABP cardiac cycle dicrotic notch amplitude with respect to systolic phase onset, mean across cardiac cycles |
| ABP_AM_RT_SD_Cpeak_wrtDpeak_ABP_AM_SD_Cpeak_wrtzero_mean | The ratio of the ABP cardiac cycle second derivative Cpeak amplitude with respect to D peak to the ABP cardiac cycle second derivative Cpeak amplitude with respect to zero, mean across cardiac cycles |
| ABP_AM_RT_SD_Dpeak_wrtzero_ABP_AM_SD_Cpeak_wrtzero_mean | The ratio of the ABP cardiac cycle second derivative Dpeak amplitude with respect to zero to the ABP cardiac cycle second derivative Cpeak amplitude with respect to zero, mean across cardiac cycles |
| ABP_AM_SD_Cpeak_wrtEpeak_mean | ABP cardiac cycle second derivative Cpeak amplitude with respect to Epeak, mean across cardiac cycles |
| ABP_AM_RT_SD_Bpeak_wrtCpeak_ABP_AM_SD_Cpeak_wrtzero_mean | The ratio of the ABP cardiac cycle second derivative Bpeak amplitude with respect to Cpeak to the ABP cardiac cycle second derivative Cpeak |

| | |
|---|---|
| | amplitude with respect to zero, mean across cardiac cycles |
| ABP_D_RMS_SYSPP_wrtDIAN_mean | Root mean square duration of the ABP cardiac cycle systolic phase peak with respect to dicrotic notch, mean across cardiac cycles |
| ABP_AM_SYSPP_wrtDIAPE_std | ABP cardiac cycle systolic phase peak amplitude with respect to diastolic phase end point, standard deviation across cardiac cycles |
| ABP_D_Med_SYSPP_wrtDIAN_std | Median duration of the ABP cardiac cycle systolic phase peak with respect to dicrotic notch, standard deviation across cardiac cycles |
| ABP_AM_RT_DIAPE_wrtZero_ABP_DIAN_wrtDIAPE_mean | The ratio of the ABP cardiac cycle diastolic phase end point amplitude with respect to zero to the ABP cardiac cycle dicrotic notch amplitude with respect to diastolic phase end point, mean across cardiac cycles |
| ABP_AM_RT_SD_Dpeak_wrtEpeak_ABP_AM_SD_Cpeak_wrtzero_mean | The ratio of the ABP cardiac cycle second derivative Dpeak amplitude with respect to Epeak to the ABP cardiac cycle second derivative Cpeak amplitude with respect to zero, mean across cardiac cycles |
| ABP_AM_RT_DIAN_wrtSYSPO_ABP_DIAN_wrtZero_mean | The ratio of the ABP cardiac cycle dicrotic notch amplitude with respect to systolic phase onset to the ABP dicrotic notch amplitude with respect to zero, mean across cardiac cycles |
| ABP_AM_RT_DIAN_wrtDIAPE_ABP_DIAN_wrtZero_mean | The ratio of the ABP cardiac cycle dicrotic notch amplitude with respect to diastolic phase end point to the ABP dicrotic notch amplitude with respect to zero, mean across cardiac cycles |
| ABP_AM_RT_DIAPE_wrtZero_ABP_DIAN_wrtZero_mean | The ratio of the ABP cardiac cycle diastolic phase end point amplitude with respect to zero to the ABP cardiac cycle dicrotic notch amplitude with respect to zero, mean across cardiac cycles |
| ABP_D_Avg_wsDIAPE_SYSPP_wrtDIAPP_mean | Average duration of the ABP cardiac cycle systolic phase peak with respect to diastolic phase peak, with subtracted diastolic phase end point, mean across cardiac cycles |
| ABP_AM_RT_SD_Dpeak_wrtEpeak_ABP_AM_SD_Cpeak_wrtEpeak_std | The ratio of the ABP cardiac cycle second derivative Dpeak amplitude with respect to Epeak to the ABP cardiac cycle second derivative Cpeak amplitude with respect to Epeak, standard deviation across cardiac cycles |
| ABP_AM_RT_DIAN_wrtDIAPE_ABP_SYSPO_wrtZero_mean | The ratio of the ABP cardiac cycle dicrotic notch amplitude with respect to diastolic phase end |

| | |
|---|---|
| | point to the ABP cardiac cycle systolic phase onset amplitude with respect to zero, mean across cardiac cycles |
| ABP_AM_RT_SD_Apeak_wrtCpeak_ABP_AM_SD_Cpeak_wrtzero_mean | The ratio of the ABP cardiac cycle second derivative Apeak amplitude with respect to Cpeak to the ABP cardiac cycle second derivative Cpeak amplitude with respect to zero, mean across cardiac cycles |
| ABP_AM_DIAPP_wrtDIAN_std | ABP cardiac cycle diastolic phase peak amplitude with respect to dicrotic notch, standard deviation across cardiac cycles |
| ABP_AM_SYSPP_wrtDIAPE_mean | ABP cardiac cycle systolic phase peak amplitude with respect to diastolic phase end point, mean across cardiac cycles |
| ABP_AM_RT_SD_Apeak_wrtEpeak_ABP_AM_SD_Cpeak_wrtzero_mean | The ratio of the ABP cardiac cycle second derivative Apeak amplitude with respect to Epeak to the ABP cardiac cycle second derivative Cpeak amplitude with respect to zero, mean across cardiac cycles |
| ABP_AM_RT_DIAPP_wrtDIAN_ABP_DIAPP_wrtZero_mean | The ratio of the ABP cardiac cycle diastolic phase peak amplitude with respect to dicrotic notch to the ABP diastolic phase peak with respect to zero, mean across cardiac cycles |
| ABP_AM_RT_SD_Apeak_wrtDpeak_ABP_AM_SD_Cpeak_wrtzero_mean | The ratio of the ABP cardiac cycle second derivative Apeak amplitude with respect to Dpeak to the ABP cardiac cycle second derivative Cpeak amplitude with respect to zero, mean across cardiac cycles |
| ABP_AM_RT_DIAPE_wrtZero_ABP_DIAN_wrtSYSPO_mean | The ratio of the ABP cardiac cycle diastolic phase end point amplitude with respect to zero to the ABP cardiac cycle dicrotic notch amplitude with respect to systolic phase onset, mean across cardiac cycles |
| ABP_AM_RT_DIAPP_wrtZero_ABP_SYSPP_wrtDIAPE_mean | The ratio of the ABP cardiac cycle diastolic phase peak amplitude with respect to zero to the ABP cardiac cycle systolic phase peak amplitude with respect to diastolic phase end point, mean across cardiac cycles |
| ABP_AM_RT_DIAPP_wrtDIAN_ABP_SYSPP_wrtDIAPP_mean | The ratio of the ABP cardiac cycle diastolic phase peak amplitude with respect to dicrotic notch to the ABP cardiac cycle systolic phase peak amplitude with respect to diastolic phase peak, mean across cardiac cycles |

| | |
|---|---|
| ABP_D_Are_SYSPO_wrtDIAPP_mean | Area under the ABP cardiac cycle systolic phase onset with respect to diastolic phase peak, mean across cardiac cycles |
| ABP_AM_DIAN_wrtDIAPE_mean | ABP cardiac cycle dicrotic notch amplitude with respect to diastolic phase end point, mean across cardiac cycles |
| ABP_D_RT_DIAN_wrtDIAPP_ABP_SYSPO_wrtDIAPP_mean | The ratio of the ABP cardiac cycle dicrotic notch duration with respect to diastolic phase peak to the ABP cardiac cycle systolic phase onset duration with respect to diastolic phase peak, mean across cardiac cycles |
| ABP_AM_RT_SD_Apeak_wrtCpeak_ABP_AM_SD_Apeak_wrtzero_mean | The ratio of the ABP cardiac cycle second derivative Apeak amplitude with respect to Cpeak to the ABP cardiac cycle second derivative Apeak amplitude with respect to zero, mean across cardiac cycles |
| ABP_AM_RT_SD_Cpeak_wrtzero_ABP_AM_SD_Apeak_wrtzero_mean | The ratio of the ABP cardiac cycle second derivative Cpeak amplitude with respect to zero to the ABP cardiac cycle second derivative Apeak amplitude with respect to zero, mean across cardiac cycles |
| ABP_AM_DIAN_wrtSYSPO_mean | ABP cardiac cycle dicrotic notch amplitude with respect to systolic phase onset, mean across cardiac cycles |
| ABP_D_Are_wsDIAPE_SYSPO_wrtDIAPP_mean | Area under the ABP cardiac cycle systolic phase onset with respect to diastolic phase peak, with subtracted diastolic phase end point, mean across cardiac cycles |
| ABP_AM_RT_SD_Bpeak_wrtDpeak_ABP_AM_SD_Apeak_wrtDpeak_mean | The ratio of the ABP cardiac cycle second derivative Bpeak amplitude with respect to Dpeak to the ABP cardiac cycle second derivative Apeak amplitude with respect to Dpeak, mean across cardiac cycles |
| ABP_AM_RT_DIAPE_wrtSYSPO_ABP_DIAPE_wrtZero_std | The ratio of the ABP cardiac cycle diastolic phase end point amplitude with respect to systolic phase onset to the ABP diastolic phase end point with respect to zero, standard deviation across cardiac cycles |
| ABP_AM_RT_FD_Vpeak_wrtWpeak_ABP_AM_FD_Wpeak_wrtzero_mean | The ratio of the ABP cardiac cycle first derivative Vpeak amplitude with respect to Wpeak to the ABP cardiac cycle first derivative Wpeak amplitude with respect to zero, mean across cardiac cycles |
| ABP_AM_RT_DIAPE_wrtZero_ABP_SYSPO_wrtZero_std | The ratio of the ABP cardiac cycle diastolic phase end point amplitude with respect to zero to the |

| | |
|---|---|
| | ABP cardiac cycle systolic phase onset amplitude with respect to zero, standard deviation across cardiac cycles |
| ABP_AM_RT_DIAPE_wrtSYSPO_ABP_SYSPO_wrtZero_std | The ratio of the ABP cardiac cycle diastolic phase end point amplitude with respect to systolic phase onset to the ABP cardiac cycle systolic phase onset amplitude with respect to zero, standard deviation across cardiac cycles |
| ABP_AM_RT_DIAPP_wrtSYSPO_ABP_DIAN_wrtZero_std | The ratio of the ABP cardiac cycle diastolic phase peak amplitude with respect to systolic phase onset to the ABP cardiac cycle dicrotic notch amplitude with respect to zero, standard deviation across cardiac cycles |
| ABP_D_Med_SYSPO_wrtSYSPP_std | Median duration of the ABP cardiac cycle systolic phase onset with respect to systolic phase peak, standard deviation across cardiac cycles |
| ABP_AM_RT_DIAPP_wrtZero_ABP_SYSPP_wrtZero_mean | The ratio of the ABP cardiac cycle diastolic phase peak amplitude with respect to zero to the ABP cardiac cycle systolic phase peak amplitude with respect to zero, mean across cardiac cycles |
| ABP_AM_RT_SYSPP_wrtDIAPP_ABP_SYSPP_wrtZero_mean | The ratio of the ABP cardiac cycle systolic phase peak amplitude with respect to diastolic phase peak to the ABP cardiac cycle systolic phase peak amplitude with respect to zero, mean across cardiac cycles |
| ABP_AM_SD_Dpeak_wrtzero_mean | ABP cardiac cycle second derivative Dpeak amplitude with respect to zero, mean across cardiac cycles |
| ABP_AM_RT_SD_Cpeak_wrtEpeak_ABP_AM_SD_Bpeak_wrtzero_mean | The ratio of the ABP cardiac cycle second derivative Cpeak amplitude with respect to Epeak to the ABP cardiac cycle second derivative Bpeak amplitude with respect to zero, mean across cardiac cycles |
| ABP_AM_SD_Cpeak_wrtzero_mean | ABP cardiac cycle second derivative Cpeak amplitude with respect to zero, mean across cardiac cycles |
| ABP_AM_RT_SD_Cpeak_wrtEpeak_ABP_AM_SD_Cpeak_wrtDpeak_mean | The ratio of the ABP cardiac cycle second derivative Cpeak amplitude with respect to Epeak to the ABP cardiac cycle second derivative Cpeak amplitude with respect to Dpeak, mean across cardiac cycles |
| ABP_AM_RT_SD_Dpeak_wrtEpeak_ABP_AM_SD_Cpeak_wrtDpeak_mean | The ratio of the ABP cardiac cycle second derivative Dpeak amplitude with respect to Epeak to the ABP cardiac cycle second derivative Cpeak |

| | |
|---|---|
| | amplitude with respect to Dpeak, mean across cardiac cycles |
| ABP_AM_RT_DIAPP_wrtDIAN_ABP_SYSPP_wrtZero_mean | The ratio of the ABP cardiac cycle diastolic phase peak amplitude with respect to dicrotic notch to the ABP cardiac cycle systolic phase peak amplitude with respect to zero, mean across cardiac cycles |
| ABP_AM_SYSPP_wrtZero_std | ABP cardiac cycle systolic phase peak amplitude with respect to zero, standard deviation across cardiac cycles |
| Systolic_Pressure_value_std | Standard deviation across cardiac cycles of the systolic blood pressure |
| ABP_AM_RT_SD_Epeak_wrtzero_ABP_AM_SD_Cpeak_wrtzero_mean | The ratio of the ABP cardiac cycle second derivative Epeak amplitude with respect to zero to the ABP cardiac cycle second derivative Cpeak amplitude with respect to zero, mean across cardiac cycles |
| ABP_AM_RT_SD_Cpeak_wrtEpeak_ABP_AM_SD_Cpeak_wrtzero_mean | The ratio of the ABP cardiac cycle second derivative Cpeak amplitude with respect to Epeak to the ABP cardiac cycle second derivative Cpeak amplitude with respect to zero, mean across cardiac cycles |
| ABP_AM_RT_SD_Bpeak_wrtEpeak_ABP_AM_SD_Apeak_wrtDpeak_mean | The ratio of the ABP cardiac cycle second derivative Bpeak amplitude with respect to Epeak to the ABP cardiac cycle second derivative Apeak amplitude with respect to Dpeak, mean across cardiac cycles |
| ABP_AM_RT_DIAN_wrtZero_ABP_SYSPO_wrtZero_mean | The ratio of the ABP cardiac cycle dicrotic notch amplitude with respect to zero to the ABP cardiac cycle systolic phase onset amplitude with respect to zero, mean across cardiac cycles |
| ABP_AM_RT_DIAN_wrtSYSPO_ABP_SYSPO_wrtZero_mean | The ratio of the ABP cardiac cycle dicrotic notch amplitude with respect to systolic phase onset to the ABP cardiac cycle systolic phase onset amplitude with respect to zero, mean across cardiac cycles |
| ABP_AM_RT_DIAPP_wrtDIAN_ABP_DIAPP_wrtZero_std | The ratio of the ABP cardiac cycle diastolic phase peak amplitude with respect to dicrotic notch to the ABP diastolic phase peak with respect to zero, standard deviation across cardiac cycles |
| ABP_RT_ODP25_W_SRP25_ODP25_W_mean | The ratio of the ABP cardiac cycle overall decay phase width at 25% of the cardiac cycle to the sum of the ABP cardiac cycle systolic rise phase width |

| | |
|---|---|
| | and overall decay phase width at 25% of the cardiac cycle, mean across cardiac cycles |
| ABP_AM_RT_SD_Bpeak_wrtCpeak_ABP_AM_SD_Apeak_wrtzero_mean | The ratio of the ABP cardiac cycle second derivative Bpeak amplitude with respect to Cpeak to the ABP cardiac cycle second derivative Apeak amplitude with respect to zero, mean across cardiac cycles |
| ABP_AM_RT_SD_Bpeak_wrtEpeak_ABP_AM_SD_Cpeak_wrtzero_mean | The ratio of the ABP cardiac cycle second derivative Bpeak amplitude with respect to Epeak to the ABP cardiac cycle second derivative Cpeak amplitude with respect to zero, mean across cardiac cycles |
| ABP_W_SRP75_ODP75_mean | Sum of the ABP cardiac cycle systolic rise phase width and overall decay phase width at 75% of the cardiac cycle, mean across cardiac cycles |
| ABP_AM_RT_SD_Cpeak_wrtDpeak_ABP_AM_SD_Dpeak_wrtzero_mean | The ratio of the ABP cardiac cycle second derivative Cpeak amplitude with respect to Dpeak to the ABP cardiac cycle second derivative Dpeak amplitude with respect to zero, mean across cardiac cycles |
| ABP_AM_RT_DIAPE_wrtZero_ABP_SYSPO_wrtZero_mean | The ratio of the ABP cardiac cycle diastolic phase end point amplitude with respect to zero to the ABP cardiac cycle systolic phase onset amplitude with respect to zero, mean across cardiac cycles |
| ABP_AM_RT_DIAPE_wrtSYSPO_ABP_SYSPO_wrtZero_mean | The ratio of the ABP cardiac cycle diastolic phase end point amplitude with respect to systolic phase onset to the ABP cardiac cycle systolic phase onset amplitude with respect to zero, mean across cardiac cycles |
| ABP_RT_SRP25_W_SRP25_ODP25_W_mean | The ratio of the ABP cardiac cycle systolic rise phase width at 25% of the cardiac cycle to the sum of the ABP cardiac cycle systolic rise phase width and overall decay phase width at 25% of the cardiac cycle, mean across cardiac cycles |
| ABP_AM_RT_SD_Apeak_wrtBpeak_ABP_AM_SD_Cpeak_wrtzero_mean | The ratio of the ABP cardiac cycle second derivative Apeak amplitude with respect to Bpeak to the ABP cardiac cycle second derivative Cpeak amplitude with respect to zero, mean across cardiac cycles |
| ABP_AM_RT_DIAPP_wrtSYSPO_ABP_SYSPO_wrtZero_std | The ratio of the ABP cardiac cycle diastolic phase peak amplitude with respect to systolic phase onset to the ABP cardiac cycle systolic phase onset amplitude with respect to zero, standard deviation across cardiac cycles |

| | |
|---|---|
| ABP_AM_RT_DIAPP_wrtZero_ABP_SYSPO_wrtZero_std | The ratio of the ABP cardiac cycle diastolic phase peak amplitude with respect to zero to the ABP cardiac cycle systolic phase onset amplitude with respect to zero, standard deviation across cardiac cycles |
| ABP_W_SRP66_mean | ABP cardiac cycle systolic rise phase width at 66% of the cardiac cycle, mean across cardiac cycles |
| ABP_D_Med_SYSPO_wrtDIAN_mean | Median duration of the ABP cardiac cycle systolic phase onset with respect to dicrotic notch, mean across cardiac cycles |
| ABP_D_Are_SYSPO_wrtSYSPP_mean | Area under the ABP cardiac cycle systolic phase onset with respect to systolic phase peak, mean across cardiac cycles |
| ABP_VAge1_mean | ABP cardiac cycle second derivative feature defined as: (Bpeak − Cpeak − Dpeak − Epeak)/Apeak, with all peak amplitudes measured relative to the zero baseline and the ratio, averaged across cardiac cycles |
| ABP_AM_RT_SD_Cpeak_wrtEpeak_ABP_AM_SD_Apeak_wrtDpeak_mean | The ratio of the ABP cardiac cycle second derivative Cpeak amplitude with respect to Epeak to the ABP cardiac cycle second derivative Apeak amplitude with respect to Dpeak, mean across cardiac cycles |
| ABP_D_Med_SYSPO_wrtSYSPP_mean | Median duration of the ABP cardiac cycle systolic phase onset with respect to systolic phase peak, mean across cardiac cycles |
| ABP_D_RMS_SYSPO_wrtDIAN_std | Root mean square duration of the ABP cardiac cycle systolic phase onset with respect to dicrotic notch, standard deviation across cardiac cycles |
| ABP_NAM_DIAPP_wrtDIAN_mean | Normalized ABP cardiac cycle diastolic phase peak amplitude with respect to dicrotic notch, mean across cardiac cycles |
| ABP_AM_DIAPP_wrtDIAN_mean | ABP cardiac cycle diastolic phase peak amplitude with respect to dicrotic notch, mean across cardiac cycles |
| ABP_D_Med_SYSPO_wrtDIAN_std | Median duration of the ABP cardiac cycle systolic phase onset with respect to dicrotic notch, standard deviation across cardiac cycles |
| ABP_AM_RT_SD_Cpeak_wrtEpeak_ABP_AM_SD_Apeak_wrtEpeak_mean | The ratio of the ABP cardiac cycle second derivative Cpeak amplitude with respect to Epeak to the ABP cardiac cycle second derivative Apeak amplitude with respect to Epeak, mean across cardiac cycles |

| | |
|---|---|
| ABP_D_Avg_wsDIAPE_DIAN_wrtDIAPP_mean | Average duration of the ABP cardiac cycle dicrotic notch with respect to diastolic phase peak, with subtracted diastolic phase end point, mean across cardiac cycles |
| ABP_AM_RT_SD_Bpeak_wrtEpeak_ABP_AM_SD_Bpeak_wrtzero_mean | The ratio of the ABP cardiac cycle second derivative Bpeak amplitude with respect to Epeak to the ABP cardiac cycle second derivative Bpeak amplitude with respect to zero, mean across cardiac cycles |
| ABP_AM_RT_SD_Epeak_wrtzero_ABP_AM_SD_Bpeak_wrtzero_mean | The ratio of the ABP cardiac cycle second derivative Epeak amplitude with respect to zero to the ABP cardiac cycle second derivative Bpeak amplitude with respect to zero, mean across cardiac cycles |
| ABP_RT_SRP10_W_SRP10_ODP10_W_mean | The ratio of the ABP cardiac cycle systolic rise phase width at 10% of the cardiac cycle to the sum of the ABP cardiac cycle systolic rise phase width and overall decay phase width at 10% of the cardiac cycle, mean across cardiac cycles |
| ABP_NAM_RT_DIAPP_wrtDIAN_ABP_SYSPP_wrtDIAPP_mean | The ratio of the normalized ABP cardiac cycle diastolic phase peak amplitude with respect to dicrotic notch to the normalized ABP cardiac cycle systolic phase peak amplitude with respect to diastolic phase peak, mean across cardiac cycles |
| ABP_NAM_RT_SYSPP_wrtDIAPE_ABP_SYSPO_wrtZero_std | The ratio of the normalized ABP cardiac cycle systolic phase peak amplitude with respect to diastolic phase end point to the normalized ABP cardiac cycle systolic phase onset amplitude with respect to zero, standard deviation across cardiac cycles |
| ABP_D_Are_wsDIAPE_SYSPO_wrtSYSPP_mean | Area under the ABP cardiac cycle systolic phase onset with respect to systolic phase peak, with subtracted diastolic phase end point, mean across cardiac cycles |
| ABP_D_RT_Are_wsDIAPE_SYSPO_wrtDIAN_ABP_D_Are_wsDIAPE_SYSPO_wrtSYSPP_mean | The ratio of the area under the ABP cardiac cycle systolic phase onset with respect to dicrotic notch, with subtracted diastolic phase end point to the area under the ABP cardiac cycle systolic phase onset with respect to systolic phase peak, with subtracted diastolic phase end point, mean across cardiac cycles |
| ABP_NAM_RT_SYSPP_wrtDIAPP_ABP_SYSPO_wrtZero_mean | The ratio of the normalized ABP cardiac cycle systolic phase peak amplitude with respect to diastolic phase peak to the normalized ABP cardiac |

| | |
|---|---|
| | cycle systolic phase onset amplitude with respect to zero, mean across cardiac cycles |
| ABP_AM_RT_SD_Cpeak_wrtEpeak_ABP_AM_SD_Bpeak_wrtDpeak_mean | The ratio of the ABP cardiac cycle second derivative Cpeak amplitude with respect to Epeak to the ABP cardiac cycle second derivative Bpeak amplitude with respect to Dpeak, mean across cardiac cycles |
| ABP_RT_ODP10_W_SRP10_ODP10_W_mean | The ratio of the ABP/ cardiac cycle overall decay phase width at 10% of the cardiac cycle to the sum of the ABP cardiac cycle systolic rise phase width and overall decay phase width at 10% of the cardiac cycle, mean across cardiac cycles |
| ABP_D_FD_Upeak_wrtWpeak_mean | Duration of the ABP cardiac cycle first derivative Upeak with respect to Wpeak, mean across cardiac cycles |
| ABP_D_RMS_SYSPP_wrtDIAPP_std | Root mean square duration of the ABP cardiac cycle systolic phase peak with respect to diastolic phase peak, standard deviation across cardiac cycles |
| ABP_D_RT_NAre_SYSPP_wrtDIAPE_ABP_D_NAre_SYSPO_wrtDIAPE_mean | The ratio of the area under the normalized ABP cardiac cycle systolic phase peak with respect to diastolic phase end point to the area under the normalized ABP cardiac cycle systolic phase onset with respect to diastolic phase end point, mean across cardiac cycles |
| ABP_Slope_DIAPP_DIAPE_std | Standard deviation across cardiac cycles of the slope between the diastolic phase peak and diastolic phase endpoint of the ABP cardiac cycle |
| ABP_AM_SD_Cpeak_wrtEpeak_std | ABP cardiac cycle second derivative Cpeak amplitude with respect to Epeak, standard deviation across cardiac cycles |
| ABP_NAM_RT_SYSPP_wrtZero_ABP_SYSPO_wrtZero_std | The ratio of the normalized ABP cardiac cycle systolic phase peak amplitude with respect to zero to the normalized ABP cardiac cycle systolic phase onset amplitude with respect to zero, standard deviation across cardiac cycles |
| ABP_NAM_RT_SYSPP_wrtSYSPO_ABP_SYSPO_wrtZero_std | The ratio of the normalized ABP cardiac cycle systolic phase peak amplitude with respect to systolic phase onset to the normalized ABP cardiac cycle systolic phase onset amplitude with respect to zero, standard deviation across cardiac cycles |
| ABP_D_RT_NAre_wsDIAPE_SYSPP_wrtDIAPE_ABP_D_NAre_wsDIAPE_SYSPO_wrtDIAPE_mean | The ratio of the area under the normalized ABP cardiac cycle systolic phase peak with respect to diastolic phase end point, with subtracted diastolic |

| | |
|---|---|
| | phase end point to the area under the normalized ABP cardiac cycle systolic phase onset with respect to diastolic phase end point, with subtracted diastolic phase end point, mean across cardiac cycles |
| ABP_pkfrq_mean | The frequency corresponding to the first peak in the FFT of the ABP cardiac cycle, mean across cardiac cycles |
| ABP_D_Med_SYSPO_wrtDIAPP_std | Median duration of the ABP cardiac cycle systolic phase onset with respect to diastolic phase peak, standard deviation across cardiac cycles |
| ABP_D_NAvg_SYSPO_wrtSYSPP_mean | Average duration of the normalized ABP cardiac cycle systolic phase onset with respect to systolic phase peak, mean across cardiac cycles |
| ABP_D_NAre_wsDIAPE_SYSPO_wrtDIAPP_mean | Area under the normalized ABP cardiac cycle systolic phase onset with respect to diastolic phase peak, with subtracted diastolic phase end point, mean across cardiac cycles |
| ABP_D_RMS_SYSPO_wrtSYSPP_std | Root mean square duration of the ABP cardiac cycle systolic phase onset with respect to systolic phase peak, standard deviation across cardiac cycles |
| ABP_D_RT_Are_wsDIAPE_SYSPP_wrtDIAPE_ABP_D_Are_wsDIAPE_SYSPO_wrtSYSPP_mean | The ratio of the area under the ABP cardiac cycle systolic phase peak with respect to diastolic phase end point, with subtracted diastolic phase end point to the area under the ABP cardiac cycle systolic phase onset with respect to systolic phase peak, with subtracted diastolic phase end point, mean across cardiac cycles |
| ABP_D_NAre_SYSPO_wrtDIAPP_mean | Area under the normalized ABP cardiac cycle systolic phase onset with respect to diastolic phase peak, mean across cardiac cycles |
| ABP_AM_RT_SD_Bpeak_wrtEpeak_ABP_AM_SD_Bpeak_wrtDpeak_mean | The ratio of the ABP cardiac cycle second derivative Bpeak amplitude with respect to Epeak to the ABP cardiac cycle second derivative Bpeak amplitude with respect to Dpeak, mean across cardiac cycles |
| ABP_AM_RT_SD_Dpeak_wrtEpeak_ABP_AM_SD_Bpeak_wrtDpeak_mean | The ratio of the ABP cardiac cycle second derivative Dpeak amplitude with respect to Epeak to the ABP cardiac cycle second derivative Bpeak amplitude with respect to Dpeak, mean across cardiac cycles |
| ABP_D_RMS_SYSPO_wrtDIAPP_std | Root mean square duration of the ABP cardiac cycle systolic phase onset with respect to diastolic |

| | |
|---|---|
| | phase peak, standard deviation across cardiac cycles |
| ABP_AM_RT_SD_Bpeak_wrtDpeak_ABP_AM_SD_Apeak_wrtzero_mean | The ratio of the ABP cardiac cycle second derivative Bpeak amplitude with respect to D peak to the ABP cardiac cycle second derivative Apeak amplitude with respect to zero, mean across cardiac cycles |
| ABP_NAM_RT_DIAPE_wrtSYSPO_ABP_SYSPO_wrtZero_mean | The ratio of the normalized ABP cardiac cycle diastolic phase end point amplitude with respect to systolic phase onset to the normalized ABP cardiac cycle systolic phase onset amplitude with respect to zero, mean across cardiac cycles |
| ABP_NAM_RT_DIAPE_wrtZero_ABP_SYSPO_wrtZero_mean | The ratio of the normalized ABP cardiac cycle diastolic phase end point with respect to zero to the normalized ABP cardiac cycle systolic phase onset amplitude with respect to zero, mean across cardiac cycles |
| ABP_D_NAre_wsDIAPE_SYSPO_wrtDIAN_mean | Area under the normalized ABP cardiac cycle systolic phase onset with respect to dicrotic notch, with subtracted diastolic phase end point, mean across cardiac cycles |
| ABP_AM_RT_DIAPP_wrtSYSPO_ABP_DIAPP_wrtZero_std | The ratio of the ABP cardiac cycle diastolic phase peak amplitude with respect to systolic phase onset to the ABP/ diastolic phase peak with respect to zero, standard deviation across cardiac cycles |
| ABP_D_NAre_SYSPO_wrtDIAN_mean | Area under the normalized ABP cardiac cycle systolic phase onset with respect to dicrotic notch, mean across cardiac cycles |
| ABP_D_RT_Are_wsDIAPE_SYSPO_wrtDIAPE_ABP_D_Are_wsDIAPE_SYSPO_wrtSYSPP_mean | The ratio of the area under the ABP cardiac cycle systolic phase onset with respect to diastolic phase end point, with subtracted diastolic phase end point to the area under the ABP cardiac cycle systolic phase onset with respect to systolic phase peak, with subtracted diastolic phase end point, mean across cardiac cycles |
| ABP_D_RT_Are_DIAN_wrtDIAPP_ABP_D_Are_SYSPO_wrtDIAPE_mean | The ratio of the area under the ABP cardiac cycle dicrotic notch with respect to diastolic phase peak to the area under the ABP cardiac cycle systolic phase onset with respect to diastolic phase end point, mean across cardiac cycles |
| ABP_D_Med_SYSPP_wrtDIAPP_std | Median duration of the ABP cardiac cycle systolic phase peak with respect to diastolic phase peak, standard deviation across cardiac cycles |

| | |
|---|---|
| ABP_D_RT_Are_wsDIAPE_DIAN_wrtDIAPP_ABP_D_Are_wsDIAPE_SYSPO_wrtDIAPE_mean | The ratio of the area under the ABP cardiac cycle dicrotic notch with respect to diastolic phase peak, with subtracted diastolic phase end point to the area under the ABP cardiac cycle systolic phase onset with respect to diastolic phase end point, with subtracted diastolic phase end point, mean across cardiac cycles |
| ABP_AM_RT_SD_Bpeak_wrtzero_ABP_AM_SD_Apeak_wrtzero_mean | The ratio of the ABP cardiac cycle second derivative Bpeak amplitude with respect to zero to the ABP cardiac cycle second derivative Apeak amplitude with respect to zero, mean across cardiac cycles |
| ABP_AM_RT_SD_Apeak_wrtBpeak_ABP_AM_SD_Apeak_wrtzero_mean | The ratio of the ABP cardiac cycle second derivative Apeak amplitude with respect to Bpeak to the ABP cardiac cycle second derivative Apeak amplitude with respect to zero, mean across cardiac cycles |
| ABP_AM_FD_Upeak_wrtVpeak_mean | ABP cardiac cycle first derivative Upeak amplitude with respect to Vpeak, mean across cardiac cycles |
| ABP_D_NAre_wsDIAPE_SYSPO_wrtSYSPP_mean | Area under the normalized ABP cardiac cycle systolic phase onset with respect to systolic phase peak, with subtracted diastolic phase end point, mean across cardiac cycles |
| ABP_AM_RT_SD_Bpeak_wrtCpeak_ABP_AM_SD_Apeak_wrtCpeak_mean | The ratio of the ABP cardiac cycle second derivative Bpeak amplitude with respect to Cpeak to the ABP cardiac cycle second derivative Apeak amplitude with respect to Cpeak, mean across cardiac cycles |
| ABP_AM_DIAPP_wrtSYSPO_std | ABP cardiac cycle diastolic phase peak amplitude with respect to systolic phase onset, standard deviation across cardiac cycles |
| ABP_D_Avg_wsDIAPE_DIAN_wrtDIAPP_std | Average duration of the ABP cardiac cycle dicrotic notch with respect to diastolic phase peak, with subtracted diastolic phase end point, standard deviation across cardiac cycles |
| ABP_AM_RT_SD_Apeak_wrtCpeak_ABP_AM_SD_Bpeak_wrtzero_mean | The ratio of the ABP cardiac cycle second derivative Apeak amplitude with respect to Cpeak to the ABP cardiac cycle second derivative Bpeak amplitude with respect to zero, mean across cardiac cycles |
| ABP_AM_RT_DIAN_wrtZero_ABP_SYSPP_wrtDIAPP_mean | The ratio of the ABP cardiac cycle dicrotic notch amplitude with respect to zero to the ABP cardiac cycle systolic phase peak amplitude with respect to diastolic phase peak, mean across cardiac cycles |

| | |
|---|---|
| ABP_AM_RT_DIAPE_wrtSYSPO_ABP_DIAN_wrtZero_mean | The ratio of the ABP cardiac cycle diastolic phase end point amplitude with respect to systolic phase onset to the ABP cardiac cycle dicrotic notch amplitude with respect to zero, mean across cardiac cycles |
| ABP_AM_RT_SD_Cpeak_wrtEpeak_ABP_AM_SD_Apeak_wrtBpeak_mean | The ratio of the ABP cardiac cycle second derivative Cpeak amplitude with respect to Epeak to the ABP cardiac cycle second derivative Apeak amplitude with respect to Bpeak, mean across cardiac cycles |
| ABP_NAM_RT_SYSPP_wrtDIAPE_ABP_SYSPO_wrtZero_mean | The ratio of the normalized ABP cardiac cycle systolic phase peak amplitude with respect to diastolic phase end point to the normalized ABP cardiac cycle systolic phase onset amplitude with respect to zero, mean across cardiac cycles |
| ABP_AM_RT_SD_Bpeak_wrtCpeak_ABP_AM_SD_Apeak_wrtDpeak_mean | The ratio of the ABP cardiac cycle second derivative Bpeak amplitude with respect to Cpeak to the ABP cardiac cycle second derivative Apeak amplitude with respect to Dpeak, mean across cardiac cycles |
| ABP_AM_RT_SD_Cpeak_wrtEpeak_ABP_AM_SD_Bpeak_wrtCpeak_mean | The ratio of the ABP cardiac cycle second derivative Cpeak amplitude with respect to Epeak to the ABP cardiac cycle second derivative Bpeak amplitude with respect to Cpeak, mean across cardiac cycles |
| ABP_D_RT_Are_SYSPP_wrtDIAPE_ABP_D_Are_SYSPO_wrtSYSPP_mean | The ratio of the area under the ABP cardiac cycle systolic phase peak with respect to diastolic phase end point to the area under the ABP cardiac cycle systolic phase onset with respect to systolic phase peak, mean across cardiac cycles |
| ABP_D_RT_Are_SYSPO_wrtDIAPE_ABP_D_Are_SYSPO_wrtSYSPP_mean | The ratio of the area under the ABP cardiac cycle systolic phase onset with respect to diastolic phase end point to the area under the ABP cardiac cycle systolic phase onset with respect to systolic phase peak, mean across cardiac cycles |
| ABP_AM_RT_SD_Bpeak_wrtEpeak_ABP_AM_SD_Bpeak_wrtCpeak_mean | The ratio of the ABP cardiac cycle second derivative Bpeak amplitude with respect to Epeak to the ABP cardiac cycle second derivative Bpeak amplitude with respect to Cpeak, mean across cardiac cycles |
| ABP_D_RT_NAre_SYSPP_wrtDIAPE_ABP_D_NAre_SYSPO_wrtSYSPP_mean | The ratio of the area under the normalized ABP cardiac cycle systolic phase peak with respect to diastolic phase end point to the area under the normalized ABP cardiac cycle systolic phase onset |

| | |
|---|---|
| | with respect to systolic phase peak, mean across cardiac cycles |
| ABP_D_RT_NAre_SYSPO_wrtDIAPE_ABP_D_NAre_SYSPO_wrtSYSPP_mean | The ratio of the area under the normalized ABP cardiac cycle systolic phase onset with respect to diastolic phase end point to the area under the normalized ABP cardiac cycle systolic phase onset with respect to systolic phase peak, mean across cardiac cycles |
| ABP_D_Avg_SYSPO_wrtSYSPP_std | Average duration of the ABP cardiac cycle systolic phase onset with respect to systolic phase peak, standard deviation across cardiac cycles |
| ABP_D_Med_SYSPP_wrtDIAN_mean | Median duration of the ABP cardiac cycle systolic phase peak with respect to dicrotic notch, mean across cardiac cycles |
| ABP_D_RT_SYSPP_wrtDIAN_ABP_SYSPO_wrtDIAN_mean | The ratio of the ABP cardiac cycle systolic phase peak duration with respect to dicrotic notch to the ABP cardiac cycle systolic phase onset duration with respect to dicrotic notch, mean across cardiac cycles |
| ABP_D_NAre_SYSPO_wrtSYSPP_mean | Area under the normalized ABP cardiac cycle systolic phase onset with respect to systolic phase peak, mean across cardiac cycles |
| ABP_AM_RT_DIAPE_wrtZero_ABP_SYSPP_wrtDIAPP_mean | The ratio of the ABP cardiac cycle diastolic phase end point amplitude with respect to zero to the ABP cardiac cycle systolic phase peak amplitude with respect to diastolic phase peak, mean across cardiac cycles |
| ABP_D_Are_SYSPP_wrtDIAPP_mean | Area under the ABP cardiac cycle systolic phase peak with respect to diastolic phase peak, mean across cardiac cycles |
| ABP_AM_RT_DIAN_wrtZero_ABP_SYSPP_wrtDIAPE_std | The ratio of the ABP cardiac cycle dicrotic notch amplitude with respect to zero to the ABP cardiac cycle systolic phase peak amplitude with respect to diastolic phase end point, standard deviation across cardiac cycles |
| ABP_W_ODP33_std | ABP cardiac cycle overall decay phase width at 33% of the cardiac cycle, standard deviation across cardiac cycles |
| ABP_AM_RT_SD_Bpeak_wrtDpeak_ABP_AM_SD_Cpeak_wrtzero_mean | The ratio of the ABP cardiac cycle second derivative Bpeak amplitude with respect to Dpeak to the ABP cardiac cycle second derivative Cpeak amplitude with respect to zero, mean across cardiac cycles |

| | |
|---|---|
| ABP_D_RT_NAre_wsDIAPE_DIAPP_wrtDIAPE_ABP_D_NAre_wsDIAPE_SYSPO_wrtSYSPP_mean | The ratio of the area under the normalized ABP cardiac cycle diastolic phase peak with respect to diastolic phase end point, with subtracted diastolic phase end point to the area under the normalized ABP cardiac cycle systolic phase onset with respect to systolic phase peak, with subtracted diastolic phase end point, mean across cardiac cycles |
| ABP_D_RT_NAre_DIAPP_wrtDIAPE_ABP_D_NAre_SYSPO_wrtSYSPP_mean | The ratio of the area under the normalized ABP cardiac cycle diastolic phase peak with respect to diastolic phase end point to the area under the normalized ABP cardiac cycle systolic phase onset with respect to systolic phase peak, mean across cardiac cycles |
| ABP_AM_RT_SD_Bpeak_wrtDpeak_ABP_AM_SD_Apeak_wrtCpeak_mean | The ratio of the ABP cardiac cycle second derivative Bpeak amplitude with respect to Dpeak to the ABP cardiac cycle second derivative Apeak amplitude with respect to Cpeak, mean across cardiac cycles |
| ABP_D_Are_wsDIAPE_SYSPP_wrtDIAPP_mean | Area under the ABP cardiac cycle systolic phase peak with respect to diastolic phase peak, with subtracted diastolic phase end point, mean across cardiac cycles |
| ABP_RT_ODP33_W_SRP33_W_mean | The ratio of the ABP cardiac cycle overall decay phase width at 33% of the cardiac cycle to the ABP cardiac cycle systolic rise phase width at 33% of the cardiac cycle, mean across cardiac cycles |
| ABP_RT_ODP66_W_SRP66_ODP66_W_mean | The ratio of the ABP cardiac cycle overall decay phase width at 66% of the cardiac cycle to the sum of the ABP cardiac cycle systolic rise phase width and overall decay phase width at 66% of the cardiac cycle, mean across cardiac cycles |
| ABP_W_SRP50_mean | ABP cardiac cycle systolic rise phase width at 50% of the cardiac cycle, mean across cardiac cycles |
| ABP_D_NAre_DIAN_wrtDIAPP_mean | Area under the normalized ABP cardiac cycle dicrotic notch with respect to diastolic phase peak, mean across cardiac cycles |
| ABP_RT_ODP10_W_SRP10_W_mean | The ratio of the ABP cardiac cycle overall decay phase width at 10% of the cardiac cycle to the ABP cardiac cycle systolic rise phase width at 10% of the cardiac cycle, mean across cardiac cycles |
| ABP_W_SRP75_mean | ABP cardiac cycle systolic rise phase width at 75% of the cardiac cycle, mean across cardiac cycles |
| ABP_D_RT_NAre_wsDIAPE_SYSPO_wrtDIAPE_ABP_D_NAre_wsDIAPE_SYSPO_wrtSYSPP_mean | The ratio of the area under the normalized ABP cardiac cycle systolic phase onset with respect to |

| | |
|---|---|
| | diastolic phase end point, with subtracted diastolic phase end point to the area under the normalized ABP cardiac cycle systolic phase onset with respect to systolic phase peak, with subtracted diastolic phase end point, mean across cardiac cycles |
| ABP_AM_RT_SD_Cpeak_wrtEpeak_ABP_AM_SD_Bpeak_wrtEpeak_mean | The ratio of the ABP cardiac cycle second derivative Cpeak amplitude with respect to Epeak to the ABP cardiac cycle second derivative Bpeak amplitude with respect to Epeak, mean across cardiac cycles |
| ABP_D_Avg_wsDIAPE_DIAPP_wrtDIAPE_std | Average duration of the ABP cardiac cycle diastolic phase peak with respect to diastolic phase end point, with subtracted diastolic phase end point, standard deviation across cardiac cycles |
| ABP_AM_RT_SD_Cpeak_wrtEpeak_ABP_AM_SD_Apeak_wrtzero_mean | The ratio of the ABP cardiac cycle second derivative Cpeak amplitude with respect to Epeak to the ABP cardiac cycle second derivative Apeak amplitude with respect to zero, mean across cardiac cycles |
| ABP_RT_SRP66_W_SRP66_ODP66_W_mean | The ratio of the ABP cardiac cycle systolic rise phase width at 66% of the cardiac cycle to the sum of the ABP cardiac cycle systolic rise phase width and overall decay phase width at 66% of the cardiac cycle, mean across cardiac cycles |
| ABP_AM_RT_DIAPE_wrtSYSPO_ABP_SYSPP_wrtZero_mean | The ratio of the ABP cardiac cycle diastolic phase end point amplitude with respect to systolic phase onset to the ABP cardiac cycle systolic phase peak amplitude with respect to zero, mean across cardiac cycles |
| ABP_NAM_RT_DIAPP_wrtDIAN_ABP_SYSPO_wrtZero_mean | The ratio of the normalized ABP cardiac cycle diastolic phase peak amplitude with respect to dicrotic notch to the normalized ABP cardiac cycle systolic phase onset amplitude with respect to zero, mean across cardiac cycles |
| ABP_NAM_RT_DIAPP_wrtDIAN_ABP_SYSPP_wrtSYSPO_mean | The ratio of the normalized ABP cardiac cycle diastolic phase peak amplitude with respect to dicrotic notch to the normalized ABP cardiac cycle systolic phase peak amplitude with respect to systolic phase onset, mean across cardiac cycles |
| ABP_AM_RT_DIAPP_wrtDIAN_ABP_SYSPP_wrtSYSPO_mean | The ratio of the ABP cardiac cycle diastolic phase peak amplitude with respect to dicrotic notch to the ABP cardiac cycle systolic phase peak amplitude with respect to systolic phase onset, mean across cardiac cycles |

| | |
|---|---|
| ABP_NAM_RT_DIAPP_wrtDIAN_ABP_SYSPP_wrtZero_mean | The ratio of the normalized ABP cardiac cycle diastolic phase peak amplitude with respect to dicrotic notch to the normalized ABP cardiac cycle systolic phase peak amplitude with respect to zero, mean across cardiac cycles |
| ABP_D_Avg_SYSPO_wrtDIAPP_std | Average duration of the ABP cardiac cycle systolic phase onset with respect to diastolic phase peak |
| ABP_Slope_SYSPP_DIAN_mean | Mean across cardiac cycles of the slope between the systolic phase peak and dicrotic notch of the ABP cardiac cycle |
| ABP_D_RT_NAre_wsDIAPE_SYSPP_wrtDIAPE_ABP_D_NAre_wsDIAPE_SYSPO_wrtSYSPP_mean | The ratio of the area under the normalized ABP cardiac cycle systolic phase peak with respect to diastolic phase end point, with subtracted diastolic phase end point to the area under the normalized ABP cardiac cycle systolic phase onset with respect to systolic phase peak, with subtracted diastolic phase end point, mean across cardiac cycles |
| ABP_D_NMed_SYSPO_wrtSYSPP_mean | Median duration of the normalized ABP cardiac cycle systolic phase onset with respect to systolic phase peak, mean across cardiac cycles |
| ABP_AM_DIAPP_wrtDIAPE_std | ABP cardiac cycle diastolic phase peak amplitude with respect to diastolic phase end point, standard deviation across cardiac cycles |
| ABP_AM_RT_SD_Dpeak_wrtzero_ABP_AM_SD_Apeak_wrtzero_mean | The ratio of the ABP cardiac cycle second derivative Dpeak amplitude with respect to zero to the ABP cardiac cycle second derivative Apeak amplitude with respect to zero, mean across cardiac cycles |
| ABP_AM_RT_SD_Apeak_wrtDpeak_ABP_AM_SD_Apeak_wrtzero_mean | The ratio of the ABP cardiac cycle second derivative Apeak amplitude with respect to Dpeak to the ABP cardiac cycle second derivative Apeak amplitude with respect to zero, mean across cardiac cycles |
| ABP_AM_SYSPP_wrtZero_mean | ABP cardiac cycle systolic phase peak amplitude with respect to zero, mean across cardiac cycles |
| Systolic_Pressure_value_mean | Mean across cardiac cycles of the systolic blood pressure |
| ABP_W_SRP10_mean | ABP cardiac cycle systolic rise phase width at 10% of the cardiac cycle, mean across cardiac cycles |
| ABP_W_ODP10_mean | ABP cardiac cycle overall decay phase width at 10% of the cardiac cycle, mean across cardiac cycles |
| ABP_NAM_RT_DIAPP_wrtZero_ABP_DIAN_wrtDIAPE_mean | The ratio of the normalized ABP cardiac cycle diastolic phase peak amplitude with respect to |

| | |
|---|---|
| | zero to the normalized ABP cardiac cycle dicrotic notch amplitude with respect to diastolic phase end point, mean across cardiac cycles |
| ABP_AM_RT_SD_Cpeak_wrtEpeak_ABP_AM_SD_Apeak_wrtCpeak_mean | The ratio of the ABP cardiac cycle second derivative Cpeak amplitude with respect to Epeak to the ABP cardiac cycle second derivative Apeak amplitude with respect to Cpeak, mean across cardiac cycles |
| ABP_AM_RT_SD_Apeak_wrtEpeak_ABP_AM_SD_Apeak_wrtCpeak_mean | The ratio of the ABP cardiac cycle second derivative Apeak amplitude with respect to Epeak to the ABP cardiac cycle second derivative Apeak amplitude with respect to Cpeak, mean across cardiac cycles |
| ABP_AM_RT_SD_Apeak_wrtDpeak_ABP_AM_SD_Apeak_wrtBpeak_mean | The ratio of the ABP cardiac cycle second derivative Apeak amplitude with respect to Dpeak to the ABP cardiac cycle second derivative Apeak amplitude with respect to Bpeak, mean across cardiac cycles |
